# An 8 week randomized Dietary Guidelines for Americans -based diet intervention improves the omega-3 index of healthy women

**DOI:** 10.1101/2021.09.22.21263899

**Authors:** Christine E. Richardson, Sridevi Krishnan, Ira J. Gray, Nancy L. Keim, John W. Newman

## Abstract

**Background:** The Dietary Guidelines for Americans (DGA) recommends consuming >1.75g/wk of long-chain omega-3 fatty acids to reduce the risk of cardiovascular disease (CVD) through triglyceride reduction, however individual responses to treatment vary.

**Objective:** We sought to determine if a DGA-conforming diet (DGAD) can increase the omega-3 index (OM3I), a diet-sensitive biomarker of omega-3 fatty acid status, into a health promoting range and reduce fasting triglycerides in 8 weeks. We further explored determinants of the basal OM3I and its response to treatment.

**Design:** This is a secondary analysis of a randomized, double-blind 8wk dietary intervention of overweight/obese women fed an 8d rotating DGAD (n =22) or typical American diet (TAD; n =20) registered at www.clinicaltrials.gov as NCT02298725. The DGAD and TAD provided individuals with 16 ± 2 g/wk and 1.2 ± 0.12 g/wk of eisocapentaenoic acid (EPA) + docosahexaenoic acid (DHA), respectively. Habitual diet and body composition were determined at baseline. OM3I, fasting triglycerides, glucose and insulin were measured at 0, 2 and 8wk.

**Results:** Baseline OM3I (5.8 ± 1.3; n =42) was positively correlated to the dietary (EPA+DHA):dietary fat ratio (p =0.006), negatively correlated to the android fat mass (p =0.0007) and was not different between diet groups. At 8wk, while the TAD-group average OM3I was unchanged (5.8 ± 0.76), the DGAD-group OM3I increased (7.33 ± 1.36; p <0.001). In the DGAD-group 9 of 22 (i.e. 41%) participants achieving an OM3I >8%. Subgroup analyses of the DGAD-group revealed that body fat content and distribution influenced the baseline-dependent response to treatment. Fasting triglyceride and OM3I changes did not correlate.

**Conclusions:** An 8wk TAD stabilized the OM3I in a healthy range, while a DGAD increased the OM3I into a health-promoting range, but did not reduce fasting triglycerides. Fat distribution and basal omega-3 status are primary factors influencing omega-3 efficacy in overweight/obese women.

## Introduction

Cardiovascular disease (CVD) is a leading cause of mortality, accounting for nearly 18 million deaths worldwide in 2016 (https://www.who.int/news-room/fact-sheets/detail/cardiovascular-diseases-(cvds)). Diet is a modifiable factor that can influence CVD risk. The consumption of the long-chain omega-3 fatty acids eicosapentaenoic acid (EPA) and docosahexaenoic acid (DHA) for CVD risk reduction has received much attention [1, 2]. Various studies have demonstrated the cardioprotective properties of EPA and DHA, with proposed mechanisms including improvements in endothelial function, fasting triglycerides (TG), blood pressure, platelet aggregation, and inflammation [3-10]. Recently, a >3g/d dose of EPA + DHA was suggested as the level required to attain these protections [9]. However, studies have shown inconsistencies in response to omega-3 fatty acid intake, which can be attributed to differences in body weight, gender, age, and genetics, among others [11-13].

Dietary polyunsaturated fatty acids are readily incorporated into cellular phospholipids, and their incorporation into red blood cells (RBCs) combined with the average RBC life span of ∼115 days provides an accessible and time-integrated measure of dietary fatty acid intake [14, 15]. The omega-3 index (OM3I), a metric that expresses the red blood cell EPA + DHA content as a percent of total fatty acids, is a validated diet-sensitive biomarker of tissues omega-3 fatty acid levels [11, 16]. The OM3I reflects a saturable process that responds inversely to the basal status, i.e., upon supplementation, the largest change happens in individuals that start with the lowest values within that population [17]. An OM3I range of 8-12% is associated with reduced CVD risk, while values <4% suggest elevated risk [11, 18]. Therefore, dietary modulation of the OM3I by adherence to a healthful diet with high intermittent doses of omega-3 fatty acid-rich foods may provide an alternative strategy to CVD risk reduction.

The Dietary Guidelines for Americans (DGA), first published in 1980 and currently modified on a 5y cycle, aim to minimize the risk of developing metabolic diseases such as CVD by focusing on consumption of a healthy, nutrient-dense diet. Accordingly, this intervention study was planned using the 2010 DGAs, which was current at the time, that recommend a diet full of fruits, vegetables, whole grains, low-fat dairy products, and a reduction of sodium, solid fats, and added sugars. It also suggests the consumption of 8oz (i.e. 225g) per week of seafood to consume a daily average of 250mg of eicosapentaenoic acid (EPA) and docosahexaenoic acid (DHA) to enhance the omega-3 fatty acid status of the population [19]. These long-chain omega-3 fatty acids have reported anti-inflammatory and anti-hyperlipidemic properties, and increasing their intake may reduce the risk of cardiometabolic disease [20]. Common interventions assessing impacts on the omega-3 status have included the consumption of EPA + DHA-rich supplements or increasing the consumption of cold-water fatty fish or eggs and poultry raised on DHA-rich diets [21-23]. The dominant form of omega-3 fatty acids consumed in the United States is the essential fatty acid alpha-linolenic acid (ALA) [24]. While increasing the consumption of plant-based foods increases the intake of ALA, due to its low rate of conversion to EPA and DHA, it is generally believed that EPA and DHA consumption are required to meaningfully increase their levels [25]. However, the rate of ALA conversion to EPA can be significant in humans [26-28], despite a high degree of variability influenced by factors including both the absolute [26] and relative [29] amounts of dietary ALA and linoleic acid, exercise [30] and genetic variance in the desaturases and elongases involved in the process [31]. Therefore, while EPA and DHA have a strong acute effect on omega-3 status, ALA-rich diets may themselves influence this outcome. To the best of our knowledge, it is not currently known how following a diet reflecting the DGAs or a typical American diet (TAD) will influence the OM3I within 8 weeks. Moreover, it is unclear if such changes in the OM3I will correlate with changes in circulating TGs of normo-lipidemic to mildly hyperlipidemic individuals in the same time frame.

The current report describes a secondary analysis of a clinical intervention in pre-and post-menopausal women evaluating the ability of a 2010 DGA-based diet (DGAD) to beneficially alter the OM3I status and/or plasma TG levels when compared to a typical American diet (TAD) defined as the median intake reported by the Centers for Disease Control and Prevention, National Health and Nutrition Examination Survey. We hypothesize that the DGAD will increase the OM3I status and changes in the OM3I will correlate with changes in plasma TG concentrations.

## Subjects and Methods

### Study design

The present study used a randomized, double-blind, 8-week dietary intervention in overweight and obese pre-and postmenopausal females registered at www.clinicaltrials.gov as NCT02298725 conducted between Dec 2014 and Mar 2017.A detailed description of the study design has already been published [32, 33]. All procedures were approved by the University of California, Davis (UC Davis), Institutional Review Board. Briefly, volunteers were stratified by menopausal status (pre- or post-) and glucose intolerance (normal compared with insulin resistant), before random assignment in a 1:1 allocation ratio to the DGAD or TAD in blocks of two using Microsoft Office Excel. The study statistician generated the randomization lists using color to indicate treatment, and the principal dietitian assigned the colors to specific diets and implemented the randomization scheme. The randomization schedules were only communicated between the statistician and the dietitian, and the dietitian’s assistant was aware of and had access to the sequence. Both diets were designed to maintain body weight, to consist of food and beverage items commonly available at grocery stores, and to have a similar appearance. The TAD was based on the 50^th^ percentile of the NHANES’s “What We Eat in America” dietary survey and the DGAD was based on food-group recommendations by the 2010 DGAs. The DGAD consisted of more fruits, vegetables, whole grains, and dairy products, whereas the TAD consisted of more refined grains, solid fats, and added sugars. Eight-day cyclical menus were used, with each day consisting of 3 meals, snacks, and beverages designed to supply adequate calories for participants to maintain their body weight. Menu planning to achieve the estimated experimental differences in omega-3 fatty acid were performed using fatty acid estimates of commercial food products contained in the 2014 version of the Nutrition Data System for Research from the University of Minnesota - Nutrition Coordinating Center (http://www.ncc.umn.edu/products/). All meals were prepared by the Metabolic Kitchen and Human Feeding Laboratory at the USDA - Agricultural Research Service - Western Human Nutrition Research Center and volunteers picked up their meals twice weekly and returned packaging so that compliance could be checked. A more detailed description of the meal preparations, menus, and nutrition composition has been previously published [33]. All analyses were performed in a blinded fashion.

### Subjects

Pre- and postmenopausal females (n = 44) aged 20-65 years with a BMI (kg/m^2^) of 25-39.9 were recruited for this intervention. Sample size was based on the study primary outcome investigating the impact of diet quality on glucose homeostasis in women with at least one risk factor for the metabolic syndrome [33].Sententary women or those with low physical activity levels were recruited with inclusion criteria below the physical activity guidelines of moderate activity of 150 min/week, a resting blood pressure of ≤140/90 mmHg, and if they had 1 or more of the following clinical measures of glucose homeostasis or lipid metabolism: 100-126 mg/dL fasting glucose; 140-199mg/dL oral glucose tolerance test 2-h glucose; Quantitative Insulin Sensitivity Check Index score <0.315 [34]; homeostasis model assessment of insulin resistance (HOMA-IR) >3.67; HbA1c between 5.7-6.5, fasting triglycerides >150mg/dL; high density lipoprotein cholesterol <50mg/dL. Exclusion criteria included: resting blood pressure >140/90mmHg; hemoglobin <11.5g/dL; total cholesterol >300mg/dL; low density lipoprotein cholesterol >189mg/dL; triglycerides >400mg/dL; clinically diagnosed abnormal thyroid or liver function; the presence of any metabolic disease, gastrointestinal disorders, cancer or other serious chronic diseases; pregnancy or lactation; use of tobacco; use of medications for elevated lipids or glucose; regular use of prescription or over-the-counter medications in the 6mo before enrolling into the study; moderate or strenuous physical activity >30 min/d on ≥5 d/wk; weight change of >5% of body weight in the 6mo before enrolling into the study; working “graveyard” shifts or forced regular all-night wake cycles; dietary restrictions interfering with the intervention foods.

### Food Frequency Questionnaires

To evaluate the impact of habitual diet on the basal OM3I, dietary intake estimates were assessed using Block Food Frequency questionnaires (FFQs). Each participant completed an FFQ approximately 1 to 2 weeks prior to the initiation of the intervention, and the responses were used to reflect habitual dietary intake.

### Body Composition

To allow for the analysis of body composition impacts on basal OM3I, the OM3I response to treatment, and as a compounding factor on fasting triglyceride levels, dual-energy x-ray absorptiometry (DEXA) were performed. Once during baseline (Wk0) and once during week 8 (Wk8), a whole-body DEXA scan was performed using a Hologic® Discovery™ QDR® Series 84994 (Hologic, Inc.). This scan provided values for total lean mass, total fat mass, percent body fat, and estimates of gynoid and android fat distribution.

### Blood collection and clinical chemistry

Fasting blood samples were collected by a licensed phlebotomist, directly into EDTA containing vacutainers, mixed gently, and centrifuged at 1300rcf for 10min at 4°C within 30min of blood collection. After plasma and buffy coat removal, RBCs from a 5mL blood tube were suspended in 14mL phosphate-buffered saline, gently mixed, and pelleted by centrifugation as described above. The washing procedure was repeated and the resulting washed RBCs were transferred to polypropylene Eppendorf tubes and stored at -70°C until analysis.

### Fatty acid analyses

To determine dietary fatty acid exposures and assess their impact on red blood cell fatty acid composition, composites of each experimental meal and red blood cell fatty acids were quantified as fatty acids methyl esters (FAMEs) by gas chromatography-mass spectrometry. FAMEs were quantified against authentic standards correcting for methodological recoveries of ∼5µmol of d31-tripalmitoylglyceride (Avanti Polar Lipids, Alabaster AL) [35, 36]. Data quality assurance / quality control measures included sample randomization, the inclusion of procedural blanks, duplication of 5% of samples, concurrent analysis of laboratory reference materials in each analytical batch, and the use of isotopically labeled extraction surrogates.

Briefly, fatty acids in ∼50 mg meal composites or 50 µL of phosphate-buffered saline-washed RBCs were extracted with 10:8:11 cylcohexane:isopropanol: 0.1M ammonium acetate in the presence of deuterated analytical surrogates [37]. Meal composite and RBC extracted lipid residues were reconstituted in either 1 mL or 200µL of 1:1 methanol/toluene (v/v), respectively, and stored at -20°C until derivatization. Isolated lipids were transformed into FAMEs by sequential incubation with methanolic sodium hydroxide and methanolic hydrochloric acid. Specifically, 25 µL of meal composite extract or 20 µL of RBC extracts were further diluted with 20µL of 1:1 toluene/methanol (v/v), enriched with 20 µL of 60 µM 10*Z*-pentadecenoic acid in methanol, diluted with 140 µL methanol, mixed with 100 µL of 0.5 M sodium methoxide, headspace flushed with nitrogen and incubated at 60 °C for 1 h. Samples were cooled 5 min before addition of 100 μL of 3 N methanolic hydrochloric acid, and returned to 60 °C for 30 min. Reaction mixtures were then neutralized with 400 µL potassium carbonate, mixed with 400 µL saturated sodium chloride, and back extracted with 400 µL hexane.

FAMEs were separated on a 30m × 0.25mm, 0.25 µm DB-225ms on 7890B gas chromatograph interfaced with a 5977B mass selective detector (Agilent Technologies) and quantified against 6-to 8-point calibration curves. Calibrants and internal standards were purchased from NuchekPrep (Elysian, MN) or Sigma-Aldrich (St. Louis, MO). Data were acquired and processed with MassHunter v. 3.2. Results were corrected for d31-tripalmitoylglyceride recoveries which were 47 ± 15% and 49 ± 12% for meal composites and RBCs, respectively, with derivatization efficiencies of ∼50%. Replicate precision indicated precision of ∼15% for RBC FAMEs above the detection limits and recoveries did not differ by treatment group or time.

### Statistics

Data normality was assessed using Shapiro-Wilkes tests and Q-Q plots of raw, log, and Johnson transformed data. Johnson transformations were the most robust and used for analyses unless indicated. Mean differences were assessed with Student’s t-test. Colinear dietary and body composition factors were clustered using an implementation of the VARCLUS algorithm. The influence of physiological parameters and habitual diet on the baseline OM3I were evaluated using linear mixed models alone and together. The minimum set of strong OM3I predictors were generated by stepwise linear regressions with the Bayesian information criterion (BIC) minimization as the stopping function. The influence of diet group and study week on RBC fatty acid composition was assessed using linear mixed models with participant as a random effect with group and week as main effects, followed by adjusted multiple comparison tests (Least Square Means Tukey HSD) if main or interaction effects were identified. Pearson’s correlations were used to assess correlations in change (wk8-wk0) after adjustement for baseline values. Participants in the DGAD group with the lowest and highest OM3I change were segregated into two magnitude response subgroups using hierarchical cluster analyses of the 0wk to 8wk change in the OM3I (Delta-OM3I_Wk0-8_). Participants were also segregated into OM3I rate of change response subgroups by diet. To this end, K means clustering, the fold-change in the Wk0 to Wk2 OM3I and the 8wk rate of change in the OM3I were used together. All statistical analyses were performed in JMP Pro v 15.1 (SAS Institute, Cary, NC).

## Results

### Participant numbers and characteristics

A complete diagram of the enrollment, allocation, follow-up and analysis is provided in **Figure 1**. Of the 52 participants enrolled, 44 completed the intervention without adverse effects. Summarized baseline characteristics of the participants included in the primary analysis are shown in **Table 1**, with participant level data provided in **Supplemental Table S1**. However, RBC fatty acid results were unavailable for one baseline and one terminal blood draw from 2 individuals in the TAD group due to poor analytical performance (n =2/132; i.e. 1.5%). Therefore, analyses for the primary outcome of intervention change in OM3I and secondary outcomes investigating the influence of body composition on the change in OM3I and the interaction between OM3I change and fasting triglyceridemia were performed with n =22 for the DGAD and n =20 for the TAD groups. In the DGAD group, a single individual showed inconsistancies between their pre-screening and baseline fasting triglyceride levels, and analyses were performed with and without this subject.

**Figure 1:**
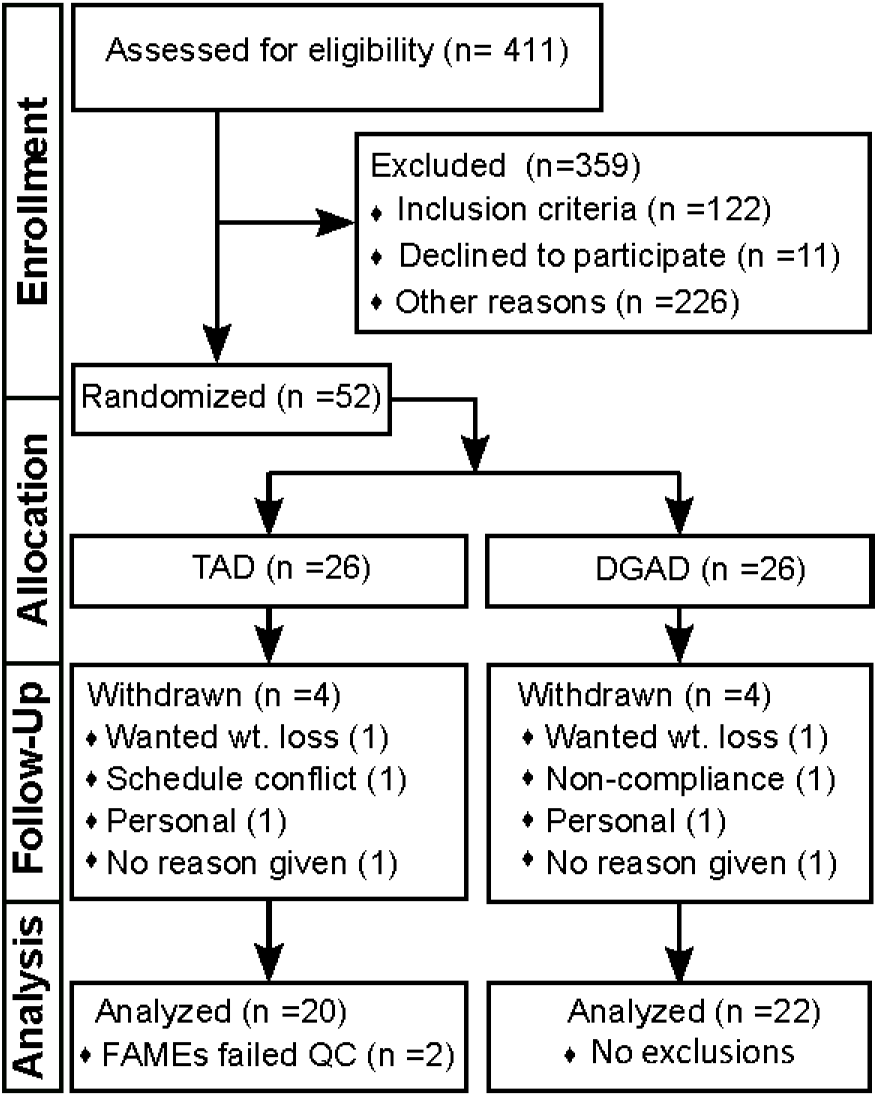
Study flow diagram documenting participant enrollement, allocation, follow-up and analysis. This diagram conforms the Consolidated Standards of Reporting Trials (i.e. CONSORT). TAD – typical American diet; DGAD - Dietary Guidelines for American’s diet; FAMEs – red blood cell fatty acid analysis; QC – quality control criterion.

**Table 1:** Study cohort baseline clinical and body composition characteristics ^1^.

| Variables | Units | TAD | DGAD |
| --- | --- | --- | --- |
| n |  | 20 | 22 |
| Age | y | 48.0 ± 10.0 | 50.7 ± 12.6 |
| OM3I | % | 5.98 ± 1.35 | 5.63 ± 1.27 |
| Fasting TGs | mg/dL | 111 ± 49 | 143 ± 101 |
| HOMA-IR |  | 3.77 ± 2.75 | 3.22 ± 2.43 |
| BMI | kg/m<sup>2</sup> | 33.0 ± 3.68 | 32.0 ± 4.0 |
| Body Weight | kg | 88.5 ± 13.8 | 88.6 ± 15.6 |
| Total Mass<sub>DEXA</sub><sup>2</sup> | kg | 87.0 ± 13.7 | 86.9 ± 15.3 |
| Lean Mass<sub>DEXA</sub><sup>2</sup> | kg | 47.1 ± 7.1 | 47.0 ± 8.0 |
| Fat Mass<sub>DEXA</sub><sup>2</sup> | kg | 38.0 ± 7.7 | 37.8 ± 7.8 |
| Android Fat Mass<sup>2</sup> | kg | 3.33 ± 0.99 | 3.48 ± 1.04 |
| Gynoid Fat Mass<sup>2</sup> | kg | 6.36 ± 1.45 | 6.37 ± 1.62 |
| Android:Gynoid<sup>2</sup> | % | 52.1 ± 0.11 | 56.0 ± 15.1 |
| % Body Fat<sup>2</sup> | % | 43.2 ± 3.9 | 43.3 ± 3.0 |
| % Trunk Fat<sup>2</sup> | % | 44.2 ± 5.3 | 45.3 ± 3.7 |

1- All values are means ± SD. Group mean differences were not detected by Student's t-tests ( $\alpha=0.05$ ). DEXA – dual-energy x-ray absorptiometry; DGAD – Dietary Guidelines for Americans diet; OM3I – omega-3 index; TAD – typical American diet; TG – triglycerides.
2- Reported values were derived from DEXA measurements

### Diet and red blood cell fatty acids profiles

The average fatty acid composition of both diet plans are shown in **Table 2**, while the corresponding average gram amounts delivered in the 8-day meal rotations are shown in **Table 3**. The individual participant daily intake of each fatty acid is provided in **Supplemental Table S2**. These analyses confirmed that the aggregate TAD contained higher saturated fatty acids (SFA) and monounsaturated fatty acids (MUFA) compared to the DGAD (**Figure 2A**). The TAD also had lower omega-3 polyunsaturated fatty acids (n3-PUFAs), with the relative abundance of n6-PUFAs being equivalent between the two diets. The average daily intake of EPA + DHA for the 8-day menu cycle was 0.18 ± 0.02 and 2.24 ± 0.33 g/d for the TAD and DGAD, respectively. However, as can be seen in **Figure 2B** in the DGAD meal plan, long-chain n3-PUFAs were provided solely in three independent meals on Day 1, Day 5, and Day 7 with the EPA + DHA levels of 3.0 ± 0.42, 2.5 ± 0.37, and 12 ± 1.7 g/d, or approximately 34 ± 3, 29 ± 3 and 133 ± 11 mg/kg body wt, respectively in menu day composites. Moreover, both meal plans contained a substantial amount of ALA. The foods providing the majority of omega-3 fatty acids are described in **Table 4**, and in more detail in **Supplemental Table 3**. The diet group average RBC fatty acid composition at baseline and after 2 and 8wk of dietary intervention are shown in **Table 5**.

**Table 2:**
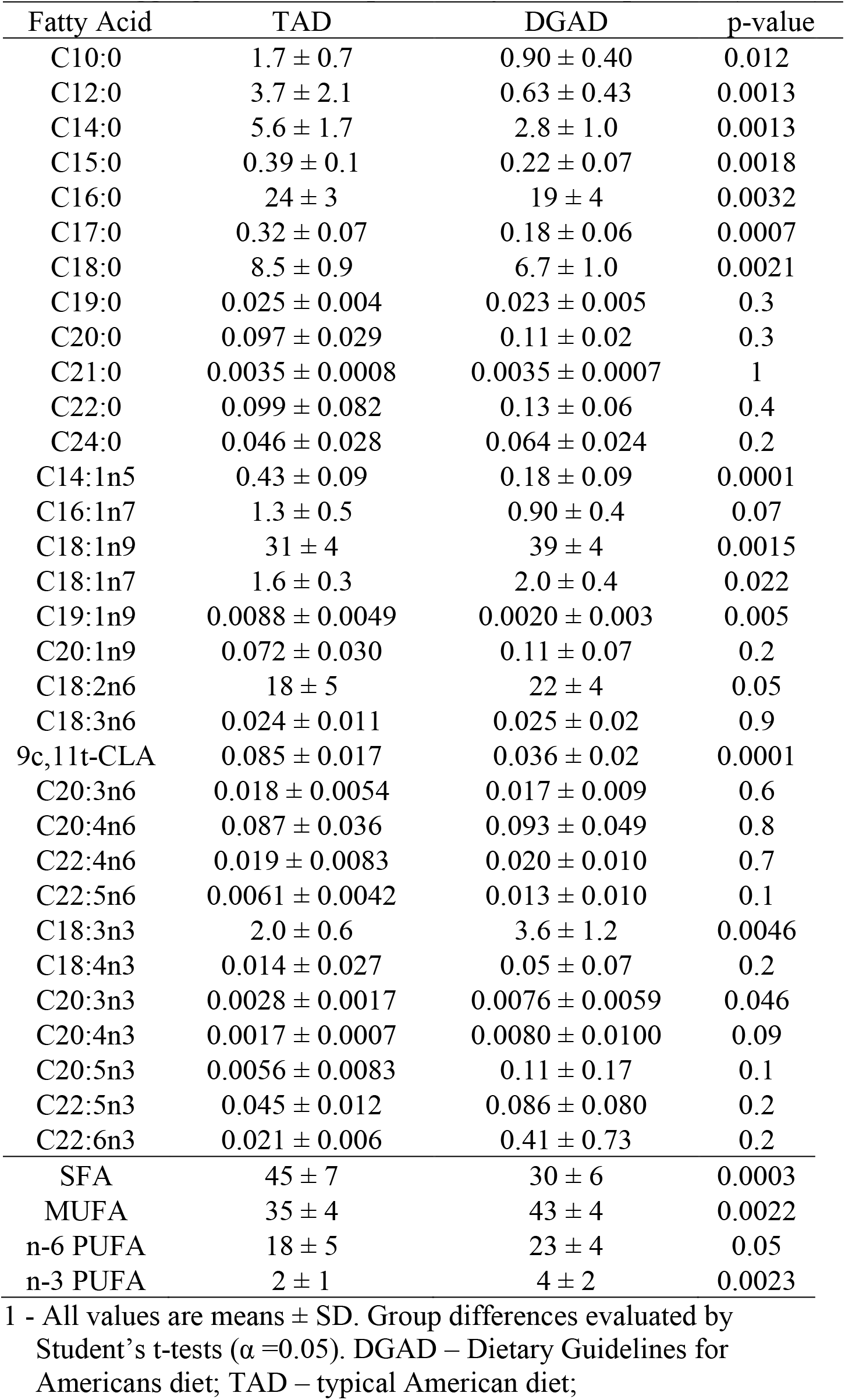
Aggregate meal composite fatty acid composition (mol%) ^1^.

| Fatty Acid | TAD | DGAD | p-value |
| --- | --- | --- | --- |
| C10:0 | 1.7 ± 0.7 | 0.90 ± 0.40 | 0.012 |
| C12:0 | 3.7 ± 2.1 | 0.63 ± 0.43 | 0.0013 |
| C14:0 | 5.6 ± 1.7 | 2.8 ± 1.0 | 0.0013 |
| C15:0 | 0.39 ± 0.1 | 0.22 ± 0.07 | 0.0018 |
| C16:0 | 24 ± 3 | 19 ± 4 | 0.0032 |
| C17:0 | 0.32 ± 0.07 | 0.18 ± 0.06 | 0.0007 |
| C18:0 | 8.5 ± 0.9 | 6.7 ± 1.0 | 0.0021 |
| C19:0 | 0.025 ± 0.004 | 0.023 ± 0.005 | 0.3 |
| C20:0 | 0.097 ± 0.029 | 0.11 ± 0.02 | 0.3 |
| C21:0 | 0.0035 ± 0.0008 | 0.0035 ± 0.0007 | 1 |
| C22:0 | 0.099 ± 0.082 | 0.13 ± 0.06 | 0.4 |
| C24:0 | 0.046 ± 0.028 | 0.064 ± 0.024 | 0.2 |
| C14:1n5 | 0.43 ± 0.09 | 0.18 ± 0.09 | 0.0001 |
| C16:1n7 | 1.3 ± 0.5 | 0.90 ± 0.4 | 0.07 |
| C18:1n9 | 31 ± 4 | 39 ± 4 | 0.0015 |
| C18:1n7 | 1.6 ± 0.3 | 2.0 ± 0.4 | 0.022 |
| C19:1n9 | 0.0088 ± 0.0049 | 0.0020 ± 0.003 | 0.005 |
| C20:1n9 | 0.072 ± 0.030 | 0.11 ± 0.07 | 0.2 |
| C18:2n6 | 18 ± 5 | 22 ± 4 | 0.05 |
| C18:3n6 | 0.024 ± 0.011 | 0.025 ± 0.02 | 0.9 |
| 9c,11t-CLA | 0.085 ± 0.017 | 0.036 ± 0.02 | 0.0001 |
| C20:3n6 | 0.018 ± 0.0054 | 0.017 ± 0.009 | 0.6 |
| C20:4n6 | 0.087 ± 0.036 | 0.093 ± 0.049 | 0.8 |
| C22:4n6 | 0.019 ± 0.0083 | 0.020 ± 0.010 | 0.7 |
| C22:5n6 | 0.0061 ± 0.0042 | 0.013 ± 0.010 | 0.1 |
| C18:3n3 | 2.0 ± 0.6 | 3.6 ± 1.2 | 0.0046 |
| C18:4n3 | 0.014 ± 0.027 | 0.05 ± 0.07 | 0.2 |
| C20:3n3 | 0.0028 ± 0.0017 | 0.0076 ± 0.0059 | 0.046 |
| C20:4n3 | 0.0017 ± 0.0007 | 0.0080 ± 0.0100 | 0.09 |
| C20:5n3 | 0.0056 ± 0.0083 | 0.11 ± 0.17 | 0.1 |
| C22:5n3 | 0.045 ± 0.012 | 0.086 ± 0.080 | 0.2 |
| C22:6n3 | 0.021 ± 0.006 | 0.41 ± 0.73 | 0.2 |
| SFA | 45 ± 7 | 30 ± 6 | 0.0003 |
| MUFA | 35 ± 4 | 43 ± 4 | 0.0022 |
| n-6 PUFA | 18 ± 5 | 23 ± 4 | 0.05 |
| n-3 PUFA | 2 ± 1 | 4 ± 2 | 0.0023 |

<sup>1</sup> - All values are means ± SD. Group differences evaluated by Student's t-tests ( $\alpha = 0.05$ ). DGAD – Dietary Guidelines for Americans diet; TAD – typical American diet;

**Table 3:**
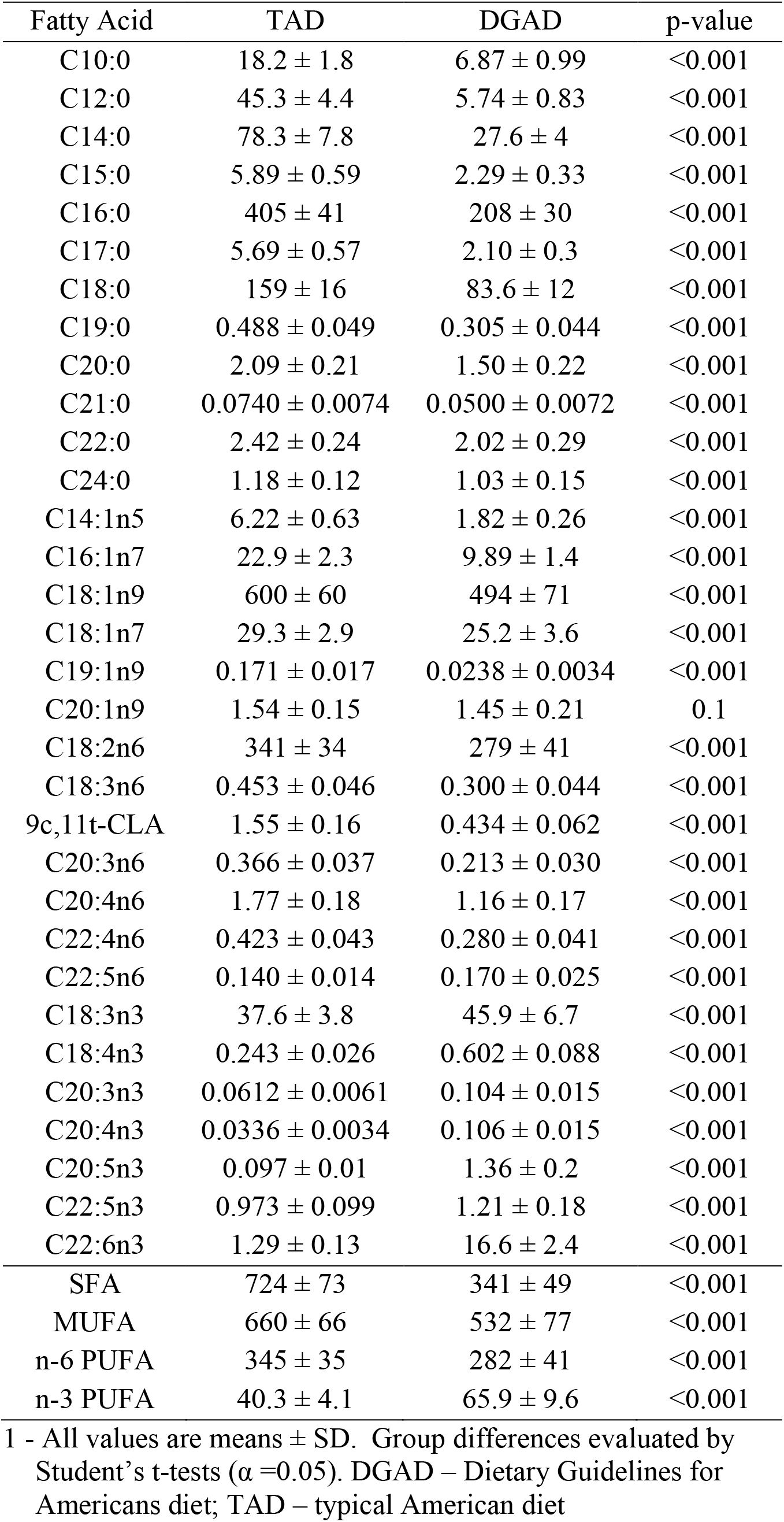
Average fatty acid intake (g/8-day rotation) ^1^.

| Fatty Acid | TAD | DGAD | p-value |
| --- | --- | --- | --- |
| C10:0 | 18.2 ± 1.8 | 6.87 ± 0.99 | <0.001 |
| C12:0 | 45.3 ± 4.4 | 5.74 ± 0.83 | <0.001 |
| C14:0 | 78.3 ± 7.8 | 27.6 ± 4 | <0.001 |
| C15:0 | 5.89 ± 0.59 | 2.29 ± 0.33 | <0.001 |
| C16:0 | 405 ± 41 | 208 ± 30 | <0.001 |
| C17:0 | 5.69 ± 0.57 | 2.10 ± 0.3 | <0.001 |
| C18:0 | 159 ± 16 | 83.6 ± 12 | <0.001 |
| C19:0 | 0.488 ± 0.049 | 0.305 ± 0.044 | <0.001 |
| C20:0 | 2.09 ± 0.21 | 1.50 ± 0.22 | <0.001 |
| C21:0 | 0.0740 ± 0.0074 | 0.0500 ± 0.0072 | <0.001 |
| C22:0 | 2.42 ± 0.24 | 2.02 ± 0.29 | <0.001 |
| C24:0 | 1.18 ± 0.12 | 1.03 ± 0.15 | <0.001 |
| C14:1n5 | 6.22 ± 0.63 | 1.82 ± 0.26 | <0.001 |
| C16:1n7 | 22.9 ± 2.3 | 9.89 ± 1.4 | <0.001 |
| C18:1n9 | 600 ± 60 | 494 ± 71 | <0.001 |
| C18:1n7 | 29.3 ± 2.9 | 25.2 ± 3.6 | <0.001 |
| C19:1n9 | 0.171 ± 0.017 | 0.0238 ± 0.0034 | <0.001 |
| C20:1n9 | 1.54 ± 0.15 | 1.45 ± 0.21 | 0.1 |
| C18:2n6 | 341 ± 34 | 279 ± 41 | <0.001 |
| C18:3n6 | 0.453 ± 0.046 | 0.300 ± 0.044 | <0.001 |
| 9c,11t-CLA | 1.55 ± 0.16 | 0.434 ± 0.062 | <0.001 |
| C20:3n6 | 0.366 ± 0.037 | 0.213 ± 0.030 | <0.001 |
| C20:4n6 | 1.77 ± 0.18 | 1.16 ± 0.17 | <0.001 |
| C22:4n6 | 0.423 ± 0.043 | 0.280 ± 0.041 | <0.001 |
| C22:5n6 | 0.140 ± 0.014 | 0.170 ± 0.025 | <0.001 |
| C18:3n3 | 37.6 ± 3.8 | 45.9 ± 6.7 | <0.001 |
| C18:4n3 | 0.243 ± 0.026 | 0.602 ± 0.088 | <0.001 |
| C20:3n3 | 0.0612 ± 0.0061 | 0.104 ± 0.015 | <0.001 |
| C20:4n3 | 0.0336 ± 0.0034 | 0.106 ± 0.015 | <0.001 |
| C20:5n3 | 0.097 ± 0.01 | 1.36 ± 0.2 | <0.001 |
| C22:5n3 | 0.973 ± 0.099 | 1.21 ± 0.18 | <0.001 |
| C22:6n3 | 1.29 ± 0.13 | 16.6 ± 2.4 | <0.001 |
| SFA | 724 ± 73 | 341 ± 49 | <0.001 |
| MUFA | 660 ± 66 | 532 ± 77 | <0.001 |
| n-6 PUFA | 345 ± 35 | 282 ± 41 | <0.001 |
| n-3 PUFA | 40.3 ± 4.1 | 65.9 ± 9.6 | <0.001 |

<sup>1</sup> - All values are means ± SD. Group differences evaluated by Student's t-tests ( $\alpha = 0.05$ ). DGAD – Dietary Guidelines for Americans diet; TAD – typical American diet

**Figure 2:**
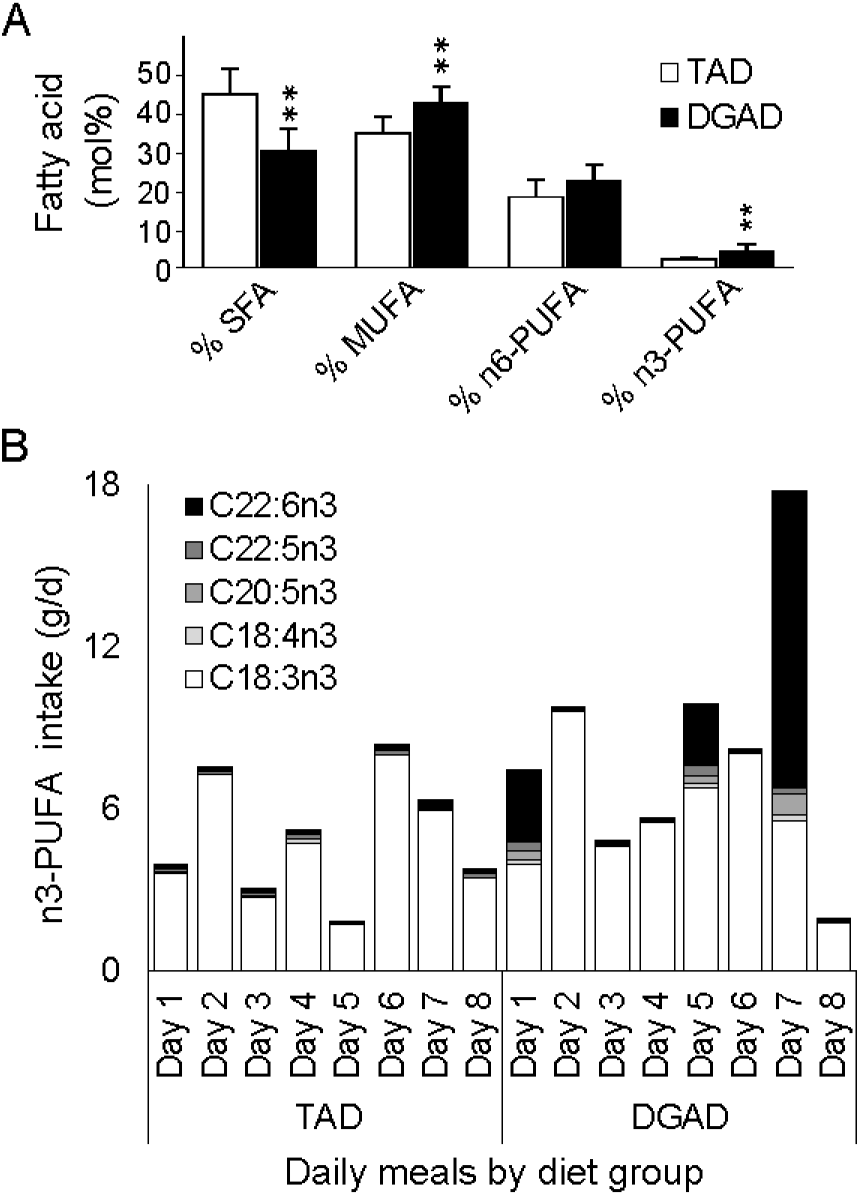
Fatty acid composition of TAD and DGAD menus. **A)** Aggregate meal composite fatty acid composition differences between the typical American diet (TAD) and Dietary Guidelines for American’s diet (DGAD) menus. Mean composition differences between meal plans are indicated at p <0.001 (**). **B)** Omega-3 PUFA fatty acid composition in individual meals of the 8-day rotating TAD and DGAD plans. Individual fatty acid residues are displayed if occurring at >0.1 mol% of any meal. Fatty acid mol% composition was determined in duplicate from a single set of meal composities by GC-MS analysis of fatty acid methyl esters.

**Table 4:**
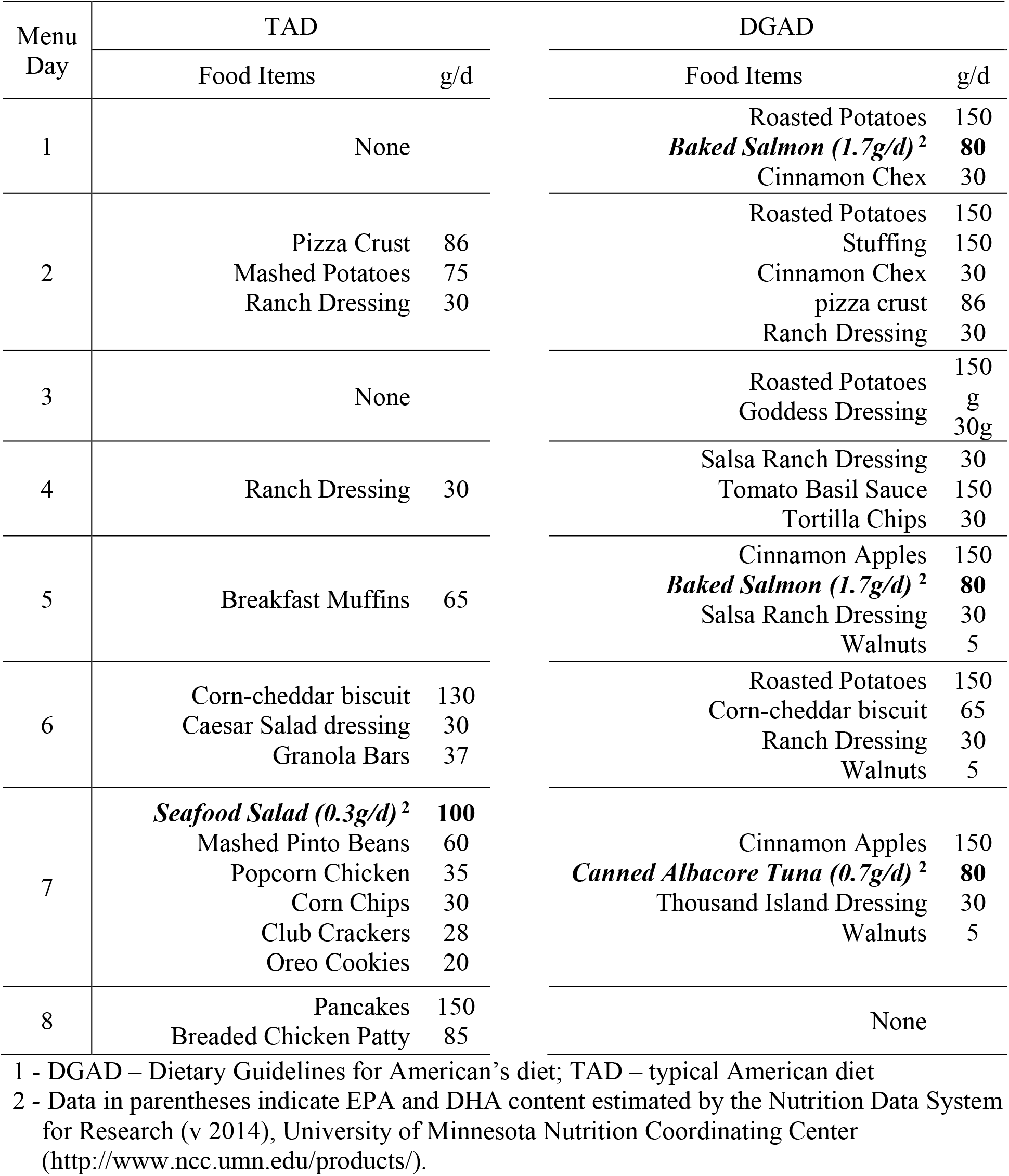
Amounts of foods providing >0.3g/d of dietary ALA, EPA and DHA^1^.

**Table 5:** RBC fatty acid mole percent composition ^1^.

| Fatty Acid | TAD (n =20) |  |  | DGAD (n =22) |  |  |
| --- | --- | --- | --- | --- | --- | --- |
|  | Wk 0 | Wk 2 | Wk 8 | Wk 0 | Wk 2 | Wk 8 |
| C14:0 | 1.28 ± 0.73 <sup>AB</sup> | 1.05 ± 1.00 <sup>B</sup> | 1.55 ± 1.20 <sup>A</sup> | 1.29 ± 0.93 | 1.11 ± 0.49 | 0.993 ± 0.66 |
| C15:0 | 0.387 ± 0.300 | 0.371 ± 0.074 | 0.288 ± 0.54 | 0.346 ± 0.130 | 0.319 ± 0.082 | 0.306 ± 0.39 |
| C16:0 | 29.5 ± 3.8 | 30.1 ± 3.3 | 29.8 ± 1.0 | 28.7 ± 4.1 | 30.0 ± 3.1 | 29.7 ± 3.3 |
| C17:0 | 0.401 ± 0.11 | 0.362 ± 0.12 | 0.449 ± 0.34 | 0.430 ± 0.110 | 0.356 ± 0.095 | 0.404 ± 0.15 |
| C18:0 | 24.2 ± 4.2 | 23.6 ± 4.1 | 23.7 ± 6.3 | 26.0 ± 5.4 | 24.9 ± 4.6 | 24.3 ± 5.2 |
| C18:1n9 | 10.9 ± 1.3 | 10.7 ± 1.1 | 10.6 ± 5.5 | 10.8 ± 1.7 | 10.7 ± 1.6 | 10.9 ± 1.7 |
| C18:1n7 | 2.07 ± 0.38 | 2.03 ± 0.26 | 1.92 ± 0.23 | 1.95 ± 0.65 | 2.02 ± 0.44 | 1.98 ± 0.43 |
| C18:2n6 | 6.52 ± 0.94 | 6.42 ± 0.90 | 6.36 ± 6.2 | 6.31 ± 0.92 | 5.94 ± 0.73 | 5.91 ± 0.95 |
| C20:3n6 | 1.25 ± 0.24 | 1.28 ± 0.35 | 1.33 ± 1.6 | 1.39 ± 0.46 | 1.42 ± 0.43 | 1.37 ± 0.19 |
| C20:4n6 | 9.55 ± 1.4 | 9.93 ± 1.50 | 10.1 ± 2.2 | 9.21 ± 1.3 | 9.29 ± 1.4 | 9.22 ± 1.4 |
| C22:4n6 | 4.06 ± 0.73 | 4.11 ± 0.83 | 4.28 ± 0.26 | 4.22 ± 0.88 | 4.20 ± 0.85 | 4.00 ± 0.88 |
| C22:5n6 | 0.834 ± 0.32 | 0.877 ± 0.28 | 0.841 ± 0.23 | 0.899 ± 0.27 <sup>ab</sup> | 0.945 ± 0.26 <sup>a</sup> | 0.727 ± 0.3 <sup>b</sup> |
| C20:5n3 | 0.430 ± 0.130 <sup>A</sup> | 0.381 ± 0.082 <sup>AB</sup> | 0.345 ± 0.64 <sup>B</sup> | 0.383 ± 0.15 <sup>b</sup> | 0.422 ± 0.13 <sup>b</sup> | 0.493 ± 0.18 <sup>a</sup> |
| C22:5n3 | 3.14 ± 0.73 | 3.20 ± 0.84 | 3.02 ± 1.20 | 2.90 ± 0.75 | 2.93 ± 0.59 | 2.89 ± 0.82 |
| C22:6n3 | 5.47 ± 1.1 | 5.57 ± 0.69 | 5.45 ± 0.46 | 5.24 ± 0.89 <sup>b</sup> | 5.47 ± 1.3 <sup>b</sup> | 6.86 ± 1.3 <sup>a</sup> |
| Σ SFA | 55.8 ± 3.5 | 55.5 ± 3.6 | 55.8 ± 9.5 | 56.7 ± 4.1 | 56.7 ± 4.1 | 55.7 ± 3.9 |
| Σ MUFA | 12.9 ± 1.6 | 12.7 ± 1.3 | 12.5 ± 1.6 | 12.7 ± 2.2 | 12.7 ± 2 | 12.8 ± 2 |
| Σ n-6 PUFA | 22.2 ± 2.3 | 22.6 ± 2.6 | 22.9 ± 1.2 | 22.0 ± 2.6 | 21.8 ± 2.2 | 21.2 ± 2.6 |
| Σ n-3 PUFA | 9.04 ± 1.4 | 9.15 ± 0.79 | 8.82 ± 1.8 | 8.52 ± 1.3 <sup>b</sup> | 8.83 ± 1.8 <sup>b</sup> | 10.3 ± 1.6 <sup>a</sup> |
| OM3-I | 5.90 ± 1.4 | 5.95 ± 1.2 | 5.8 ± 0.72 | 5.63 ± 1.3 <sup>b</sup> | 5.89 ± 0.95 <sup>b</sup> | 7.36 ± 1.4 <sup>a</sup> |

1- All values are means ± SD. Mean differences for each fatty acid between times within diet groups were assessed by ANOVAs controlling for participant as a random effect and a least squares mean difference Tukey HSD post-hoc tests. For each fatty acid residue, values within diet groups with different superscripts are different at $\alpha = 0.05$ . TAD – typical American diet; DGAD – Dietary Guidelines for Americans diet; OM3I – omega-3 index

### Habitual diet and physiological correlates with baseline OM3I

Combined analyses of FFQ in **Supplemental Table S4** and baseline body composition data in **Supplemental Table S1** using the entire set of completeing subjects (n = 44) identified the estimated dietary EPA + DHA:dietary saturated fat ratio (i.e. (EPA+DHA):SFA) and android fat mass as the strongest predictors of baseline OM3I (OM3I_(Wk0)_). Variable clustering of FFQ data showed the OM3I to be contained in a cluster with the dietary EPA + DHA, the (EPA+DHA):dietary saturated fat (SFAT) ratio, the (EPA+DHA):total dietary fat (TFAT) ratio, and the MUFA:TFAT ratio. Of these, the (EPA+DHA):SFAT ratio and (EPA+DHA):TFAT ratio were the most strongly correlated with the OM3I_(Wk0)_ (p <0.01) and FFQ estimated EPA + DHA intake was the weakest (p =0.06). Exploring the baseline OM3I correlations with body composition data showed android fat to be negatively correlated and the single best body composition predictor of this status marker (r^2^ =0.25, p =0.0007). Combining dietary and body composition factors into a stepwise linear regression analysis yielded an improved model explaining 43% of the variance in the baseline OM3I (r^2^ =0.43, p <0.0001), which included android fat mass (p =0.00012) and the (EPA+DHA):SFA ratio (p =0.0018) or (EPA+DHA):TFAT ratio (**Supplemental Figure 1**). The android fat mass and (EPA+DHA):SFA or (EPA+DHA):TFAT were not correlated (p =0.9).

### Intervention diet impact on RBC lipids and the OM3I

The fatty acid composition of RBCs was differentially altered by intervention diets. While no changes in SFAs or MUFAs were observed, a variety of changes in the PUFAs were noted. In terms of the amount of lipid per mg of RBC, DHA was increased by the DGAD (p =0.041) but not TAD (p =0.3). Concentrations of EPA were marginally increased (p =0.08) and decreased (p =0.1) by the DGAD and TAD, respectively. When expressed as a mole percent composition, the DGAD group showed increases in EPA and DHA (p <0.0001), and decreases in C22:5n6 (DPA; p =0.012), while the TAD showed a decreased EPA composition (p =0.013), reflected in a 20% reduction in the EPA/DHA relative abundance (p = 0.0019). The level of ALA in RBCs was near the analytical detection limit in the assay as applied, and the resulting data were not available for analysis. This is in agreement with reports of RBC ALA content of ∼0.1mol% [38].

While baseline OM3I of ∼5.6 did not differ by group (**Table1**), at study completion, the DGAD-group OM3I increased to 7.33 ± 1.36 (p<0.001), while the the TAD-group OM3I was unchanged at 5.8 ± 0.76 (p =0.6). As previously reported [17, 21], the baseline omega-3 status influenced the individual response to the intervention. As seen in **Figure 3**, after 2 weeks of feeding changes in the OM3I were driven by baseline associated interactions in both groups. However, after 8 weeks of feeding, a significant elevation in the OM3I is observed in the DGAD-group, with 21 of 22 (i.e. 95%) participants demonstrating an increase, as opposed to only 30% of the TAD-group.

**Figure 3:**
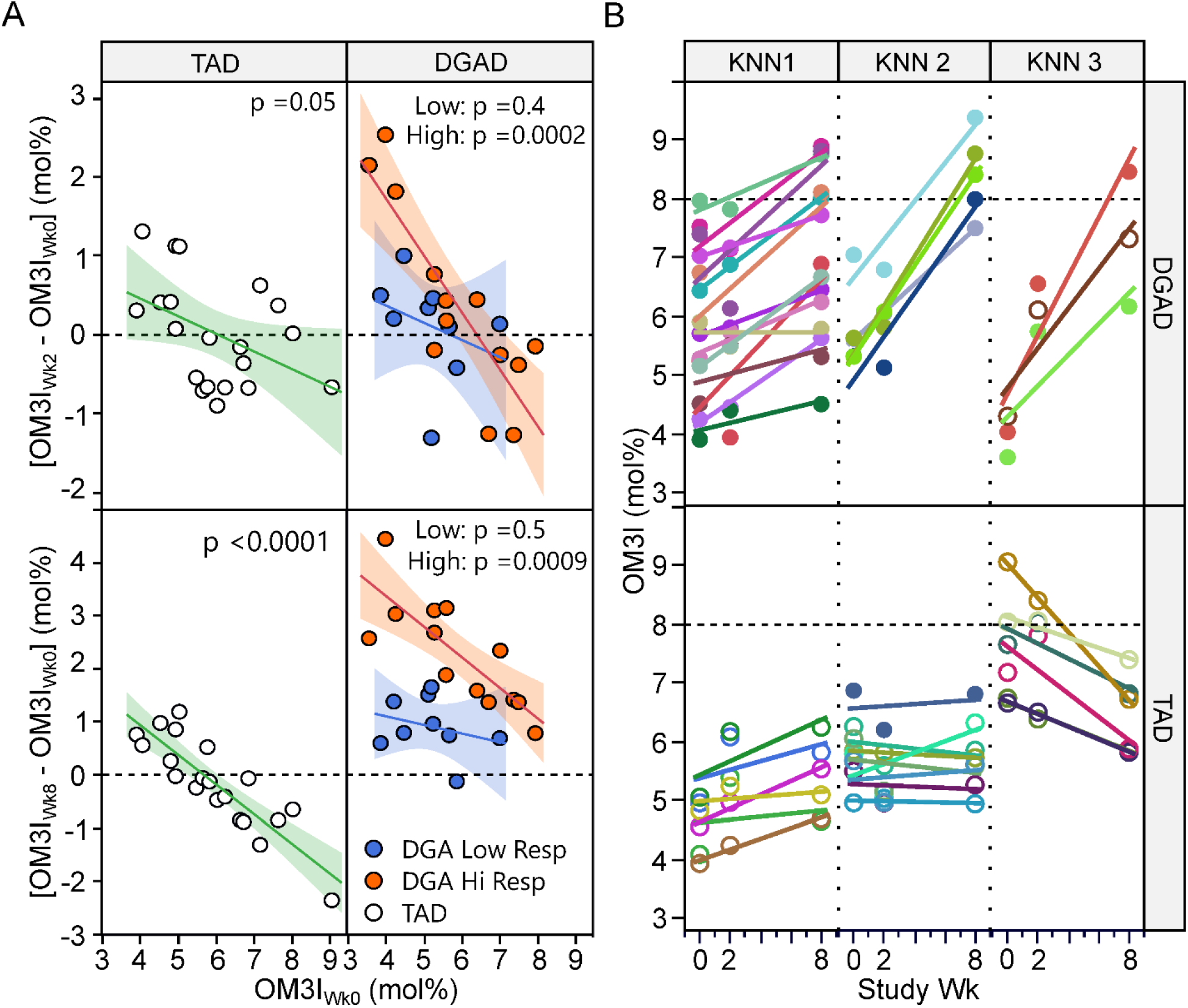
Omega-3 index response variance by diet group and time. A) Participants within the typical American diet (TAD) and Dietary Guidelines for American’s diet (DGAD) groups were clustered according to their baseline adjusted omega-3 index (OM3I) fold change at 8wks by hierarchical cluster analysis and projected on both 2wk (top) and 8wk (bottom) data. Identified DGAD subgroups are described as Low (blue) and High (orange) responding groups The correlations between basal status and magnitude of change were significant in the High, but not Low response groups. B) Participants within the TAD and DGAD groups were clustered based on their OM3I rate of change and basal status, by K-means clustering, with each diet group being segregated into 3 groups of participants with different responses. In the TAD group (n =20), the OM3I stabilized at 5.8 ± 0.7 across all groups, while participants in the DGAD group (n =22) generally increased after 8wks of feeding.

### OM3I response subgroup analysis

Despite equivalent dietary omega-3 intake in the DGAD, substantial variability in the response was observed that were not fully explained by the basal OM3I status. A cluster analysis of the baseline adjusted 8wk change in the OM3I segregated the DGAD-group into high and low responsive clusters, while the TAD-group remained as a single group (**Figure 3A**). A linear mixed model of the DGAD 8wk change in OM3I (Delta-OM3I_(Wk0-8)_) using OM3I_(Wk0)_ and the OM3I response group, revealed a significant interaction between OM3I_(Wk0)_ and the 8wk Delta-OM3I_(Wk0-8)_ (p =0.049), with the OM3I_(Wk0)_ having a weaker influence in the low reponders. The low responsive group also showed evidence of reduced insulin sensitivity as measured by HOMA-IR (p =0.045), with 8 of 9 low responsive individuals with HOMA-IR scores >2.1, as opposed to only 6 of 13 high responsive individuals. A K-means clustering of the rate of change in the OM3I between 0 and 8wk and the log of the fold-change between wk 0 and 2, identified 3 unique clusters for both the TAD and DGAD groups as shown in **Figure 3B**. In the DGAD-group, the OM3I increased 1.0 ± 0.5, 2.6 ± 0.5, and 3.4 ± 0.8 in clusters 1, 2, and 3, respectively. In the TAD-group, the average OM3I increased by 0.8 ± 0.3 in cluster 1, remained unchanged (i.e. -0.1 ± 0.3) in cluster 2, and decreased by 1.4 ± 0.6 in cluster 3.

### Determinants of OM3I change

It has previously been reported that baseline OM3I, body mass, age, sex, and physical activity are strong determinants of the OM3I response to supplementation [12]. Considering the TAD- and DGAD-groups together (n =42), the baseline OM3I alone explained 34% of the OM3I response (**Supplemental Figure 2**). However as intervention exposures were significantly different, an analysis by diet was performed. In the TAD group 74% of the variance in the Delta-OM3I_(Wk0-8)_ was explained by negative correlations with the baseline OM3I alone (p <0.0001). In the DGA group only 30% of the variance was explained by the OM3I_(Wk0)_. Models including OM3I_(Wk0)_ and the average EPA + DHA daily intake were improved (r^2^ = 0.56, RMSE = 0.16, p =0.0004), however, the Delta-OM3I_(Wk0-8)_ showed a counterintuitive negative correlation with intake (i.e. adjusting for basal status, as dose increased change decreased). A stepwise linear regression built from DEXA-based body composition, BMI and age data accounted for 70% of the variance in the DGAD Delta-OM3I_(Wk0-8)_. This false discovery rate adjusted standard least squares regression model (r^2^ = 0.70, RMSE =0.12; p =0.002) included negative associations with the OM3I_(Wk0)_ (p_adj_ =0.027), trunk % fat (p_adj_ =0.012), lean body mass (p_adj_ =0.013) and positive associations with android fat mass (p_adj_ =0.027) and BMI (p_adj_ =0.033). As seen in **Figure 4**, the omega-3 fatty acid and body variables condense into four correlated variable clusters. Of these factors, android fat mass is positively correlated with lean body mass, BMI and trunk % fat (p <0.001), along with the average EPA+DHA daily intake. Notably, multidimensional outliers in highly correlated independent variables were not observed. Exploring model factor interactions revealed OM3I_(Wk0)_ interactions with android fat mass (p =0.014), lean mass (p = 0.020) and BMI (p =0.025). Modeling the dose dependent change (i.e. [Delta-OM3I_(Wk0-8)_/(EPA+DHA)] and including the OM3I_(Wk0)_ by android fat interaction improved the model performance (r^2^ =0.85; p <0.0001, RMSE = 0.011; **Supplemental Figure 3**). A boosted tree analysis of the Delta-OM3I_(Wk0-8)_, also showed the OM3I_(Wk0)_, along with % body fat, android % fat, and the android:gynoid fat ratio as important explanatory variables (data not shown). These analyses suggest that after adjusting for the basal OM3I status, the omega-3 fatty acid response to dietary EPA+DHA intake is lower as body size increases, but higher as android fat deposition increases.

**Figure 4:**
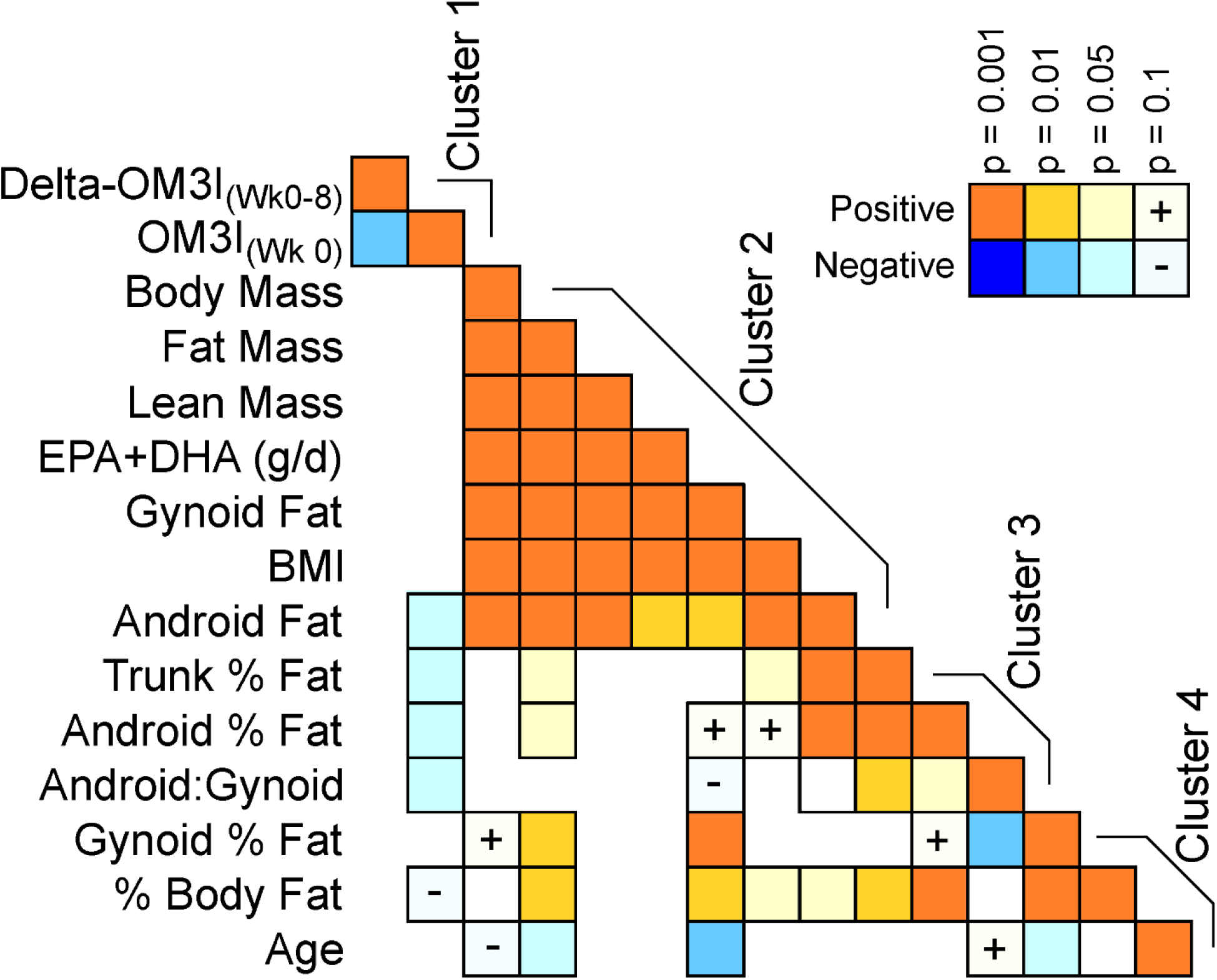
Pearson’s correaltion heatmap of the DGAD-group 8wk change in omega-3 status, basal omega-3 statis and DEXA-dependent body composition measures. Correlation statistics are calculated on normally transformed data, with positive (orange) and negative (blue) correlation strength indicated by color intensity. Variables were clustered in Jmp v16 using implementation of the SAS Varclus algorithm. Delta-OM3I_(Wk0-8)_ – 8wk change in OM3I; EPA+DHA – estimated daily intake; OM3I_(Wk0)_ – basal omega 3 index.

### Triglyceride response to OM3I changes

Omega 3 fatty acid intake has been reported to lower circulating triglycerides. The current data set was evaluated for evidence of triglyceride interactions with OM3I status. Changes in triglycerides themselves were inversely related to their baseline levels, and evenly distributed between increases and decreases regardless of the diet intervention group, suggesting a substantial regression to the mean effect with respect to the measured changes in fasting triglycerides (data not shown). Fasting triglyceride levels were not directly correlated with the OM3I. In the DGAD-group, changes in the TGs, adjusted for baseline TGs, were positively correlated with changes in the OM3I, adjusted for baseline OM3I (r^2^ =0.27, p =0.007). However, removal of a single fasting TG measure, a statistical outlier at baseline in poor agreement with its study pre-screening TG concentration, reduced the strength of this correlation (r^2^ =0.16, p =0.08), although it remained stronger than that seen in the TAD-group (r^2^ =0.02, p =0.6). Analysis of DGAD OM3I response subgroups also showed no association between the OM3I and fasting TGs.

## Discussion

Increasing long-chain omega-3 fatty acid consumption is associated with reductions in lipidomic cardiovascular disease risk factors [39]. The omega-3 index, defined as the sum of the EPA and DHA mole percent relative to the total fatty acid content in RBCs is an established and valuable omega-3 fatty acid status marker [11, 16, 18]. An OM3I between 8-12% appears protective, while a value <4% is associated with increased CVD risk [11, 18]. The present study was established to determine if a diet based on the 2010 DGA can beneficially alter the OM3I and/or blood TG concentrations when compared to a TAD, in pre-and postmenopausal women, within 8 wks. A secondary analysis was performed to evaluate the influence of body composition factors on the responsiveness of the OM3I to dietary omega-3 fatty acid intake.

It has been established that the baseline OM3I is negatively associated with the OM3I response to omega-3 fatty acid intake, and various factors have been reported to influence this status marker. For instance, FFQ-based estimates of the omega-3 fatty acid intake correlate with the OM3I, and an increase in weekly additional servings of fish has been associated with a 6-13% increase in the OM3I [40-43]. Fasting triglyceride levels, age, a history of high cholesterol, and smoking have also been reported to influence the OM3I [21-23, 43]. In the current study, the Block FFQ estimated EPA + DHA intake was poorly correlated with the basal OM3I unless expressed as a percentage of the total dietary fat, or its relative abundance with the saturated fat intake. Exploring associations between body composition and the OM3I, android fat mass was found to have a strong negative association with the baseline OM3I, which was independent of the dietary factors. Contradicting this finding, in a previous study of older women, a high dietary omega-3/omega-6 ratio was associated with higher android fat [44]. That study also found that a high dietary SFA/PUFA ratio was positively associated with android fat, a relationship that was weakly observed in the current data (p=0.1). Regardless, elevations in android fat deposition are reportedly associated with elevated plasma triglycerides, endothelial dysfunction, type 2 diabetes, and fatty liver disease [45-48], all conditions beneficially affected by high omega-3 fatty acid intake.

As demonstrated by the diet composition analysis, following the DGA recommendations significantly alters the dietary fat balance relative to a TAD, with elevations in n3-PUFAs and reductions in saturated fats. The DGAD provided ∼2.25g/d of EPA+DHA primarily in 3 meals of the 8-day rotating menu. Meals on these days included servings of 80g of salmon or albacore tuna in multiple forms including seafood pasta, a corn and salmon chowder, a salmon dinner, and a seafood salad. In contrast, the TAD provided ∼0.25g/d of EPA+DHA. A more detailed description of the meals and there composition has been previously reported [33]. Ultimately, in this sample of overweight/obese women with a mean basal OM3I of ∼5.6%, 8 weeks of the DGAD increased the OM3I by 1.7 ± 1.1%, while the TAD only reduced it by -0.15 ± 0.86%, with a final OM3I of 7.4 ± 1.35% and 5.8 ± 0.72%, respectively. While most individuals on the DGAD showed OM3I improvements of ∼1% over the course of the intervention, one person was unchanged, and 7 participants had 2.3 – 4.5% change in the OM3I, with 9 exceeding the recommended 8% OM3I by the end of the intervention. Therefore, it is possible to achieve a putatively beneficial OM3I status in 8 weeks by adhering to DGAD recommendations with significant EPA and DHA sources in only 3 meals per week. However, there is significant interindividual variability in response, and rates of improvement slow as the OM3I increases, consistent with previous findings [21, 31]. In mixed models adjusting for the basal OM3I effect, changes in the OM3I were lower as the percent trunk fat and lean body mass increased, but higher as absolute android fat mass and BMI increased. Therefore, it appears that in women with increased abdominal and overall adiposity the omega-3 intervention efficacy was enhanced after adjusting for basal status and body size and android adiposity. However, it is also important to note that women with a low OM3I intervention response also had reduced insulin sensitivity. Considering that men with a higher OM3I were found to have higher insulin sensitivity [49], these findings bring into question the cause-effect relationship between these factors. In contrast, after the 8wk TAD intervention, the average OM3I of individuals either increased or decreased toward an average OM3I of ∼5.8%, again with some variability influence primarily by the basal omega-3 fatty acid status. It is unclear if this maintenance of a modest OM3I was due to the low level of EPA+DHA in the diet, or associated with ALA-dependent biosynthesis of EPA. Had this change been purely associated with ALA conversion, an increase in EPA relative to DHA in the TAD group might be expected [26-28]. However, the EPA:DHA ratio was actually reduced in the TAD group at 8wks, suggesting that dietary DHA and/or it biosynthesis in this diet group were sufficient to maintain the OM3I in a neutral range with respect to cardiovascular disease risk.

Dietary long chain omega-3 fatty acids can have strong hypolipidemic effects in both normal and hyperlipidemic individuals, with these lipid-lowering effects being both dose and time-dependent [9, 39, 50]. For instance, in an 8wk intervention comparing 2 g/d of daily supplements and 2 meals/wk of 250g of trout, both treatments reduced TGs in hyperlipidemic participants [22]. In another effort, 0.85 g/d and 3.4g/d doses of EPA+DHA were compared in a population with a baseline OM3I of 4.46 ± 1.13%. After 8wks of treatment, the OM3I increased to 6.49 ± 0.21% and 8.79 ± 0.21%, for the low and high doses, respectively. However, only the high dose resulted in TG lowering in the moderately hypertriglyceridemic cohort. Similarly, a weak non-linear association between an increase in the OM3I and a decrease in fasting TG levels was reported in dyslipidemic patients receiving 1.67 g/d of EPA+DHA as fish oil capsules for 6mo, with individuals having a change in the OM3I of >4% showing the greatest TG reduction [51]. In the present study of women with a basal OM3I of 5.6 ± 1.3% and normal to mild hypertriglyceridemia, only weak associations between increasing OM3I and reduced fasting TGs were observed at 8wk in the DGAD group after outlier removal. Therefore, it cannot be concluded from the current data that habitual consumption of a DGAD with elevated ALA and intermittent exposure to substantial dietary EPA + DHA would reduce fasting TG in subjects if maintained for a longer duration.

## Limitations

While foods were purchased in lots to minimize variance in nutritional exposures, measured fatty acids in meal composites were generated at a single time point during the course of the study and the exact dietary exposures may differ from the measured composite meals. Thus omega-3 intake could have varied between participants over the course of the study. The current study has a relatively small sample size and includes individuals with a range of normal to mild fasting triglyceridemia, limiting the power to detect hypotriglyceridemic effects. This fact is exacerbated in the reported subgroup analysis. Moreover, genetic factors influencing both long chain omega-3 fatty acid biosynthesis and triglyceridemic responsiveness were not evaluated.

## Conclusion

Adherence to a DGAD with ∼16g of EPA+DHA provided in 3 meals/wk for 8wks improves the OM3I status in pre- and post-menopausal women with a basal OM3I of 5.6 ± 1.3%, while an ALA-rich TAD providing 1.4g/wk of EPA+DHA maintained an OM3I of ∼6%, which is in the neutral range with respect to CVD risk. While a cardioprotective OM3I of >8% was achieved in 40% of participants, no concurrent decreases in fasting TGs were observed in the current timeframe. Notably in women with greater abdominal and overall adiposity the omega-3 intervention efficacy was higher after adjusting for basal omega-3 fatty acid status and body size.

## Supporting information

Consort Checklist

## Data Availability

All data are included as supplement in PDF form. All data will be available upon publication. Early requests for a more accessible data will be considered by the senior authors.

## Abbreviations

ALA: alpha-linolenic acid
FFQs: Block Food Frequency questionnaires
CVD: cardiovascular disease
Delta-OM3I_(Wk0-8)_: 8wk omega 3 index change from baseline
DGA: Dietary Guidelines for Americans
GC-MS: gas chromatography-mass spectrometry
DEXA: dual-energy x-ray absorptiometry
FAME: fatty acid methyl esters
OM3I: omega-3 index
OM3I_(Wk0)_: baseline omega 3 index
RBC: red blood cell
SFAT: dietary saturated fat
TAD: typical American diet
TFAT: total dietary fat
TG: triglycerides

## Acknowledgements

This study was supported by the National Dairy Council, Campbell Soup Co., and USDA-ARS Projects 2032-51530-022-00D, 2032-51530-025-00D and 6026-51000-010-05S. The USDA is an equal opportunity provider and employer. The National Dairy Council and Campbell Soup Company did not participate in the final design, implementation, analysis, or interpretation of the data from this study. Campbell Soup provided some bulk foods and associated nutrient information. None of the authors had a conflict of interest to report. We thank our colleagues Sean H. Adams, Lindsay H. Allen, Kevin D. Laugero, and Charles B. Stephenson for their efforts in the study design and execution of the larger study in which the current experiments were nested; Dustin Burnett for menu design and production management; Leslie Woodhouse for managing the generation of clinical data; Janet Peerson for guidance and review of statistical analyses. The authors’ responsibilities were as follows **-** JWN and NLK: conceived and developed the research plan; JWN, SK, IJG, and NLK: conducted the research; JWN and IJG: analyzed the data and performed the statistical analysis; CER, JWN: wrote the primary manuscript; SK and NLK: provided significant editorial input of manuscript; all authors: have primary responsibility for final content and read and approved the final manuscript.

