## Supplemental Figures 1-3 for "An 8 week randomized Dietary Guidelines for Americans -based diet intervention improves the omega-3 index of healthy women"

---

**Description and Index of contained information**: This file three supplemental figures describing detailed statistical analyses associated with this manuscript as listed below.

**Supplemental Figure 1:** Linear regression analysis of study cohort baseline OM3I as a function of Android fat and estimated dietary EPA+DHA intake.

**Supplemental Figure 2**: Correlation of the log of the 8 week change in the omega 3 index (Log[Delta-OM3I(8wk)]) and baseline OM3I ($OM3I_{(Wk0)}$) in the entire study cohort (n=42).

**Supplemental Figure 3:** Linear regression analysis of the DGAD-group 8 wk change in the OM3I corrected for the ingested dose, as a function of the baseline OM3I and body composition factors.

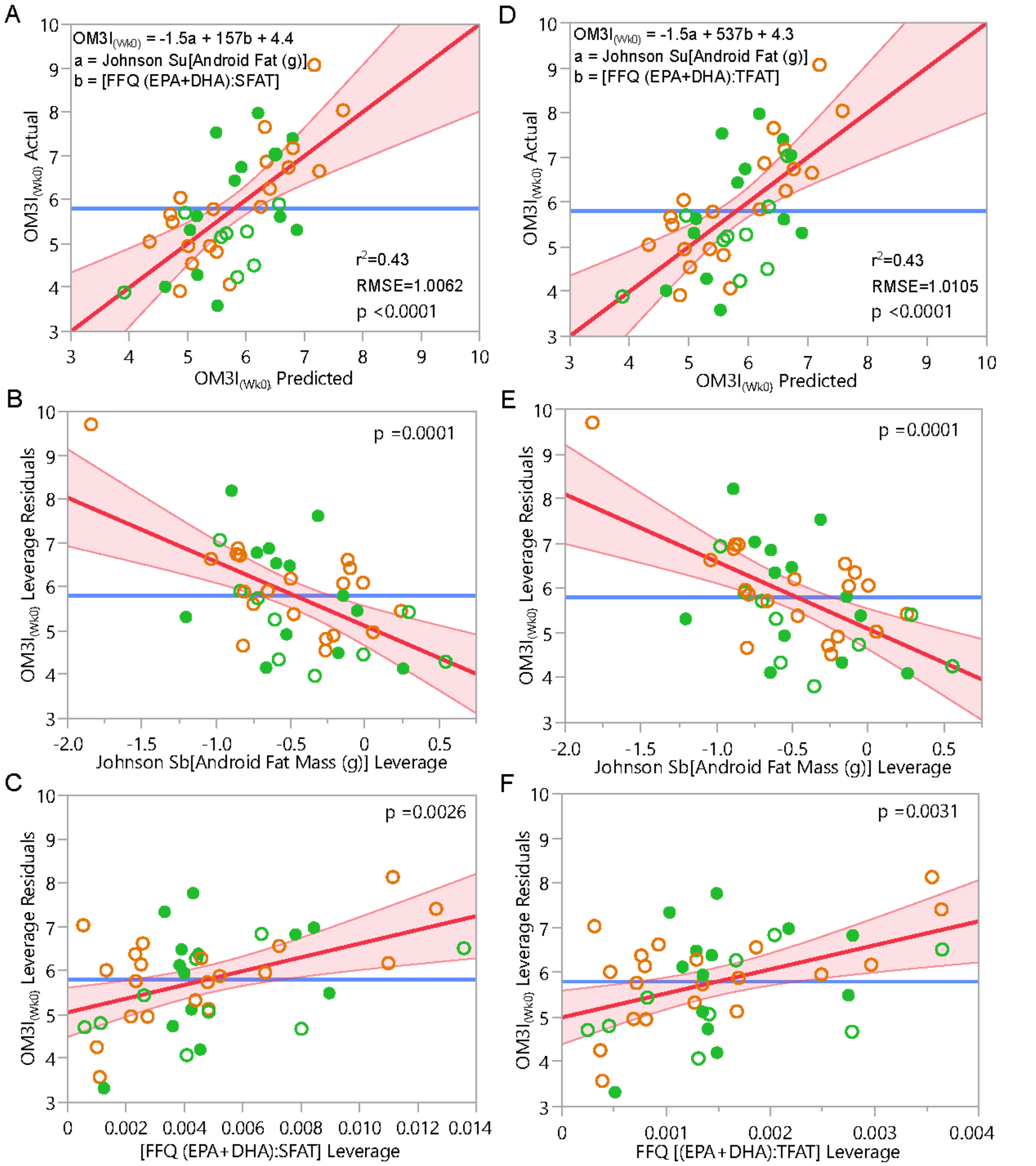


**Supplemental Figure 1: Linear regression analysis of study cohort baseline OM3I as a function of Android fat and estimated dietary EPA+DHA intake.** The relative abundancy of EPA + DHA to dietary saturated fat (SFAT) and total fat (TFAT) are stronger predictors than the absolute estimated intake. A) Actual by predicted plot showing results of the whole model with SFAT. B) Leverage plot of normalized Android fat adjusted for estimated (EPA+DHA) to SFAT ratio. C) Leverage plot of estimated (EPA+DHA) to SFAT ratio adjusted normalized Android fat. D) Actual by predicted plot showing results of the whole model with SFAT. E) Leverage plot of normalized Android fat adjusted for estimated (EPA+DHA) to TFAT ratio. F) Leverage plot of estimated (EPA+DHA) to TFAT ratio adjusted normalized Android fat. DGAD – green; TAD – orange; closed circle – High Response

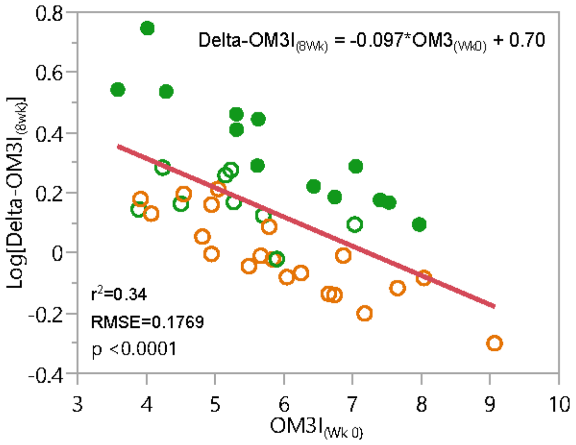


**Supplemental Figure 2: Correlation of the log of the 8 week change in the omega 3 index (Log[Delta-OM3I(8wk)]) and baseline OM3I (OM3I$_{(Wk0)}$) in the entire study cohort (n=42).** The basal OM3I is a strong predictor of the intervention associated change in the OM3I. DGAD – green; TAD – orange; closed circle – High Response

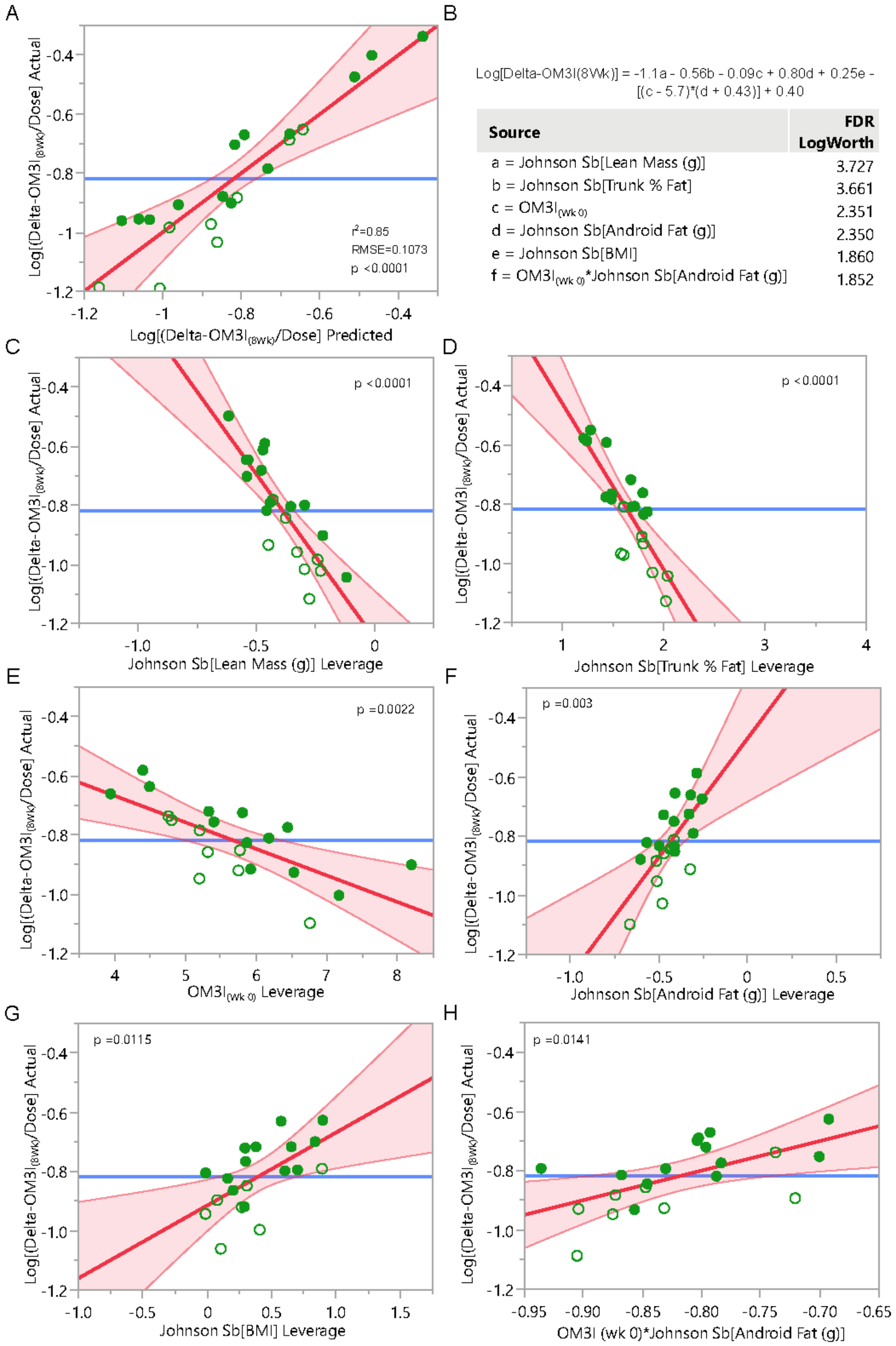


**Supplemental Figure 3: Linear regression analysis of the DGAD-group 8 wk change in the OM3I corrected for the ingested dose, as a function of the baseline OM3I and body composition factors.** A) Actual by predicted plot showing results of the whole model and B-H) leverage plots of each adjusted factor. open circles – Low Responders; closed circle – High Responsders
