## Supplemental Table 1 for "An 8 week randomized Dietary Guidelines for Americans -based diet intervention improves the omega-3 index of healthy women"

**Supplemental Table S1: Participant clinical, DEXA and RBC FAME data**

| Subject_ID | Exclusion Comments | Diet Group | Diet Week | Baseline Adj OM3 Change Cluster | 5y Age Range | Sex | Final BMI | BMI Category | HOMA-IR | IR Status | HbA1c (%) | Fasting TG (mg/dL) | Fasting Triglyceridemia | DiastolicPressure | SystolicPressure | Sistolic:Diastolic | Weight (kg) | Total Mass (g) | Total Fat Mass (g) | Total Lean Mass (g) | Total Lean + BMC (g) | Trunk % Fat | Total % Body Fat | Android Fat Mass (g) | Gynoid Fat Mass (g) | Android % Fat | Gynoid % Fat |
|---|---|---|---|---|---|---|---|---|---|---|---|---|---|---|---|---|---|---|---|---|---|---|---|---|---|---|---|
| 3 | | TAD | 0 | TAD | 26-30 | Female | 33.9 | Obese | 13.17 | IR | 6.15 | 55 | Normal | 73.5 | 126 | 1.71 | 109.4 | 107138.4 | 47095.5 | 57437.7 | 60042.9 | 47.7 | 44 | 4612 | 8602 | 53 | 48.7 |
| 9 | | DGAD | 0 | DGAD Low Resp | 21-25 | Female | 28.5 | Overweight | 2.05 | IR | 5.12 | 74 | Normal | 63.5 | 119 | 1.87 | 77.85 | 76351.2 | 35374.3 | 39148.3 | 40976.9 | 43.9 | 46.3 | 2601 | 6867 | 46 | 50.2 |
| 20 | | TAD | 0 | TAD | 41-45 | Female | 33.3 | Obese | 1.32 | no-IR | 5.54 | 207 | Mild-Moderate | 71.5 | 110.5 | 1.55 | 81.4 | 80784.2 | 34679.9 | 43783 | 46104.3 | 41.6 | 42.9 | 2631 | 7109 | 41 | 46.8 |
| 32 | | DGAD | 0 | DGAD Low Resp | 36-40 | Female | 37.7 | Obese | 3.15 | IR | 5.02 | 348 | Mild-Moderate | 78 | 122 | 1.56 | 114.4 | 112297.5 | 51066.3 | 58502.7 | 61231.2 | 48.8 | 45.5 | 5252 | 9403 | 52 | 47.9 |
| 51 | Missing terminal blood draw | TAD | 0 | TAD | 46-50 | Female | 37.7 | Obese | 3.24 | IR | 5.56 | 260 | Mild-Moderate | 73.5 | 110.5 | 1.50 | 95.8 | 94509.5 | 44474.3 | 47466.1 | 50035.2 | 46 | 47.1 | 3468 | 8457 | 46.9 | 52.8 |
| 52 | | TAD | 0 | TAD | 36-40 | Female | 32.1 | Obese | 5.41 | IR | 4.9 | 165 | Mild-Moderate | 80 | 117 | 1.46 | 82.9 | 81453.1 | 33786.5 | 45874.8 | 47666.5 | 44.3 | 41.5 | 2890 | 4633 | 47.1 | 42.2 |
| 53 | | TAD | 0 | TAD | 41-45 | Female | 35.6 | Obese | 2.55 | IR | 5.53 | 84 | Normal | 71.5 | 127 | 1.78 | 88.15 | 87156 | 44668.1 | 40607.4 | 42487.9 | 53.8 | 51.3 | 3948 | 7842 | 54.2 | 53 |
| 69 | | TAD | 0 | TAD | 46-50 | Female | 31.1 | Obese | 3.69 | IR | 5.21 | 121 | Normal | 74 | 121 | 1.64 | 77.3 | 76169 | 34235.3 | 40049.6 | 41933.7 | 47.3 | 44.9 | 3792 | 5863 | 51.9 | 46.8 |
| 72 | | DGAD | 0 | DGAD Hi Resp | 41-45 | Female | 29.9 | Overweight | 1.44 | no-IR | 4.88 | 202 | Mild-Moderate | 74.5 | 127 | 1.70 | 84.75 | 83167.1 | 35456 | 45349.5 | 47711.1 | 45.6 | 42.6 | 3232 | 6833 | 47.5 | 47.7 |
| 79 | | DGAD | 0 | DGAD Hi Resp | 26-30 | Female | 33.5 | Obese | 1.64 | no-IR | 5.23 | 36 | Normal | 58 | 112.5 | 1.94 | 94.1 | 92282.1 | 43000.7 | 46218 | 49281.4 | 47.8 | 46.6 | 3669 | 7463 | 51.6 | 47.8 |
| 84 | | DGAD | 0 | DGAD Hi Resp | 46-50 | Female | 33.9 | Obese | 1.39 | no-IR | 5.11 | 200 | Mild-Moderate | 72 | 125.5 | 1.74 | 98.8 | 95819.2 | 41078.4 | 52293.9 | 54740.8 | 45.8 | 42.9 | 4096 | 7088 | 48 | 44.9 |
| 85 | | TAD | 0 | TAD | 46-50 | Female | 34.8 | Obese | 0.62 | no-IR | 5.27 | 76 | Normal | 72.5 | 119.5 | 1.65 | 83.15 | 81987.2 | 35830.8 | 43672.2 | 46156.5 | 44.3 | 43.7 | 3322 | 7181 | 46.6 | 48.1 |
| 87 | | TAD | 0 | TAD | 51-55 | Female | 31 | Obese | 1.92 | no-IR | 5.23 | 127 | Normal | 63.5 | 107.5 | 1.69 | 82.55 | 81106.1 | 28411.5 | 50149.9 | 52694.7 | 37.6 | 35 | 2606 | 4517 | 40.9 | 34.9 |
| 96 | | DGAD | 0 | DGAD Hi Resp | 61-65 | Female | 28 | Overweight | 3.43 | IR | 5.66 | 126 | Normal | 68.5 | 127 | 1.85 | 70.2 | 68910 | 28414.9 | 38763 | 40495.1 | 43.6 | 41.2 | 2463 | 4824 | 44.5 | 42.8 |
| 97 | | DGAD | 0 | DGAD Hi Resp | 56-60 | Female | 27.5 | Overweight | 0.45 | no-IR | 5.69 | 151 | Mild-Moderate | 74 | 123 | 1.66 | 59.65 | 58717.7 | 22945.8 | 34439.2 | 35771.9 | 42.8 | 39.1 | 1949 | 3177 | 42.8 | 36.7 |
| 102 | | DGAD | 0 | DGAD Hi Resp | 61-65 | Female | 25.3 | Overweight | 2.18 | IR | 5.16 | 121 | Normal | 65.5 | 128.5 | 1.96 | 69.95 | 68450 | 26600.1 | 39799.1 | 41850 | 43.2 | 38.9 | 2725 | 4252 | 46.3 | 40.3 |
| 107 | | DGAD | 0 | DGAD Hi Resp | 56-60 | Female | 32 | Obese | 1.76 | no-IR | 5.5 | 59 | Normal | 72 | 126 | 1.75 | 92.3 | 90823 | 36850.6 | 51568 | 53972.4 | 42.3 | 40.6 | 2941 | 6208 | 42.3 | 42.9 |
| 109 | | TAD | 0 | TAD | 51-55 | Female | 27.7 | Overweight | 4.11 | IR | 5.36 | 59 | Normal | 69.5 | 116 | 1.67 | 64.15 | 63375.4 | 29835.7 | 31826.5 | 33539.7 | 48.3 | 47.1 | 2205 | 4936 | 48 | 49.5 |
| 112 | | TAD | 0 | TAD | 26-30 | Female | 33.7 | Obese | 2.54 | IR | 5.19 | 143 | Normal | 68 | 111.5 | 1.64 | 84.75 | 82637 | 34873.1 | 45674.4 | 47763.9 | 43.3 | 42.2 | 3275 | 6362 | 46.4 | 44.7 |
| 113 | | DGAD | 0 | DGAD Hi Resp | 51-55 | Female | 34.2 | Obese | 3.08 | IR | 5.38 | 29 | Normal | 72.5 | 139 | 1.92 | 105.9 | 104060.3 | 41413.1 | 59996.2 | 62647.2 | 39 | 39.8 | 3125 | 6607 | 40.3 | 40.9 |
| 135 | | TAD | 0 | TAD | 36-40 | Female | 31.8 | Obese | 2.56 | IR | 4.96 | 121 | Normal | 72 | 120 | 1.67 | 97.55 | 96637.2 | 40317.1 | 54211.9 | 56320.1 | 44.1 | 41.7 | 4134 | 5703 | 46 | 38.8 |
| 137 | | TAD | 0 | TAD | 56-60 | Female | 31.9 | Obese | 4.81 | IR | 5.48 | 116 | Normal | 88 | 135 | 1.53 | 100.2 | 97242.6 | 42203.7 | 52599.4 | 55038.9 | 45.5 | 43.4 | 3841 | 5637 | 45.7 | 40.7 |
| 139 | | DGAD | 0 | DGAD Hi Resp | 56-60 | Female | 29.5 | Overweight | 3.07 | IR | 5.27 | 93 | Normal | 82 | 148 | 1.80 | 72.55 | 71338.3 | 28772 | 40867 | 42566.3 | 41.8 | 40.3 | 2935 | 4337 | 45.7 | 39.6 |
| 144 | | TAD | 0 | TAD | 46-50 | Female | 37 | Obese | 3.07 | IR | 5.58 | 148 | Normal | 56.5 | 109 | 1.93 | 115.2 | 113580.7 | 50325.5 | 60037.4 | 63255.2 | 44.2 | 44.3 | 4063 | 8463 | 45.1 | 47.5 |
| 151 | Missing baseline RBC data | TAD | 0 | TAD | 41-45 | Female | 29.5 | Overweight | | IR | 5.75 | | | 77 | 109 | 1.42 | 86.65 | 85687.8 | 35928.2 | 47368.1 | 49759.6 | 43.7 | 41.9 | 2891 | 5675 | 44.9 | 45 |
| 154 | | DGAD | 0 | DGAD Low Resp | 61-65 | Female | 35.5 | Obese | 3.78 | IR | 5.49 | 130 | Normal | 82.5 | 141 | 1.71 | 93.8 | 92081.4 | 44553.1 | 45447.8 | 47528.3 | 46.9 | 48.4 | 3010 | 7542 | 44.7 | 48.4 |
| 168 | | TAD | 0 | TAD | 46-50 | Female | 26.8 | Overweight | 0.78 | no-IR | 5.2 | 75 | Normal | 79.5 | 132.5 | 1.67 | 72.25 | 70099.3 | 24673.7 | 43270.7 | 45425.6 | 28.4 | 35.2 | 1209 | 4338 | 28.6 | 39.2 |
| 170 | | TAD | 0 | TAD | 51-55 | Female | 37 | Obese | 3.96 | IR | 5.64 | 67 | Normal | 72 | 120 | 1.67 | 98.6 | 96801.5 | 48335 | 46244.1 | 48466.4 | 51.4 | 49.9 | 4237 | 8253 | 52.3 | 50.9 |
| 172 | | DGAD | 0 | DGAD Hi Resp | 46-50 | Female | 32.2 | Obese | 1.15 | no-IR | 4.93 | 52 | Normal | 53 | 107 | 2.02 | 92.65 | 91296.6 | 40424.3 | 48551.1 | 50872.3 | 41.4 | 44.3 | 2992 | 8092 | 45.1 | 51 |
| 175 | | TAD | 0 | TAD | 51-55 | Female | 27.9 | Overweight | 3.67 | IR | 6.1 | 33 | Normal | 75 | 131 | 1.75 | 81.55 | 80299.6 | 31304.3 | 46825.8 | 48995.3 | 42.1 | 39 | 2649 | 5479 | 42.8 | 41.9 |
| 180 | | TAD | 0 | TAD | 61-65 | Female | 30.6 | Obese | 1.05 | no-IR | 5.44 | 145 | Normal | 68 | 114.5 | 1.68 | 81.6 | 80201.3 | 33562.8 | 44773.3 | 46638.5 | 39.1 | 41.8 | 2483 | 5680 | 40.8 | 43.4 |
| 182 | | DGAD | 0 | DGAD Low Resp | 26-30 | Female | 33.9 | Obese | 9.39 | IR | 5.59 | 60 | Normal | 74.5 | 111.5 | 1.50 | 108.5 | 106283.8 | 45635.1 | 58033 | 60648.7 | 46.7 | 42.9 | 4297 | 8371 | 49.8 | 48.3 |
| 183 | | TAD | 0 | TAD | 56-60 | Female | 40.2 | Obese | 4.00 | IR | 5.39 | 157 | Mild-Moderate | 70 | 99.5 | 1.42 | 108.45 | 107520.4 | 48611.6 | 56788.4 | 58908.8 | 45.1 | 45.2 | 5174 | 8105 | 50 | 47.3 |
| 186 | | TAD | 0 | TAD | 51-55 | Female | 39.1 | Obese | 4.02 | IR | 5.72 | 83 | Normal | 85 | 142.5 | 1.68 | 104.55 | 102843.7 | 46633.4 | 53986.8 | 56210.3 | 47.1 | 45.3 | 4470 | 8081 | 49.9 | 47.4 |
| 189 | | DGAD | 0 | DGAD Low Resp | 61-65 | Female | 30.7 | Obese | 3.49 | IR | 5.63 | 142 | Normal | 86 | 171.5 | 1.99 | 77.45 | 75180 | 33188.2 | 40120.4 | 41991.8 | 47.3 | 44.1 | 3075 | 4829 | 51 | 45 |
| 194 | | DGAD | 0 | DGAD Low Resp | 61-65 | Female | 27.2 | Overweight | 0.88 | no-IR | 5.34 | 156 | Mild-Moderate | 77.5 | 129 | 1.66 | 69.75 | 68912.5 | 32188.7 | 35257.7 | 36723.7 | 52.6 | 46.7 | 3567 | 5325 | 52.7 | 46.4 |
| 198 | screening and Visit 1 TG mismatch | DGAD | 0 | DGAD Hi Resp | 51-55 | Female | 38.7 | Obese | 5.41 | IR | 5.48 | 426 | Mild-Moderate | 68 | 125 | 1.84 | 104.1 | 101912 | 48033.1 | 51561.6 | 53878.9 | 49.5 | 47.1 | 5184 | 9016 | 54.5 | 50.6 |
| 232 | | TAD | 0 | TAD | 61-65 | Female | 29.5 | Overweight | 6.05 | IR | 5.27 | 193 | Mild-Moderate | 66 | 116 | 1.76 | 73.6 | 71933.6 | 30681.7 | 39618.9 | 41251.9 | 44.6 | 42.7 | 2465 | 4934 | 45.5 | 42.9 |
| 234 | | DGAD | 0 | DGAD Low Resp | 56-60 | Female | 35.9 | Obese | 5.65 | IR | 5.82 | 259 | Mild-Moderate | 80 | 139.5 | 1.74 | 98.7 | 96795.5 | 43663.6 | 51362 | 53131.9 | 51.2 | 45.1 | 6047 | 5507 | 54.3 | 38.3 |
| 239 | | TAD | 0 | TAD | 51-55 | Female | 30 | Obese | 6.24 | IR | 5.84 | 52 | Normal | 78.5 | 128.5 | 1.64 | 82.7 | 81006.8 | 34530.6 | 43842.1 | 46476.2 | 43.9 | 42.6 | 2525 | 5576 | 43.1 | 43.2 |
| 243 | | DGAD | 0 | DGAD Low Resp | 56-60 | Female | 26.3 | Overweight | 9.49 | IR | 5.47 | 201 | Mild-Moderate | 62 | 103.5 | 1.67 | 75.75 | 73978.1 | 27909.8 | 43977.2 | 46068.3 | 38.1 | 37.7 | 2313 | 4944 | 37 | 40 |
| 244 | | DGAD | 0 | DGAD Hi Resp | 36-40 | Female | 34.7 | Obese | 1.14 | no-IR | 5.46 | 67 | Normal | 73 | 121 | 1.66 | 101.25 | 99835.9 | 44504 | 52730.6 | 55331.8 | 47 | 44.6 | 4017 | 6568 | 48.6 | 43.1 |
| 245 | | DGAD | 0 | DGAD Hi Resp | 61-65 | Female | 38.5 | Obese | 3.41 | IR | 6.04 | 64 | Normal | 86.5 | 136 | 1.57 | 108.8 | 106963.5 | 45163 | 59225.8 | 61800.6 | 44.1 | 42.2 | 4337 | 7133 | 43.9 | 43.6 |
| 258 | | DGAD | 0 | DGAD Low Resp | 56-60 | Female | 29.4 | Overweight | 3.53 | IR | 5.59 | 152 | Mild-Moderate | 74 | 123.5 | 1.67 | 78.1 | 76690.2 | 34898.9 | 39715.8 | 41791.2 | 46.8 | 45.5 | 2823 | 5741 | 44.5 | 47.3 |
| 3 | | TAD | 2 | TAD | 26-30 | Female | 33.4 | Obese | 9.59 | IR | 6.22 | 84 | Normal | 71 | 116 | 1.63 | 107.73 | | | | | | | | | | |
| 9 | | DGAD | 2 | DGAD Low Resp | 21-25 | Female | 28.4 | Overweight | 1.44 | no-IR | 5.12 | 104 | Normal | 67 | 113 | 1.69 | 77.57 | | | | | | | | | | |
| 20 | | TAD | 2 | TAD | 41-45 | Female | 32.8 | Obese | 1.26 | no-IR | 5.5 | 122 | Normal | 70 | 106.5 | 1.52 | 80.23 | | | | | | | | | | |
| 32 | | DGAD | 2 | DGAD Low Resp | 36-40 | Female | 37.9 | Obese | 3.45 | IR | 4.95 | 325 | Mild-Moderate | 73 | 131 | 1.79 | 115.03 | | | | | | | | | | |
| 51 | Missing terminal blood draw | TAD | 2 | TAD | 46-50 | Female | 37.4 | Obese | 3.53 | IR | 5.83 | 202 | Mild-Moderate | 72 | 121.5 | 1.69 | 95.00 | | | | | | | | | | |
| 52 | | TAD | 2 | TAD | 36-40 | Female | 32.3 | Obese | 4.37 | IR | 4.92 | 185 | Mild-Moderate | 76 | 111.5 | 1.47 | 83.53 | | | | | | | | | | |
| 53 | | TAD | 2 | TAD | 41-45 | Female | 35.3 | Obese | 3.96 | IR | 5.73 | 88 | Normal | 76 | 123 | 1.62 | 87.37 | | | | | | | | | | |
| 69 | | TAD | 2 | TAD | 46-50 | Female | 31.2 | Obese | 3.35 | IR | 5.15 | 89 | Normal | 68.5 | 118 | 1.72 | 77.53 | | | | | | | | | | |
| 72 | | DGAD | 2 | DGAD Hi Resp | 41-45 | Female | 29.8 | Overweight | 2.56 | IR | 4.88 | 221 | Mild-Moderate | 69.5 | 122 | 1.76 | 84.55 | | | | | | | | | | |
| 79 | | DGAD | 2 | DGAD Hi Resp | 26-30 | Female | 33.2 | Obese | 2.08 | IR | 5.06 | 51 | Normal | 68.5 | 129.5 | 1.89 | 93.27 | | | | | | | | | | |
| 84 | | DGAD | 2 | DGAD Hi Resp | 46-50 | Female | 33.7 | Obese | 2.08 | IR | 4.89 | 162 | Mild-Moderate | 66 | 120.5 | 1.83 | 98.30 | | | | | | | | | | |
| 85 | | TAD | 2 | TAD | 46-50 | Female | 34.2 | Obese | 1.53 | no-IR | 5.18 | 101 | Normal | 76.5 | 133.5 | 1.75 | 81.85 | | | | | | | | | | |
| 87 | | TAD | 2 | TAD | 51-55 | Female | 30.9 | Obese | 1.26 | no-IR | | 97 | Normal | 71.5 | 111.5 | 1.56 | 82.35 | | | | | | | | | | |
| 96 | | DGAD | 2 | DGAD Hi Resp | 61-65 | Female | 27.3 | Overweight | 2.45 | IR | 5.65 | 113 | Normal | 70.5 | 108.5 | 1.54 | 68.63 | | | | | | | | | | |
| 97 | | DGAD | 2 | DGAD Hi Resp | 56-60 | Female | 27 | Overweight | 3.25 | IR | 5.54 | 139 | Normal | 69 | 113 | 1.64 | 58.43 | | | | | | | | | | |
| 102 | | DGAD | 2 | DGAD Hi Resp | 61-65 | Female | 25 | Overweight | 1.44 | no-IR | 5.35 | 88 | Normal | 70.5 | 127.5 | 1.81 | 69.1 | | | | | | | | | | |
| 107 | | DGAD | 2 | DGAD Hi Resp | 56-60 | Female | 31.7 | Obese | 3.85 | IR | 5.54 | 47 | Normal | 60 | 111 | 1.85 | 91.65 | | | | | | | | | | |
| 109 | | TAD | 2 | TAD | 51-55 | Female | 27.4 | Overweight | 3.04 | IR | 5.4 | 78 | Normal | 66.5 | 111 | 1.67 | 63.5 | | | | | | | | | | |
| 112 | | TAD | 2 | TAD | 26-30 | Female | 33.6 | Obese | 2.71 | IR | 5.2 | 159 | Mild-Moderate | 70 | 115 | 1.64 | 84.65 | | | | | | | | | | |
| 113 | | DGAD | 2 | DGAD Hi Resp | 51-55 | Female | 34.3 | Obese | 2.37 | IR | 5.31 | 61 | Normal | 72.5 | 122 | 1.68 | 105.95 | | | | | | | | | | |
| 135 | | TAD | 2 | TAD | 36-40 | Female | 31.5 | Obese | 4.09 | IR | 4.93 | 84 | Normal | 69 | 127 | 1.84 | 96.75 | | | | | | | | | | |
| 137 | | TAD | 2 | TAD | 56-60 | Female | 31.6 | Obese | 9.04 | IR | 5.5 | 170 | Mild-Moderate | 75.5 | 120 | 1.59 | 99.3 | | | | | | | | | | |
| 139 | | DGAD | 2 | DGAD Hi Resp | 56-60 | Female | 29.5 | Overweight | 3.22 | IR | 5.4 | 65 | Normal | 79 | 134 | 1.70 | 72.55 | | | | | | | | | | |
| 144 | | TAD | 2 | TAD | 46-50 | Female | 37.4 | Obese | 2.43 | IR | 5.21 | 126 | Normal | 68 | 130.5 | 1.92 | 116.25 | | | | | | | | | | |
| 151 | Missing baseline blood draw | TAD | 2 | TAD | 41-45 | Female | 29.7 | Overweight | 11.74 | IR | 5.87 | 86 | Normal | 64.5 | 122.5 | 1.90 | 87.35 | | | | | | | | | | |
| 154 | | DGAD | 2 | DGAD Low Resp | 61-65 | Female | 35.2 | Obese | 3.26 | IR | 5.64 | 121 | Normal | 89 | 130.5 | 1.47 | 92.95 | | | | | | | | | | |
| 168 | | TAD | 2 | TAD | 46-50 | Female | 26.2 | Overweight | 0.84 | no-IR | 4.99 | 88 | Normal | 80.5 | 137.5 | 1.71 | 70.6 | | | | | | | | | | |
| 170 | | TAD | 2 | TAD | 51-55 | Female | 37.1 | Obese | 4.44 | IR | 5.69 | 60 | Normal | 73 | 122 | 1.67 | 98.9 | | | | | | | | | | |
| 172 | | DGAD | 2 | DGAD Hi Resp | 46-50 | Female | 32.4 | Obese | 2.12 | IR | 5.06 | 81 | Normal | 61 | 112 | 1.84 | 93.2 | | | | | | | | | | |
| 175 | | TAD | 2 | TAD | 51-55 | Female | 27.8 | Overweight | 1.57 | no-IR | 5.95 | 66 | Normal | 64 | 129.5 | 2.02 | 81.25 | | | | | | | | | | |
| 180 | | TAD | 2 | TAD | 61-65 | Female | 30.5 | Obese | 2.19 | IR | 5.36 | 205 | Mild-Moderate | 73.5 | 125 | 1.70 | 81.3 | | | | | | | | | | |
| 182 | | DGAD | 2 | DGAD Low Resp | 26-30 | Female | 33.8 | Obese | 9.26 | IR | 5.59 | 75 | Normal | 67 | 116.5 | 1.74 | 108.35 | | | | | | | | | | |
| 183 | | TAD | 2 | TAD | 56-60 | Female | 39.8 | Obese | 4.26 | IR | 5.23 | 192 | Mild-Moderate | 67 | 107.5 | 1.60 | 107.47 | | | | | | | | | | |
| 186 | | TAD | 2 | TAD | 51-55 | Female | 39.1 | Obese | 3.10 | IR | 5.66 | 80 | Normal | 71.5 | 116.5 | 1.63 | 104.5 | | | | | | | | | | |
| 189 | | DGAD | 2 | DGAD Low Resp | 61-65 | Female | 30.6 | Obese | 2.34 | IR | 5.67 | 133 | Normal | 81.5 | 136 | 1.67 | 77.25 | | | | | | | | | | |
| 194 | | DGAD | 2 | DGAD Low Resp | 61-65 | Female | 27.1 | Overweight | 1.43 | no-IR | 5.32 | 172 | Mild-Moderate | 86 | 141 | 1.64 | 69.45 | | | | | | | | | | |
| 198 | screening and Visit 1 TG mismatch | DGAD | 2 | DGAD Hi Resp | 51-55 | Female | 38.3 | Obese | 11.32 | IR | 5.3 | 205 | Mild-Moderate | 70.5 | 120 | 1.70 | 103.1 | | | | | | | | | | |
| 232 | | TAD | 2 | TAD | 61-65 | Female | 29.7 | Overweight | 2.90 | IR | 5.38 | 326 | Mild-Moderate | 63.5 | 118.5 | 1.87 | 74 | | | | | | | | | | |
| 234 | | DGAD | 2 | DGAD Low Resp | 56-60 | Female | 36 | Obese | 6.28 | IR | 5.81 | 318 | Mild-Moderate | 77 | 120 | 1.56 | 98.9 | | | | | | | | | | |
| 239 | | TAD | 2 | TAD | 51-55 | Female | 29.6 | Overweight | 6.24 | IR | 5.74 | 62 | Normal | 71.5 | 111 | 1.55 | 81.55 | | | | | | | | | | |
| 243 | | DGAD | 2 | DGAD Low Resp | 56-60 | Female | 26.4 | Overweight | 7.18 | IR | 5.51 | 144 | Normal | 71.5 | 105 | 1.47 | 76.1 | | | | | | | | | | |
| 244 | | DGAD | 2 | DGAD Hi Resp | 36-40 | Female | 34.6 | Obese | 1.45 | no-IR | 5.34 | 61 | Normal | 74.5 | 128 | 1.72 | 100.95 | | | | | | | | | | |
| 245 | | DGAD | 2 | DGAD Hi Resp | 61-65 | Female | 38.2 | Obese | 1.55 | no-IR | 6 | 57 | Normal | 72 | 122 | 1.69 | 107.95 | | | | | | | | | | |
| 258 | | DGAD | 2 | DGAD Low Resp | 56-60 | Female | 29.1 | Overweight | 3.88 | IR | 5.65 | 112 | Normal | 61 | 110.5 | 1.81 | 77.25 | | | | | | | | | | |
| 3 | | TAD | 8 | TAD | 26-30 | Female | 32.6 | Obese | 5.15 | IR | 5.9 | 83 | Normal | 83 | 112 | 1.35 | 105.03 | 102540.3 | 45457.4 | 54413.2 | 57083 | 48.2 | 44.3 | 4318 | 8534 | 51.4 | 49.4 |
| 9 | | DGAD | 8 | DGAD Low Resp | 21-25 | Female | 27.6 | Overweight | 1.83 | no-IR | 4.86 | 128 | Normal | 52 | 99.5 | 1.91 | 75.47 | 74849.9 | 34235.3 | 38767.1 | 40614.6 | 42 | 45.7 | 2505 | 6617 | 45.1 | 50.7 |
| 20 | | TAD | 8 | TAD | 41-45 | Female | 32.5 | Obese | 1.10 | no-IR | 5.29 | 134 | Normal | 61 | 107.5 | 1.76 | 79.43 | 78604 | 35849.6 | 40468 | 42754.4 | 44.8 | 45.6 | 2755 | 7574 | 46 | 50.2 |
| 32 | | DGAD | 8 | DGAD Low Resp | 36-40 | Female | 37.1 | Obese | 2.27 | IR | 4.93 | 343 | Mild-Moderate | 69 | 116.5 | 1.69 | 112.57 | 111065.9 | 53839 | 54463.7 | 57226.9 | 52.1 | 48.5 | 5527 | 9707 | 56.9 | 50.7 |
| 51 | Missing terminal RBC data | TAD | 8 | TAD | 46-50 | Female | 36.9 | Obese | 3.06 | IR | 5.5 | 246 | Mild-Moderate | 73.5 | 123 | 1.67 | 93.70 | 92456.3 | 42260.9 | 47631.6 | 50195.4 | 43.9 | 45.7 | 3265 | 8159 | 45.9 | 52.2 |
| 52 | | TAD | 8 | TAD | 36-40 | Female | 32.3 | Obese | 7.13 | IR | 4.93 | 186 | Mild-Moderate | 73 | 108 | 1.48 | 83.30 | 82078.5 | 33462.2 | 46826.2 | 48616.3 | 44.6 | 40.8 | 2843 | 4506 | 47.4 | 40.4 |
| 53 | | TAD | 8 | TAD | 41-45 | Female | 34.6 | Obese | 3.04 | IR | 5.59 | 72 | Normal | 82 | 116 | 1.41 | 85.70 | 84197.4 | 38385.5 | 43910.9 | 45811.8 | 47.8 | 45.6 | 3322 | 6648 | 48.6 | 46.6 |
| 69 | | TAD | 8 | TAD | 46-50 | Female | 30.8 | Obese | 2.78 | IR | 5.18 | 101 | Normal | 76 | 124.5 | 1.64 | 76.37 | 75415.8 | 34303.3 | 39218.7 | 41112.6 | 48.2 | 45.5 | 3827 | 5813 | 52.4 | 47 |
| 72 | | DGAD | 8 | DGAD Hi Resp | 41-45 | Female | 29.3 | Overweight | 2.12 | IR | 4.96 | 244 | Mild-Moderate | 76.5 | 127.5 | 1.67 | 83.10 | 82488.8 | 34544 | 45606 | 47944.7 | 45.4 | 41.9 | 3208 | 6545 | 47.5 | 46.4 |
| 79 | | DGAD | 8 | DGAD Hi Resp | 26-30 | Female | 32.2 | Obese | 0.19 | no-IR | 5.55 | 53 | Normal | 68 | 120.5 | 1.77 | 90.60 | 89353.4 | 36292.5 | 49985.8 | 53060.9 | 41.9 | 40.6 | 2897 | 6269 | 42.8 | 41.9 |
| 84 | | DGAD | 8 | DGAD Hi Resp | 46-50 | Female | 32.9 | Obese | 1.98 | no-IR | 5.05 | 192 | Mild-Moderate | 64 | 119.5 | 1.87 | 95.83 | 94060.7 | 38980.7 | 52667.2 | 55080 | 42.3 | 41.4 | 3666 | 6570 | 45.4 | 42.8 |
| 85 | | TAD | 8 | TAD | 46-50 | Female | 33.8 | Obese | 0.84 | no-IR | 5.29 | 92 | Normal | 67 | 121 | 1.81 | 80.7 | 79431.8 | 34482.9 | 42410.5 | 44949 | 44.6 | 43.4 | 3129 | 6834 | 46.8 | 47.5 |
| 87 | | TAD | 8 | TAD | 51-55 | Female | 30.7 | Obese | 1.03 | no-IR | 5.04 | 89 | Normal | 70 | 115.5 | 1.65 | 81.85 | 80314.1 | 27818.9 | 49933.8 | 52495.3 | 37.5 | 34.6 | 2301 | 4549 | 38.6 | 35.8 |
| 96 | | DGAD | 8 | DGAD Hi Resp | 61-65 | Female | 26.6 | Overweight | 2.44 | IR | 5.46 | 108 | Normal | 66 | 109.5 | 1.66 | 66.9 | 65952.2 | 24293.7 | 39920.7 | 41658.5 | 37.4 | 36.8 | 2056 | 4169 | 40 | 38.9 |
| 97 | | DGAD | 8 | DGAD Hi Resp | 56-60 | Female | 26.4 | Overweight | 3.29 | IR | 5.41 | 139 | Normal | 83.5 | 114 | 1.37 | 57.2 | 56279.4 | 22980.6 | 31946 | 33298.7 | 44.2 | 40.8 | 1787 | 3227 | 42.6 | 38.5 |
| 102 | | DGAD | 8 | DGAD Hi Resp | 61-65 | Female | 24.4 | Overweight | 2.43 | IR | 5.54 | 113 | Normal | 66 | 119.5 | 1.81 | 67.5 | 66497.1 | 24756.8 | 39631 | 41740.3 | 41.1 | 37.2 | 2307 | 4254 | 43.2 | 40.5 |
| 107 | | DGAD | 8 | DGAD Hi Resp | 56-60 | Female | 31.1 | Obese | 2.16 | IR | 5.5 | 60 | Normal | 64 | 121 | 1.89 | 89.85 | 88535.5 | 35359.9 | 50755.4 | 53175.6 | 42 | 39.9 | 2652 | 6209 | 40.5 | 44.3 |
| 109 | | TAD | 8 | TAD | 51-55 | Female | 26.6 | Overweight | 3.62 | IR | 5.35 | 69 | Normal | 70.5 | 114 | 1.62 | 61.65 | 60859.6 | 28519.6 | 30592.1 | 32340 | 48 | 46.9 | 2447 | 4821 | 47.5 | 47.4 |
| 112 | | TAD | 8 | TAD | 26-30 | Female | 32.9 | Obese | 2.17 | IR | 4.95 | 121 | Normal | 62.5 | 112.5 | 1.80 | 82.8 | 82360.7 | 34804.5 | 45466.1 | 47556.2 | 43.6 | 42.3 | 3391 | 6363 | 47.9 | 45.1 |
| 113 | | DGAD | 8 | DGAD Hi Resp | 51-55 | Female | 33.8 | Obese | 3.00 | IR | 5.27 | 40 | Normal | 68.5 | 119.5 | 1.74 | 104.6 | 102470.1 | 38043.5 | 61710.7 | 64426.5 | 36.5 | 37.1 | 2856 | 5921 | 35.1 | 38.1 |
| 135 | | TAD | 8 | TAD | 36-40 | Female | 31.1 | Obese | 2.09 | IR | 5.03 | 70 | Normal | 62 | 126 | 2.03 | 95.45 | 94608.8 | 39477.6 | 52994.5 | 55131.2 | 44.6 | 41.7 | 4029 | 5276 | 45.5 | 37.9 |
| 137 | | TAD | 8 | TAD | 56-60 | Female | 32.3 | Obese | 4.54 | IR | 5.36 | 130 | Normal | 77.5 | 129.5 | 1.67 | 101.45 | 98284.6 | 43677.3 | 52220.8 | 54607.3 | 45.9 | 44.4 | 4030 | 6302 | 48.5 | 44.4 |
| 139 | | DGAD | 8 | DGAD Hi Resp | 56-60 | Female | 28.8 | Overweight | 2.34 | IR | 5.22 | 69 | Normal | 80 | 130 | 1.63 | 70.85 | 70024.6 | 28768.9 | 39490.8 | 41255.6 | 42.6 | 41.1 | 2819 | 4414 | 47.5 | 40.8 |
| 144 | | TAD | 8 | TAD | 46-50 | Female | 37.3 | Obese | 1.72 | no-IR | 5.17 | 129 | Normal | 66.5 | 120.5 | 1.81 | 116 | 114147.8 | 52269.9 | 58721.3 | 61877.8 | 45.5 | 45.8 | 4254 | 8650 | 47.1 | 48.3 |
| 151 | Missing baseline blood draw | TAD | 8 | TAD | 41-45 | Female | 29.7 | Overweight | | IR | 6.01 | 177 | Mild-Moderate | 63 | 110 | 1.75 | 87.35 | 86446 | 34747.1 | 49363.9 | 51698.9 | 39.9 | 40.2 | 3101 | 5882 | 40 | 43.4 |
| 154 | | DGAD | 8 | DGAD Low Resp | 61-65 | Female | 35.1 | Obese | 1.40 | no-IR | 5.7 | 139 | Normal | 78 | 121.5 | 1.56 | 92.6 | 91651.1 | 42198 | 47424 | 49453.1 | 44.8 | 46 | 2864 | 7139 | 43 | 46.4 |
| 168 | | TAD | 8 | TAD | 46-50 | Female | 25.8 | Overweight | 0.86 | no-IR | 5.18 | 76 | Normal | 79.5 | 134 | 1.69 | 69.65 | 68909 | 23885.3 | 42895.3 | 45023.6 | 27.4 | 34.7 | 1093 | 4044 | 25.3 | 38.5 |
| 170 | | TAD | 8 | TAD | 51-55 | Female | 37.1 | Obese | 6.44 | IR | 5.56 | 72 | Normal | 70 | 131 | 1.87 | 98.95 | 96893.8 | 48194.7 | 46440.2 | 48699.1 | 51.3 | 49.7 | 4378 | 8229 | 52.3 | 51.5 |
| 172 | | DGAD | 8 | DGAD Hi Resp | 46-50 | Female | 32.2 | Obese | 1.75 | no-IR | 5.13 | 81 | Normal | 65.5 | 114 | 1.74 | 92.6 | 91471.5 | 39152.1 | 50063.3 | 52319.5 | 40.1 | 42.8 | 2637 | 7906 | 40.1 | 49.2 |
| 175 | | TAD | 8 | TAD | 51-55 | Female | 27.4 | Overweight | 2.49 | IR | 5.98 | 59 | Normal | 66.5 | 111 | 1.67 | 80.1 | 78419.2 | 30135.9 | 46168.3 | 48283.3 | 37.9 | 38.4 | 2645 | 5383 | 44 | 42.6 |
| 180 | | TAD | 8 | TAD | 61-65 | Female | 30.5 | Obese | 2.56 | IR | 5.42 | 237 | Mild-Moderate | 66.5 | 116.5 | 1.75 | 81.15 | 79888.8 | 34783.6 | 43254.7 | 45105.2 | 41.3 | 43.5 | 2612 | 5947 | 44.3 | 45.8 |
| 182 | | DGAD | 8 | DGAD Low Resp | 26-30 | Female | 33.1 | Obese | 2.09 | IR | 5.55 | 48 | Normal | 68 | 120.5 | 1.77 | 106.1 | 103286.6 | 45614.2 | 54969.5 | 57672.4 | 47 | 44.2 | 4396 | 8232 | 51.4 | 50.4 |
| 183 | | TAD | 8 | TAD | 56-60 | Female | 39 | Obese | 5.27 | IR | 5.23 | 222 | Mild-Moderate | 72 | 104 | 1.44 | 105.3 | 103554.9 | 48461.8 | 52878.6 | 55093.2 | 47.6 | 46.8 | 4643 | 8227 | 47.4 | 47.2 |
| 186 | | TAD | 8 | TAD | 51-55 | Female | 38 | Obese | 2.14 | IR | 5.74 | 71 | Normal | 68 | 119.5 | 1.76 | 101.7 | 99937.2 | 45561.4 | 52135.8 | 54375.8 | 45.8 | 45.6 | 4071 | 7609 | 49.8 | 47.8 |
| 189 | | DGAD | 8 | DGAD Low Resp | 61-65 | Female | 29.9 | Overweight | 3.04 | IR | 5.71 | 150 | Mild-Moderate | 73.5 | 138.5 | 1.88 | 75.35 | 74441.6 | 32573 | 39944.2 | 41868.7 | 47.6 | 43.8 | 3346 | 4899 | 50.2 | 43.5 |
| 194 | | DGAD | 8 | DGAD Low Resp | 61-65 | Female | 27.4 | Overweight | 1.87 | no-IR | 5.61 | 240 | Mild-Moderate | 79 | 129.5 | 1.64 | 70.25 | 69554.4 | 33601.4 | 34474.6 | 35953 | 54 | 48.3 | 3746 | 5296 | 55.4 | 47.4 |
| 198 | screening and Visit 1 TG mismatch | DGAD | 8 | DGAD Hi Resp | 51-55 | Female | 38 | Obese | 13.34 | IR | 5.38 | 266 | Mild-Moderate | 71.5 | 119.5 | 1.67 | 102.45 | 100720.6 | 45818 | 52504.9 | 54902.6 | 47.1 | 45.5 | 4847 | 8511 | 51.3 | 49.1 |
| 232 | | TAD | 8 | TAD | 61-65 | Female | 29.7 | Overweight | 4.16 | IR | 5.33 | 361 | Mild-Moderate | 67.5 | 117.5 | 1.74 | 74.1 | 72985.8 | 32997.9 | 38331.9 | 39987.9 | 46.9 | 45.2 | 3165 | 6025 | 48.8 | 47.5 |
| 234 | | DGAD | 8 | DGAD Low Resp | 56-60 | Female | 36.2 | Obese | 5.80 | IR | 5.94 | 355 | Mild-Moderate | 80 | 127 | 1.59 | 99.65 | 97344.9 | 45433.5 | 50078.2 | 51911.3 | 52.8 | 46.7 | 5800 | 5627 | 53.9 | 38.9 |
| 239 | | TAD | 8 | TAD | 51-55 | Female | 29.2 | Overweight | 6.74 | IR | 5.62 | 53 | Normal | 69.5 | 107 | 1.54 | 80.45 | 79126.6 | 33418.4 | 42980.3 | 45708.2 | 42.8 | 42.2 | 2334 | 5599 | 40.7 | 44.1 |
| 243 | | DGAD | 8 | DGAD Low Resp | 56-60 | Female | 26 | Overweight | 8.22 | IR | 5.37 | 92 | Normal | 69.5 | 104 | 1.50 | 74.7 | 74716.6 | 27728.4 | 44882.3 | 46988.3 | 37.7 | 37.1 | 2111 | 4834 | 36.9 | 40.1 |
| 244 | | DGAD | 8 | DGAD Hi Resp | 36-40 | Female | 34.6 | Obese | 1.07 | no-IR | 5.36 | 88 | Normal | 79 | 111.5 | 1.41 | 100.85 | 99106.5 | 44062.7 | 52512.6 | 55043.9 | 46.5 | 44.5 | 4047 | 6404 | 49 | 43.2 |
| 245 | | DGAD | 8 | DGAD Hi Resp | 61-65 | Female | 37.8 | Obese | 2.57 | IR | 5.98 | 72 | Normal | 75 | 123.5 | 1.65 | 106.8 | 104798.4 | 47034.1 | 55116.5 | 57764.3 | 46.6 | 44.9 | 3815 | 7086 | 45.6 | 45.2 |
| 258 | | DGAD | 8 | DGAD Low Resp | 56-60 | Female | 29.4 | Overweight | 2.95 | IR | 5.52 | 136 | Normal | 65 | 107 | 1.65 | 78.15 | 76947.7 | 35790.5 | 39056 | 41157.2 | 49 | 46.5 | 2884 | 5619 | 45.2 | 46.6 |

| Android:Gynoid Ratio | Total BMC (g) | Total BMD (g/cm^2) | RBC FAME Sample ID: | RBC FAME Acq. Date-Time: | RBC FAME Analysis Batch: | TG-16:0-d31 % Rec. | Wk | OM3-I | C14:0 | C15:0 | C16:0 | C17:0 | C18:0 | C18:1n9 | C18:1n7 | C18:2n6 | C20:3n6 | C20:4n6 | C22:4n6 | C22:5n6 | C20:5n3 | C22:5n3 | C22:6n3 | Σ SFA | Σ MUFA | Σ (n-6) PUFA | Σ (n-3) PUFA | Total Fatty Acids | % C14:0 | % C15:0 | % C16:0 | % C17:0 |
|---|---|---|---|---|---|---|---|---|---|---|---|---|---|---|---|---|---|---|---|---|---|---|---|---|---|---|---|---|---|---|---|---|
| 0.536154383 | 2605.22 | 1.229 | FL95-003 Wk1B1 | 7/25/2018 21:32 | Batch 2 | 37.2 | 1 | 4.95 | 9.15 | 3.54 | 302.21 | 5.63 | 323.77 | 115.62 | 23.66 | 78.26 | 18.38 | 102.86 | 45.85 | 7.9 | 5.37 | 36.52 | 50.5 | 644.3 | 139.28 | 253.25 | 92.39 | 1129.22 | 0.81 | 0.31 | 26.76 | 0.50 |
| 0.378768021 | 1828.6 | 0.989 | FL95-009 Wk1B1 | 7/26/2018 15:40 | Batch 3 | 46.7 | 1 | 5.23 | 33.9 | 5.32 | 485.65 | 5.46 | 334.67 | 164.88 | 32.89 | 109.1 | 23.75 | 152.01 | 70.61 | 13.97 | 4.36 | 37.14 | 76.74 | 865 | 197.77 | 369.44 | 118.24 | 1550.45 | 2.19 | 0.34 | 31.32 | 0.35 |
| 0.370094247 | 2321.25 | 1.236 | FL95-020 Wk1B1 | 7/27/2018 7:13 | Batch 3 | 74.2 | 1 | 4.07 | 14.25 | 2.53 | 285.04 | 1.47 | 139.8 | 101.89 | 20.91 | 56.48 | 9.41 | 75.18 | 28.39 | 5.87 | 2.61 | 25.56 | 29.93 | 443.09 | 122.8 | 175.33 | 58.1 | 799.32 | 1.78 | 0.32 | 35.66 | 0.18 |
| 0.558545145 | 2728.41 | 1.231 | FL95-032 Wk1B1 | 7/25/2018 20:02 | Batch 2 | 48.4 | 1 | 5.70 | 11.48 | 3.23 | 312.17 | 3.61 | 260.18 | 117.12 | 20.19 | 52.51 | 16.61 | 86.12 | 43.33 | 7.88 | 3.31 | 28.15 | 54.87 | 590.67 | 137.31 | 206.45 | 86.33 | 1020.76 | 1.12 | 0.32 | 30.58 | 0.35 |
| 0.410074495 | 2569.12 | 1.319 | FL95-051 Wk1B1 | 7/25/2018 19:39 | Batch 2 | 45.1 | 1 | 4.41 | 9.27 | 2.97 | 264.58 | 4.45 | 270.32 | 86.19 | 12.84 | 42.25 | 10.79 | 62.36 | 36.37 | 7.35 | 2.24 | 20.19 | 36.02 | 551.59 | 99.03 | 159.12 | 58.45 | 868.19 | 1.07 | 0.34 | 30.48 | 0.51 |
| 0.623785884 | 1791.73 | 1.023 | FL95-052 Wk1B1 | 7/27/2018 5:45 | Batch 3 | 57.2 | 1 | 6.25 | 20.24 | 3.32 | 322.54 | 3.05 | 188.14 | 116.28 | 24.36 | 72.17 | 14.66 | 92.42 | 31.02 | 6.29 | 4.26 | 26.73 | 57.16 | 537.29 | 140.64 | 216.56 | 88.15 | 982.64 | 2.06 | 0.34 | 32.82 | 0.31 |
| 0.503442999 | 1880.49 | 1.088 | FL95-053 Wk1B1 | 7/27/2018 4:39 | Batch 3 | 90.9 | 1 | 4.54 | 25.27 | 5.44 | 547.07 | 6.98 | 294.72 | 213.05 | 29.83 | 132.03 | 16.76 | 205.75 | 63.51 | 16.03 | 9.22 | 56.82 | 67.59 | 879.48 | 242.88 | 434.08 | 133.63 | 1690.07 | 1.50 | 0.32 | 32.37 | 0.41 |
| 0.646767866 | 1884.09 | 1.112 | FL95-069 Wk1B1 | 7/25/2018 7:10 | Batch 1 | 39.9 | 1 | 4.82 | 3.71 | 1.31 | 268.84 | 2.34 | 261.68 | 80.78 | 16.81 | 46.23 | 9.65 | 87.7 | 40.96 | 12.65 | 2.7 | 25.96 | 40.74 | 537.88 | 97.59 | 197.19 | 69.4 | 902.06 | 0.41 | 0.15 | 29.80 | 0.26 |
| 0.472998683 | 2361.57 | 1.192 | FL95-072 Wk1B1 | 7/26/2018 19:45 | Batch 3 | 54.2 | 1 | 6.43 | 23.89 | 3.46 | 376.86 | 7.37 | 207.04 | 147.68 | 31.06 | 58.58 | 12.37 | 76.88 | 38.92 | 11.34 | 2.67 | 20.2 | 67.18 | 618.62 | 178.74 | 198.09 | 90.05 | 1085.5 | 2.20 | 0.32 | 34.72 | 0.68 |
| 0.491625352 | 3063.34 | 1.441 | FL95-079 Wk1B1 | 7/25/2018 22:39 | Batch 2 | 39.8 | 1 | 7.53 | 16.47 | 3.04 | 201.06 | 4.68 | 265.05 | 73.87 | 16.02 | 62.46 | 8.97 | 67.45 | 31.56 | 7.2 | 2.37 | 21.98 | 61.13 | 490.3 | 89.89 | 177.64 | 85.48 | 843.31 | 1.95 | 0.36 | 23.84 | 0.55 |
| 0.577878104 | 2446.91 | 1.193 | FL95-084 Wk1B1 | 7/25/2018 7:32 | Batch 1 | 57.1 | 1 | 5.63 | 12.87 | 3.59 | 420.33 | 4.1 | 287.26 | 127.34 | 27.98 | 87.87 | 21.98 | 110.4 | 53.06 | 15.63 | 4.61 | 33.91 | 67.32 | 728.15 | 155.32 | 288.94 | 105.84 | 1278.25 | 1.01 | 0.28 | 32.88 | 0.32 |
| 0.462609664 | 2484.3 | 1.319 | FL95-085 Wk1B1 | 7/24/2018 21:53 | Batch 1 | 40.1 | 1 | 4.95 | 2.85 | 5.72 | 377.6 | 8.32 | 392.73 | 151.15 | 27.14 | 96.89 | 21.73 | 168.35 | 74.56 | 17.6 | 7.75 | 79.85 | 66.41 | 787.22 | 178.29 | 379.13 | 154.01 | 1498.65 | 0.19 | 0.38 | 25.20 | 0.56 |
| 0.576931592 | 2544.75 | 1.282 | FL95 087 reshoot | 7/30/2018 11:10 | Batch x | 64.2 | 1 | 5.84 | 7.92 | 1.52 | 277.71 | 1.43 | 127.31 | 90.96 | 19.57 | 51.73 | 8.06 | 88.95 | 29.54 | 4.76 | 3.07 | 17.68 | 42.02 | 415.89 | 110.53 | 183.04 | 62.77 | 772.23 | 1.03 | 0.20 | 35.96 | 0.19 |
| 0.510572139 | 1732.09 | 1.001 | FL95-096 Wk1B1 | 7/24/2018 22:37 | Batch 1 | 46.5 | 1 | 7.97 | 1.01 | 2.44 | 255.86 | 5.34 | 232.71 | 81.39 | 12.51 | 47.17 | 9.14 | 78.51 | 41.19 | 8.18 | 5.37 | 27.19 | 64.17 | 497.36 | 93.9 | 184.19 | 96.73 | 872.18 | 0.12 | 0.28 | 29.34 | 0.61 |
| 0.613471829 | 1332.71 | 0.909 | FL95-097 Wk1B1 | 7/26/2018 15:18 | Batch 3 | 43.1 | 1 | 5.31 | 26.84 | 4.98 | 275.69 | 5.15 | 253.3 | 94.34 | 20.91 | 63.28 | 15.79 | 69.38 | 37.01 | 5.02 | 3.25 | 24.26 | 47.01 | 565.96 | 115.25 | 190.48 | 74.52 | 946.21 | 2.84 | 0.53 | 29.14 | 0.54 |
| 0.640874882 | 2050.85 | 1.107 | FL95-102 Wk1B1 | 7/25/2018 0:52 | Batch 1 | 33.1 | 1 | 7.40 | 2.85 | 2.99 | 395.72 | 5.78 | 414.24 | 168.38 | 24.4 | 99.32 | 18.76 | 145.76 | 61.4 | 15.93 | 9.4 | 68.66 | 104.41 | 821.58 | 192.78 | 341.17 | 182.47 | 1538 | 0.19 | 0.19 | 25.73 | 0.38 |
| 0.473743557 | 2404.44 | 1.172 | FL95-107 Wk1B1 | 7/27/2018 2:48 | Batch 3 | 33.1 | 1 | 6.74 | 9.63 | 4.49 | 138.2 | 5.08 | 307.66 | 72.81 | 16.61 | 58.69 | 9.54 | 99.15 | 48.14 | 4.24 | 3.18 | 31.87 | 55.09 | 465.06 | 89.42 | 219.76 | 90.14 | 864.38 | 1.11 | 0.52 | 15.99 | 0.59 |
| 0.44671799 | 1713.24 | 1.052 | FL95-109 Wk1B1 | 7/25/2018 11:54 | Batch 1 | 45.7 | 1 | 6.74 | 8.57 | 2.95 | 232.26 | 3.81 | 251.09 | 74.13 | 17.55 | 51.15 | 10.21 | 82.45 | 40.49 | 9.69 | 2.58 | 24.52 | 55.85 | 498.68 | 91.68 | 193.99 | 82.95 | 867.3 | 0.99 | 0.34 | 26.78 | 0.44 |
| 0.514775228 | 2089.51 | 1.136 | FL95-112 Wk1B1 | 7/26/2018 7:09 | Batch 2 | 48.7 | 1 | 5.79 | 11.06 | 2.47 | 246.27 | 2.76 | 179.72 | 81.13 | 12.12 | 59.91 | 11.25 | 72.95 | 38.83 | 5.59 | 2.45 | 22.69 | 43.41 | 442.28 | 93.25 | 188.53 | 68.55 | 792.61 | 1.40 | 0.31 | 31.07 | 0.35 |
| 0.4729832 | 2650.98 | 1.182 | FL95-113 Wk1B1 | 7/25/2018 18:10 | Batch 2 | 54.7 | 1 | 5.62 | 6.99 | 2.42 | 241.79 | 3.88 | 266.64 | 88.14 | 14.91 | 71.41 | 10.42 | 89.46 | 45.51 | 10.63 | 3.11 | 21.6 | 48.88 | 521.72 | 103.05 | 227.43 | 73.59 | 925.79 | 0.76 | 0.26 | 26.12 | 0.42 |
| 0.724881641 | 2108.26 | 1.054 | FL95-135 Wk1B1 | 7/26/2018 18:15 | Batch 3 | 26.6 | 1 | 5.49 | 33.94 | 2.55 | 320.65 | 8.1 | 302.32 | 128.84 | 28.04 | 69.17 | 13.17 | 117.7 | 52.11 | 12.26 | 3.8 | 45.46 | 62.11 | 667.56 | 156.88 | 264.41 | 111.37 | 1200.22 | 2.83 | 0.21 | 26.72 | 0.67 |
| 0.681390811 | 2439.52 | 1.12 | FL95-137 Wk1B1 | 7/26/2018 9:22 | Batch 2 | 47.2 | 1 | 3.92 | 23.31 | 4.42 | 385.71 | 5.41 | 368.63 | 170.9 | 27.3 | 89.57 | 18.77 | 124.12 | 57.97 | 14.55 | 3.41 | 37.15 | 50.71 | 787.48 | 198.2 | 304.98 | 91.27 | 1381.93 | 1.69 | 0.32 | 27.91 | 0.39 |
| 0.67673507 | 1699.34 | 1.023 | FL95-139 Wk1B1 | 7/27/2018 3:32 | Batch 3 | 56.4 | 1 | 3.59 | 21.51 | 5.52 | 498.22 | 5.16 | 330.08 | 123.66 | 18.37 | 99.21 | 18.06 | 114.97 | 49.9 | 8.81 | 4.76 | 33.74 | 44.59 | 860.49 | 142.03 | 290.95 | 83.09 | 1376.56 | 1.56 | 0.40 | 36.19 | 0.37 |
| 0.480089803 | 3217.85 | 1.401 | FL95-144 Wk1B1 | 7/27/2018 10:32 | Batch 3 | 60.1 | 1 | 7.66 | 23.97 | 4.85 | 366.55 | 3.9 | 189.45 | 143.08 | 32.02 | 67.58 | 14.47 | 81.26 | 30.35 | 3.35 | 5.37 | 17.39 | 75.72 | 588.72 | 175.1 | 197.01 | 98.48 | 1059.31 | 2.26 | 0.46 | 34.60 | 0.37 |
| 0.509427313 | 2391.48 | 1.152 | | | | | | | | | | | | | | | | | | | | | | | | | | | | | | |
| 0.399098382 | 2080.57 | 1.035 | FL95 154 reshoot | 7/30/2018 10:48 | Batch x | 70.4 | 1 | 5.27 | 1.29 | 3.34 | 214.68 | 1.2 | 157.13 | 59.81 | 16.64 | 40.63 | 7.63 | 60.25 | 21.71 | 6.24 | 2.44 | 16.68 | 31.36 | 377.64 | 76.45 | 136.46 | 50.48 | 641.03 | 0.20 | 0.52 | 33.49 | 0.19 |
| 0.278699862 | 2154.87 | 1.072 | FL95-168 Wk1B1 | 7/26/2018 11:12 | Batch 2 | 49.3 | 1 | 9.07 | 6.88 | 1.01 | 180.52 | 0.92 | 148.27 | 55.44 | 13.88 | 44.39 | 8.08 | 40.67 | 10.18 | 0.44 | 6.15 | 25.33 | 47.3 | 337.6 | 69.32 | 103.76 | 78.78 | 589.45 | 1.17 | 0.17 | 30.63 | 0.16 |
| 0.513389071 | 2222.32 | 1.143 | FL95-170 Wk1B1 | 7/26/2018 19:22 | Batch 3 | 22.7 | 1 | 6.04 | 21.66 | 4 | 296.28 | 6.72 | 146.57 | 124.63 | 26.21 | 72.17 | 13.7 | 113.15 | 59.84 | 10.31 | 3.51 | 43.29 | 56.87 | 475.23 | 150.84 | 269.17 | 103.67 | 998.91 | 2.17 | 0.40 | 29.66 | 0.67 |
| 0.369747899 | 2321.12 | 1.115 | FL95-172 Wk1B1 | 7/25/2018 17:26 | Batch 2 | 33.8 | 1 | 7.05 | 1.21 | 1.1 | 164.85 | 2.72 | 270.57 | 61.89 | 8.23 | 38.49 | 7.49 | 63.15 | 31.5 | 5.18 | 2.28 | 22.72 | 49.2 | 440.45 | 70.12 | 145.81 | 74.2 | 730.58 | 0.17 | 0.15 | 22.56 | 0.37 |
| 0.483482387 | 2169.56 | 1.12 | FL95-175 Wk1B1 | 7/24/2018 22:15 | Batch 1 | 44.8 | 1 | 6.65 | 7.42 | 4.52 | 344.67 | 7.44 | 292.45 | 132.27 | 24.84 | 102.08 | 17.93 | 127.78 | 48.45 | 10.34 | 8.02 | 47.29 | 75.19 | 656.5 | 157.11 | 306.58 | 130.5 | 1250.69 | 0.59 | 0.36 | 27.56 | 0.59 |
| 0.437147887 | 1865.21 | 0.989 | FL95-180 Wk1B1 | 7/26/2018 11:34 | Batch 2 | 48.8 | 1 | 7.18 | 11.29 | 1.19 | 230.23 | 3.3 | 247.67 | 119.55 | 18.48 | 57.12 | 10.44 | 80.35 | 39.19 | 8.11 | 3.51 | 20.92 | 62.04 | 493.68 | 138.03 | 195.21 | 86.47 | 913.39 | 1.24 | 0.13 | 25.21 | 0.36 |
| 0.513319795 | 2615.64 | 1.192 | FL95-182 Wk1B1 | 7/25/2018 7:54 | Batch 1 | 40.5 | 1 | 5.90 | 3.72 | 3.27 | 301.62 | 5.95 | 374.87 | 111.41 | 16.94 | 77.63 | 17 | 112.12 | 44.76 | 10.43 | 4.7 | 34.59 | 65.15 | 689.43 | 128.35 | 261.94 | 104.44 | 1184.16 | 0.31 | 0.28 | 25.47 | 0.50 |
| 0.638371376 | 2120.41 | 1.112 | FL95-183 Wk1B1 | 7/25/2018 4:13 | Batch 1 | 45.2 | 1 | 5.05 | 7.65 | 3.57 | 268.26 | 4.82 | 299.75 | 70.34 | 11.63 | 45.39 | 7.85 | 65.69 | 31.62 | 4.89 | 3.84 | 24.99 | 41.14 | 584.05 | 81.97 | 155.44 | 69.97 | 891.43 | 0.86 | 0.40 | 30.09 | 0.54 |
| 0.553149363 | 2223.59 | 1.151 | FL95-186 Wk1B1 | 7/26/2018 3:27 | Batch 2 | 48.3 | 1 | 5.67 | 6.01 | 2.91 | 205.98 | 2.8 | 199.33 | 86.49 | 14.67 | 44.33 | 10.23 | 84.26 | 37.1 | 7.84 | 2.47 | 23.16 | 41.08 | 417.03 | 101.16 | 183.76 | 66.71 | 768.66 | 0.78 | 0.38 | 26.80 | 0.36 |
| 0.636777801 | 1871.36 | 1.013 | FL95-189 Wk1B1 | 7/25/2018 6:48 | Batch 1 | 46.3 | 1 | 4.24 | 13.01 | 3.24 | 349.78 | 5 | 328.46 | 176.45 | 23.82 | 90.39 | 14.14 | 127.47 | 56.08 | 18.2 | 2.09 | 35.86 | 52.87 | 699.49 | 200.27 | 306.28 | 90.82 | 1296.86 | 1.00 | 0.25 | 26.97 | 0.39 |
| 0.669859155 | 1466.04 | 0.892 | FL95-194 Wk1B1 | 7/26/2018 5:41 | Batch 2 | 44.5 | 1 | 4.50 | 16.05 | 3.03 | 337.9 | 4.81 | 324.5 | 167.78 | 23.71 | 67.8 | 14.19 | 144.95 | 60.35 | 14.6 | 6.52 | 47.35 | 51.35 | 686.29 | 191.49 | 301.89 | 105.22 | 1284.89 | 1.25 | 0.24 | 26.30 | 0.37 |
| 0.574977817 | 2317.27 | 1.204 | FL95-198 Wk1B1 | 7/27/2018 7:57 | Batch 3 | 58.6 | 1 | 4.02 | 16.98 | 2.32 | 291.57 | 3.06 | 184.77 | 99.16 | 15.73 | 45.28 | 10.68 | 58.55 | 26.98 | 4.03 | 2.32 | 15.21 | 30.09 | 498.7 | 114.89 | 145.52 | 47.62 | 806.73 | 2.10 | 0.29 | 36.14 | 0.38 |
| 0.499594649 | 1633.04 | 0.907 | FL95-232 Wk1B1 | 7/25/2018 13:00 | Batch 1 | 42.0 | 1 | 8.04 | 10.38 | 2.56 | 330.04 | 2.91 | 278.15 | 136.68 | 24.65 | 83.39 | 12.54 | 119.41 | 41.38 | 10.77 | 4.46 | 35.74 | 90.7 | 624.04 | 161.33 | 267.49 | 130.9 | 1183.76 | 0.88 | 0.22 | 27.88 | 0.25 |
| 1.098057018 | 1769.82 | 0.946 | FL95-234 Wk1B1 | 7/26/2018 21:37 | Batch 3 | 53.2 | 1 | 3.89 | 24.23 | 5.91 | 289.22 | 3.77 | 149.31 | 114.78 | 24.09 | 65.84 | 20.54 | 81.74 | 41.43 | 5.54 | 3.36 | 18.28 | 30.82 | 472.44 | 138.87 | 215.09 | 52.46 | 878.86 | 2.76 | 0.67 | 32.91 | 0.43 |
| 0.452833573 | 2634.06 | 1.302 | FL95-239 Wk1B1 | 7/25/2018 6:26 | Batch 1 | 25.9 | 1 | 6.87 | 10.87 | 18.41 | 232.42 | 3.11 | 250.38 | 73.68 | 15.16 | 48.76 | 11.14 | 89.85 | 40.6 | 8.46 | 5 | 28.29 | 56.29 | 515.19 | 88.84 | 198.81 | 89.58 | 892.42 | 1.22 | 2.06 | 26.04 | 0.35 |
| 0.467839806 | 2091.1 | 1.044 | FL95-243 Wk1B1 | 7/26/2018 4:34 | Batch 2 | 42.2 | 1 | 7.03 | 23.37 | 4.73 | 283.16 | 4.69 | 237.38 | 112.73 | 15.23 | 55.33 | 12.75 | 95.56 | 35.04 | 6.25 | 6.57 | 34.93 | 63.11 | 553.33 | 127.96 | 204.93 | 104.61 | 990.83 | 2.36 | 0.48 | 28.58 | 0.47 |
| 0.611601705 | 2601.22 | 1.27 | FL95-244 Wk1B1 | 7/26/2018 4:12 | Batch 2 | 45.4 | 1 | 4.29 | 6.89 | 2.01 | 169.26 | 2.17 | 197.53 | 66.63 | 12.56 | 36.39 | 13 | 51.66 | 30.12 | 6.7 | 2.26 | 21.74 | 25.36 | 377.86 | 79.19 | 137.87 | 49.36 | 644.28 | 1.07 | 0.31 | 26.27 | 0.34 |
| 0.608019066 | 2574.74 | 1.227 | FL95-245 Wk1B1 | 7/27/2018 5:23 | Batch 3 | 53.0 | 1 | 5.31 | 27.4 | 5.52 | 455.95 | 7.34 | 363.88 | 195.12 | 43.73 | 104.2 | 20.02 | 202.64 | 80.34 | 20.71 | 6.29 | 48.96 | 82.05 | 860.09 | 238.85 | 427.91 | 137.3 | 1664.15 | 1.65 | 0.33 | 27.40 | 0.44 |
| 0.49172618 | 2075.43 | 1.122 | FL95-258 Wk1B1 | 7/25/2018 20:24 | Batch 2 | 48.0 | 1 | 5.15 | 5.79 | 3.2 | 308.7 | 4.36 | 240.25 | 142.86 | 21.49 | 61.49 | 20.38 | 110.27 | 44.69 | 11.38 | 5.27 | 42.08 | 49.95 | 562.3 | 164.35 | 248.21 | 97.3 | 1072.16 | 0.54 | 0.30 | 28.79 | 0.41 |
| | | | FL95-003 Wk3B1 | 7/26/2018 12:41 | Batch 2 | 35.5 | 3 | 6.07 | 6.96 | 1.52 | 267.55 | 1.31 | 259.18 | 106.94 | 23.91 | 76.73 | 19.42 | 113.32 | 47.22 | 5.9 | 5.05 | 39.15 | 57.62 | 536.52 | 130.85 | 262.59 | 101.82 | 1031.79 | 0.67 | 0.15 | 25.93 | 0.13 |
| | | | FL95-009 Wk3B1 | 7/26/2018 9:00 | Batch 2 | 107.7 | 3 | 3.92 | 4.8 | 1.98 | 320.07 | 3.54 | 392.25 | 99.62 | 15.52 | 57.73 | 11.15 | 86.47 | 33.96 | 7.92 | 1.97 | 20.14 | 41.14 | 722.64 | 115.14 | 197.23 | 63.25 | 1098.26 | 0.44 | 0.18 | 29.14 | 0.32 |
| | | | FL95-020 Wk3B1 | 7/25/2018 3:50 | Batch 1 | 51.3 | 3 | 5.38 | 8.4 | 3.48 | 344.18 | 4.71 | 265.43 | 129.34 | 23.75 | 82.62 | 17.42 | 146.43 | 61.45 | 15.85 | 3.74 | 57.02 | 62.25 | 626.2 | 153.09 | 323.77 | 123.01 | 1226.07 | 0.69 | 0.28 | 28.07 | 0.38 |
| | | | FL95-032 Wk3B1 | 7/26/2018 11:57 | Batch 2 | 52.5 | 3 | 5.80 | 16.34 | 3.02 | 343.48 | 4.72 | 262.69 | 135.5 | 20.92 | 59.93 | 18.55 | 101.4 | 50.51 | 9.06 | 3.86 | 25.38 | 60.9 | 630.25 | 156.42 | 239.45 | 90.14 | 1116.26 | 1.46 | 0.27 | 30.77 | 0.42 |
| | | | FL95-051 Wk3B1 | 7/27/2018 3:10 | Batch 3 | 58.6 | 3 | 4.99 | 22.63 | 3.67 | 288.3 | 3.66 | 189.64 | 97.34 | 15.62 | 49.41 | 11.83 | 78.82 | 38.16 | 7.87 | 3.29 | 24.41 | 40.38 | 507.9 | 112.96 | 186.09 | 68.08 | 875.03 | 2.59 | 0.42 | 32.95 | 0.42 |
| | | | FL95-052 Wk3B1 | 7/26/2018 20:30 | Batch 3 | 50.4 | 3 | 5.58 | 25.2 | 4.52 | 368.36 | 5.09 | 221.58 | 116.81 | 19.71 | 69.84 | 15.23 | 94.33 | 37.65 | 10.23 | 3.41 | 31.89 | 56.93 | 624.75 | 136.52 | 227.28 | 92.23 | 1080.78 | 2.33 | 0.42 | 34.08 | 0.47 |
| | | | FL95-053 Wk3B1 | 7/27/2018 9:26 | Batch 3 | 58.7 | 3 | 4.96 | 15.15 | 2.08 | 362.71 | 2.52 | 145.88 | 165.68 | 34.47 | 87.23 | 13.42 | 127.01 | 45.1 | 6.09 | 4.39 | 33.63 | 49.89 | 528.34 | 200.15 | 278.85 | 87.91 | 1095.25 | 1.38 | 0.19 | 33.12 | 0.23 |
| | | | FL95-069 Wk3B1 | 7/25/2018 0:30 | Batch 1 | 39.8 | 3 | 5.23 | 1.43 | 3.5 | 462 | 6.11 | 480.76 | 177.41 | 34.05 | 101.57 | 15.51 | 219.72 | 96.75 | 23.26 | 4.3 | 68.02 | 89.02 | 953.8 | 211.46 | 456.81 | 161.34 | 1783.41 | 0.08 | 0.20 | 25.91 | 0.34 |
| | | | FL95-072 Wk3B1 | 7/27/2018 8:42 | Batch 3 | 62.8 | 3 | 6.88 | 24.3 | 2.13 | 368.49 | 1.11 | 140.42 | 141.85 | 40.27 | 44.26 | 12.03 | 74.39 | 33.24 | 9.56 | 2.93 | 15.5 | 64.13 | 536.45 | 182.12 | 173.48 | 82.56 | 974.61 | 2.49 | 0.22 | 37.81 | 0.11 |
| | | | FL95-079 Wk3B1 | 7/26/2018 10:06 | Batch 2 | 47.0 | 3 | 7.15 | 6.85 | 2.94 | 218.91 | 2.34 | 249.76 | 71.01 | 14.75 | 62.34 | 9.17 | 65.08 | 28.61 | 8.79 | 2.85 | 24.25 | 56.03 | 480.8 | 85.76 | 173.99 | 83.13 | 823.68 | 0.83 | 0.36 | 26.58 | 0.28 |
| | | | FL95-084 Wk3B1 | 7/26/2018 8:38 | Batch 2 | 42.6 | 3 | 5.81 | 4.18 | 1.42 | 295.43 | 2.07 | 209.14 | 93.01 | 15.83 | 55.85 | 18.53 | 73.83 | 39.9 | 8.88 | 3.06 | 19.47 | 48.57 | 512.24 | 108.84 | 196.99 | 71.1 | 889.17 | 0.47 | 0.16 | 33.23 | 0.23 |
| | | | FL95-085 Wk3B1 | 7/25/2018 5:42 | Batch 1 | 48.1 | 3 | 5.02 | 0.96 | 3.56 | 464.14 | 6.3 | 360.48 | 173.92 | 36.34 | 108.65 | 21.43 | 191.63 | 76.57 | 21.84 | 6.64 | 76.56 | 74.96 | 835.44 | 210.26 | 420.12 | 158.16 | 1623.98 | 0.06 | 0.22 | 28.58 | 0.39 |
| | | | FL95-087 Wk3B1 | 7/25/2018 18:32 | Batch 2 | 46.5 | 3 | 5.80 | 9.19 | 2.56 | 411.65 | 5.89 | 389.97 | 177.83 | 28.4 | 106.4 | 12.19 | 177.53 | 62.25 | 11.75 | 4.35 | 47.86 | 84.54 | 819.26 | 206.23 | 370.12 | 136.75 | 1532.36 | 0.60 | 0.17 | 26.86 | 0.38 |
| | | | FL95-096 Wk3B1 | 7/25/2018 14:07 | Batch 1 | 41.6 | 3 | 7.83 | 8.42 | 2.85 | 393.6 | 4.74 | 324.95 | 157.88 | 29.98 | 90.02 | 14.46 | 154.2 | 62.84 | 16.53 | 7.25 | 47.21 | 103.8 | 734.56 | 187.86 | 338.05 | 158.26 | 1418.74 | 0.59 | 0.20 | 27.74 | 0.33 |
| | | | FL95-097 Wk3B1 | 7/26/2018 16:02 | Batch 3 | 49.6 | 3 | 5.12 | 16.43 | 5.37 | 514.65 | 6.77 | 392.87 | 211.83 | 41.8 | 120.69 | 22.42 | 147.5 | 60.74 | 16.13 | 4.28 | 50.21 | 82.45 | 936.09 | 253.63 | 367.48 | 136.94 | 1694.14 | 0.97 | 0.32 | 30.38 | 0.40 |
| | | | FL95-102 Wk3B1 | 7/27/2018 6:29 | Batch 3 | 72.6 | 3 | 6.13 | 19.59 | 4.93 | 246.26 | 2.82 | 178.75 | 91.68 | 17.96 | 39.81 | 10.53 | 60.45 | 23.03 | 2.6 | 5.05 | 23.85 | 42.13 | 452.35 | 109.64 | 136.42 | 71.03 | 769.44 | 2.55 | 0.64 | 32.01 | 0.37 |
| | | | FL95-107 Wk3B1 | 7/27/2018 9:48 | Batch 3 | 55.6 | 3 | 5.49 | 23.14 | 2.33 | 246.24 | 2.7 | 89.82 | 61.2 | 8.08 | 27.22 | 6.6 | 48.13 | 20.78 | 3.17 | 3.19 | 9.36 | 28.66 | 364.23 | 69.28 | 105.9 | 41.21 | 580.62 | 3.99 | 0.40 | 42.41 | 0.47 |
| | | | FL95-109 Wk3B1 | 7/25/2018 13:45 | Batch 1 | 36.7 | 3 | 6.38 | 6.52 | 2.25 | 292.73 | 3.11 | 306.58 | 92.93 | 20.76 | 69.92 | 13.51 | 108.93 | 45.78 | 10.04 | 2.95 | 34.69 | 65.7 | 611.19 | 113.69 | 248.18 | 103.34 | 1076.41 | 0.61 | 0.21 | 27.20 | 0.29 |
| | | | FL95-112 Wk3B1 | 7/27/2018 10:10 | Batch 3 | 80.5 | 3 | 5.12 | 16.21 | 14.52 | 306.9 | 3.87 | 192.45 | 93.52 | 12.88 | 59.44 | 11.33 | 76.78 | 26.34 | 2.83 | 2.48 | 18.59 | 42.65 | 533.95 | 106.4 | 176.72 | 63.72 | 880.79 | 1.84 | 1.65 | 34.84 | 0.44 |
| | | | FL95-113 Wk3B1 | 7/25/2018 1:59 | Batch 1 | 50.9 | 3 | 6.05 | 5.07 | 3.66 | 363.74 | 5.72 | 361.22 | 121.02 | 23.27 | 92.16 | 14.13 | 136.72 | 69.58 | 19.03 | 4.41 | 39.73 | 76.39 | 739.41 | 144.29 | 331.62 | 120.53 | 1335.85 | 0.38 | 0.27 | 27.23 | 0.43 |
| | | | FL95-135 Wk3B1 | 7/26/2018 20:07 | Batch 3 | 46.0 | 3 | 4.95 | 15.17 | 2.61 | 289.9 | 4.73 | 205.87 | 98.63 | 16.01 | 49.87 | 9.61 | 85 | 36.71 | 8.68 | 2.14 | 32.35 | 42.38 | 518.28 | 114.64 | 189.87 | 76.87 | 899.66 | 1.69 | 0.29 | 32.22 | 0.53 |
| | | | FL95-137 Wk3B1 | 7/25/2018 17:03 | Batch 1 | 48.9 | 3 | 4.23 | 0.38 | 3.42 | 383.41 | 4.21 | 293.95 | 122.09 | 24.85 | 70.49 | 17.3 | 97.08 | 49.6 | 14.59 | 2.39 | 34.9 | 46.86 | 685.37 | 146.94 | 249.06 | 84.15 | 1165.52 | 0.03 | 0.29 | 32.90 | 0.36 |
| | | | FL95-139 Wk3B1 | 7/26/2018 8:16 | Batch 2 | 43.3 | 3 | 5.74 | 7.44 | 2.7 | 163.21 | 1.62 | 95.9 | 53.45 | 7.21 | 41.33 | 11.25 | 50.8 | 24.65 | 3.09 | 3.05 | 20.3 | 26.34 | 270.87 | 60.66 | 131.12 | 49.69 | 512.34 | 1.45 | 0.53 | 31.86 | 0.32 |
| | | | FL95-144 Wk3B1 | 7/25/2018 2:44 | Batch 1 | 51.6 | 3 | 8.03 | 10.44 | 5.42 | 340.78 | 5.5 | 311.71 | 121.26 | 28 | 72.58 | 18.82 | 94.67 | 43.54 | 10.58 | 5.1 | 33.8 | 90.66 | 673.85 | 149.26 | 240.19 | 129.56 | 1192.86 | 0.88 | 0.45 | 28.57 | 0.46 |
| | | | FL95-151 Wk3B1 | 7/27/2018 5:01 | Batch 3 | 70.9 | 3 | 5.96 | 17.85 | 3.08 | 309.36 | 4.1 | 178.89 | 106.21 | 17.47 | 47.83 | 12.49 | 90.28 | 38.76 | 7.18 | 2.74 | 17.74 | 51.24 | 513.28 | 123.68 | 196.54 | 71.72 | 905.22 | 1.97 | 0.34 | 34.18 | 0.45 |
| | | | FL95-154 Wk3B1 | 7/26/2018 14:56 | Batch 3 | 49.2 | 3 | 5.74 | 22.19 | 4.71 | 342.84 | 6.61 | 442.08 | 120.3 | 23.51 | 74.14 | 13.34 | 122.33 | 46.87 | 9.72 | 6.11 | 37.63 | 70.95 | 818.43 | 143.81 | 266.4 | 114.69 | 1343.33 | 1.65 | 0.35 | 25.52 | 0.49 |
| | | | FL95-168 Wk3B1 | 7/26/2018 7:54 | Batch 2 | 55.3 | 3 | 8.40 | 11.99 | 2.96 | 310.95 | 3.56 | 229.33 | 91.12 | 14.85 | 66.87 | 12.85 | 74.34 | 23.24 | 6.18 | 7.63 | 41.41 | 73.99 | 558.79 | 105.97 | 183.48 | 123.03 | 971.27 | 1.23 | 0.30 | 32.01 | 0.37 |
| | | | FL95-170 Wk3B1 | 7/27/2018 7:35 | Batch 3 | 49.1 | 3 | 5.15 | 8.8 | 3.52 | 265.5 | 1.08 | 110.13 | 75.12 | 15.06 | 39.49 | 8.65 | 55.91 | 33.65 | 1.86 | 2.79 | 13.27 | 31.5 | 389.03 | 90.18 | 139.56 | 47.56 | 666.33 | 1.32 | 0.53 | 39.85 | 0.16 |
| | | | FL95-172 Wk3B1 | 7/25/2018 14:29 | Batch 1 | 49.5 | 3 | 6.79 | 0.38 | 3.27 | 321.11 | 3.79 | 311.81 | 90.71 | 16.26 | 57.74 | 9.24 | 93.99 | 41.28 | 10.12 | 3.83 | 28.69 | 68.21 | 640.36 | 106.97 | 212.37 | 100.73 | 1060.43 | 0.04 | 0.31 | 30.28 | 0.36 |
| | | | FL95-175 Wk3B1 | 7/26/2018 10:28 | Batch 2 | 46.8 | 3 | 6.50 | 3.26 | 2.57 | 188.76 | 2.25 | 151.2 | 70.48 | 14.04 | 58.51 | 9.69 | 64.83 | 19.87 | 4.07 | 3.13 | 18.27 | 39.12 | 348.04 | 84.52 | 156.97 | 60.52 | 650.05 | 0.50 | 0.40 | 29.04 | 0.35 |
| | | | FL95-180 Wk3B1 | 7/25/2018 12:16 | Batch 1 | 43.2 | 3 | 7.80 | 8.84 | 3 | 339.15 | 4.76 | 296.27 | 144.55 | 31.74 | 82.53 | 15.99 | 138.43 | 64.55 | 18.52 | 5.34 | 42.07 | 95.42 | 652.02 | 176.29 | 320.02 | 142.83 | 1291.15 | 0.68 | 0.23 | 26.27 | 0.37 |
| | | | FL95-182 Wk3B1 | 7/25/2018 5:19 | Batch 1 | 43.1 | 3 | 5.48 | 4.62 | 1.54 | 144.47 | 1.59 | 122.24 | 31.87 | 4.61 | 24.78 | 7.76 | 40.64 | 17.97 | 5.23 | 1.41 | 13.56 | 23 | 274.46 | 36.48 | 96.38 | 37.97 | 445.29 | 1.04 | 0.35 | 32.44 | 0.36 |
| | | | FL95-183 Wk3B1 | 7/25/2018 21:54 | Batch 2 | 50.0 | 3 | 6.17 | 17.66 | 4.14 | 403.26 | 6.53 | 365.33 | 136.9 | 29.66 | 82.19 | 13 | 112.77 | 50 | 7.57 | 6.52 | 43.9 | 77.12 | 796.92 | 166.56 | 265.53 | 127.54 | 1356.55 | 1.30 | 0.31 | 29.73 | 0.48 |
| | | | FL95-186 Wk3B1 | 7/26/2018 9:44 | Batch 2 | 30.8 | 3 | 4.96 | 14.7 | 3.87 | 268.54 | 1.72 | 323.11 | 88.12 | 17.83 | 49.99 | 12.09 | 102.03 | 43.18 | 11.04 | 3.46 | 30.5 | 46.95 | 611.94 | 105.95 | 218.33 | 80.91 | 1017.13 | 1.45 | 0.38 | 26.40 | 0.17 |
| | | | FL95-189 Wk3B1 | 7/25/2018 16:19 | Batch 1 | 55.5 | 3 | 4.44 | 9.62 | 1.29 | 188.31 | 2.11 | 236.11 | 71.86 | 21.13 | 35.68 | 8.03 | 54.27 | 24.57 | 10.4 | 1.35 | 15.47 | 30.21 | 437.44 | 92.99 | 132.95 | 47.03 | 710.42 | 1.35 | 0.18 | 26.51 | 0.30 |
| | | | FL95-194 Wk3B1 | 7/26/2018 6:25 | Batch 2 | 47.9 | 3 | 5.51 | 5.86 | 2.95 | 317.38 | 3.85 | 254.72 | 139.02 | 25.46 | 57.38 | 11.39 | 129.29 | 52.69 | 9.19 | 6.47 | 41.84 | 54.78 | 584.76 | 164.48 | 259.94 | 103.09 | 1112.27 | 0.53 | 0.27 | 28.53 | 0.35 |
| | | | FL95-198 Wk3B1 | 7/25/2018 21:09 | Batch 2 | 55.5 | 3 | 6.55 | 2.3 | 1.92 | 151.94 | 0.69 | 161.84 | 64.59 | 13.37 | 31.83 | 9.71 | 52.83 | 28.17 | 6.89 | 2.86 | 19.17 | 35.38 | 318.69 | 77.96 | 129.43 | 57.41 | 583.49 | 0.39 | 0.33 | 26.04 | 0.12 |
| | | | FL95-232 Wk3B1 | 7/25/2018 3:06 | Batch 1 | 40.2 | 3 | 8.06 | 15.17 | 8.29 | 466.43 | 7.47 | 367.23 | 196.76 | 38.86 | 117.74 | 20.34 | 176.04 | 61.87 | 14.32 | 7.67 | 50.33 | 127.34 | 864.59 | 235.62 | 390.31 | 185.34 | 1675.86 | 0.91 | 0.49 | 27.83 | 0.45 |
| | | | FL95-234 Wk3B1 | 7/24/2018 23:22 | Batch 1 | 40.5 | 3 | 4.39 | 0.31 | 2.11 | 244.21 | 4.65 | 194.58 | 75.98 | 15.2 | 49.95 | 18.7 | 75.02 | 47.11 | 9.24 | 3.4 | 28.09 | 31.73 | 445.86 | 91.18 | 200.02 | 63.22 | 800.28 | 0.04 | 0.26 | 30.52 | 0.58 |
| | | | FL95-239 Wk3B1 | 7/25/2018 15:57 | Batch 1 | 35.2 | 3 | 6.19 | 2.91 | 3.22 | 335.11 | 4.45 | 394.85 | 119.02 | 19.11 | 68.72 | 14.33 | 132.75 | 52.95 | 12.73 | 5.71 | 42.09 | 73.68 | 740.54 | 138.13 | 281.48 | 121.48 | 1281.63 | 0.23 | 0.25 | 26.15 | 0.35 |
| | | | FL95-243 Wk3B1 | 7/25/2018 18:55 | Batch 2 | 42.1 | 3 | 7.17 | 9.52 | 5.58 | 509.4 | 6.93 | 502.13 | 219.94 | 34.64 | 110.3 | 22.12 | 202.23 | 68.55 | 14.53 | 14.71 | 68.68 | 122.33 | 1033.56 | 254.58 | 417.73 | 205.72 | 1911.59 | 0.50 | 0.29 | 26.65 | 0.36 |
| | | | FL95-244 Wk3B1 | 7/26/2018 4:56 | Batch 2 | 39.2 | 3 | 6.10 | 3.75 | 2.55 | 121.6 | 1.56 | 93.77 | 51.16 | 7.41 | 27.91 | 10.37 | 39.47 | 27.88 | 4.57 | 1.68 | 19.57 | 25.08 | 223.23 | 58.57 | 110.2 | 46.33 | 438.33 | 0.86 | 0.58 | 27.74 | 0.36 |
| | | | FL95-245 Wk3B1 | 7/26/2018 22:22 | Batch 3 | 53.2 | 3 | 6.08 | 15.86 | 5.04 | 500.87 | 6.61 | 346.51 | 178.33 | 35.77 | 109.79 | 19.59 | 198.88 | 78.14 | 14.73 | 7.26 | 50.85 | 93.72 | 874.89 | 214.1 | 421.13 | 151.83 | 1661.95 | 0.95 | 0.30 | 30.14 | 0.40 |
| | | | FL95-258 Wk3B1 | 7/26/2018 3:49 | Batch 2 | 48.2 | 3 | 5.49 | 25.9 | 4.57 | 470.33 | 8.73 | 425.86 | 220.73 | 44.09 | 110.02 | 26.59 | 204.13 | 68.92 | 18.08 | 8.15 | 60.51 | 89.94 | 935.39 | 264.82 | 427.74 | 158.6 | 1786.55 | 1.45 | 0.26 | 26.33 | 0.49 |
| 0.505976096 | 2669.79 | 1.246 | FL95-003 Wk9B1 | 7/26/2018 21:15 | Batch 3 | 46.9 | 9 | 5.81 | 15.14 | 0.87 | 153.77 | 3.69 | 63.3 | 59.62 | 10.32 | 36.58 | 12.22 | 52.33 | 22.74 | 3.56 | 2.09 | 10.23 | 25.32 | 236.77 | 69.94 | 127.43 | 37.64 | 471.78 | 3.21 | 0.18 | 32.59 | 0.78 |
| 0.378570349 | 1847.53 | 1.008 | FL95-009 Wk9B1 | 7/25/2018 4:35 | Batch 1 | 44.3 | 9 | 6.89 | 10.26 | 3.21 | 280.55 | 3.77 | 275.93 | 72.97 | 17.89 | 46.04 | 13.82 | 84.51 | 40.62 | 6.07 | 3.17 | 25.28 | 62.02 | 573.72 | 90.86 | 191.06 | 90.47 | 946.11 | 1.08 | 0.34 | 29.65 | 0.40 |
| 0.363744389 | 2286.39 | 1.242 | FL95-020 Wk9B1 | 7/26/2018 10:50 | Batch 2 | 43.5 | 9 | 4.64 | 28.49 | 4.31 | 362.04 | 5.28 | 262.18 | 140.33 | 24.48 | 85.57 | 16.66 | 125.11 | 44.98 | 10.68 | 3.03 | 44.7 | 53.12 | 662.3 | 164.81 | 283 | 100.85 | 1210.96 | 2.35 | 0.36 | 29.90 | 0.44 |
| 0.56938292 | 2763.21 | 1.239 | FL95-032 Wk9B1 | 7/26/2018 7:31 | Batch 2 | 47.5 | 9 | 6.45 | 10.93 | 3.93 | 590.13 | 5.05 | 392.2 | 232.38 | 35.21 | 98.9 | 27.71 | 192.78 | 74.23 | 19.28 | 5.67 | 46.84 | 113.63 | 1002.24 | 267.59 | 412.9 | 166.14 | 1848.87 | 0.59 | 0.21 | 31.92 | 0.27 |
| 0.40017159 | 2563.88 | 1.329 | | | | | | | | | | | | | | | | | | | | | | | | | | | | | | |
| 0.630936529 | 1790.14 | 1.014 | FL95-052 Wk9B1 | 7/26/2018 22:45 | Batch 3 | 44.7 | 9 | 5.85 | 21.82 | 1.97 | 333.77 | 4.95 | 185.66 | 108.44 | 17.37 | 67.93 | 15.69 | 97.07 | 39.09 | 6.52 | 3.41 | 30.21 | 54.37 | 548.17 | 125.81 | 226.3 | 87.99 | 988.27 | 2.21 | 0.20 | 33.77 | 0.50 |
| 0.499699158 | 1900.94 | 1.075 | FL95-053 Wk9B1 | 7/25/2018 1:37 | Batch 1 | 28.7 | 9 | 5.53 | 7.03 | 5.38 | 532.01 | 8.15 | 422.83 | 230.86 | 46.49 | 144.46 | 26.55 | 253.48 | 102.81 | 23.09 | 8.02 | 90.07 | 102.76 | 975.4 | 277.35 | 550.39 | 200.85 | 2003.99 | 0.35 | 0.27 | 26.55 | 0.41 |
| 0.65835197 | 1893.81 | 1.118 | FL95-069 Wk9B1 | 7/26/2018 2:43 | Batch 2 | 45.1 | 9 | 5.08 | 16.8 | 5.59 | 373.72 | 6.84 | 342.55 | 142.54 | 27.05 | 70.57 | 12.14 | 152.8 | 57.54 | 13.2 | 3.48 | 38.09 | 63.96 | 745.5 | 169.59 | 306.25 | 105.53 | 1326.87 | 1.27 | 0.42 | 28.17 | 0.52 |
| 0.490145149 | 2338.7 | 1.175 | FL95-072 Wk9B1 | 7/24/2018 23:45 | Batch 1 | 31.8 | 9 | 8.03 | 12.88 | 3.93 | 508.27 | 6.25 | 426.96 | 186.39 | 47.05 | 77.26 | 18.45 | 132.2 | 72.73 | 22.63 | 4.33 | 49.68 | 132.22 | 958.29 | 233.44 | 323.27 | 186.23 | 1701.23 | 0.76 | 0.23 | 29.88 | 0.37 |
| 0.46211517 | 3075.07 | 1.435 | FL95-079 Wk9B1 | 7/25/2018 17:48 | Batch 2 | 51.5 | 9 | 8.90 | 0.54 | 1.58 | 163.93 | 2.71 | 212.28 | 52.36 | 7.99 | 40.25 | 7.73 | 46.17 | 21.56 | 3.39 | 3.16 | 19.99 | 53.58 | 381.04 | 60.35 | 119.1 | 76.73 | 637.22 | 0.08 | 0.25 | 25.73 | 0.43 |
| 0.557990868 | 2412.76 | 1.2 | FL95-084 Wk9B1 | 7/26/2018 17:31 | Batch 3 | 25.9 | 9 | 8.78 | 1.71 | 2.37 | 242.27 | 4.53 | 148.51 | 70.26 | 12.66 | 39.87 | 14.16 | 58.95 | 28.9 | 6.4 | 3.12 | 17.25 | 59.21 | 399.39 | 82.92 | 148.28 | 79.58 | 710.17 | 0.24 | 0.33 | 34.11 | 0.64 |
| 0.45785777 | 2538.5 | 1.324 | FL95-085 Wk9B1 | 7/25/2018 0:07 | Batch 1 | 42.8 | 9 | 4.93 | 4.72 | 6.39 | 354.08 | 6.36 | 378.46 | 129.36 | 24.44 | 70.67 | 15.32 | 151.82 | 64.27 | 16.05 | 4.2 | 63.27 | 62.45 | 750.01 | 153.8 | 318.13 | 129.92 | 1351.86 | 0.35 | 0.47 | 26.19 | 0.47 |
| 0.505825456 | 2561.45 | 1.301 | FL95-087 Wk9B1 | 7/25/2018 3:28 | Batch 1 | 46.4 | 9 | 5.72 | 10.59 | 3.25 | 304.08 | 5.94 | 288.7 | 97.6 | 17.92 | 68.79 | 12.74 | 127.03 | 52.27 | 8.61 | 3.44 | 41.86 | 59.61 | 612.56 | 115.52 | 269.44 | 104.91 | 1102.43 | 0.96 | 0.29 | 27.58 | 0.54 |
| 0.493163828 | 1737.77 | 1.018 | FL95-096 Wk9B1 | 7/26/2018 12:19 | Batch 2 | 44.7 | 9 | 8.77 | 8.76 | 3.95 | 239.87 | 3.29 | 180.58 | 107.51 | 22.27 | 56.56 | 8.32 | 77.28 | 32.44 | 8.22 | 4.72 | 26.14 | 69.75 | 436.45 | 129.78 | 182.82 | 100.61 | 849.66 | 1.03 | 0.46 | 28.23 | 0.39 |
| 0.553765107 | 1352.79 | 0.913 | FL95-097 Wk9B1 | 7/26/2018 3:05 | Batch 2 | 48.0 | 9 | 8.00 | 8.03 | 1.85 | 137.88 | 1.96 | 124.09 | 55.64 | 9.62 | 30.67 | 10.68 | 40.42 | 21.56 | 3.67 | 2.54 | 15.74 | 37.6 | 273.81 | 65.26 | 107 | 55.88 | 501.95 | 1.60 | 0.37 | 27.47 | 0.39 |
| 0.542313117 | 2109.27 | 1.131 | FL95-102 Wk9B1 | 7/24/2018 23:00 | Batch 1 | 48.9 | 9 | 8.82 | 7.07 | 3.05 | 385.2 | 6.95 | 331.24 | 166.34 | 28.69 | 89.41 | 18.68 | 147.48 | 53.3 | 12.41 | 9.98 | 66.08 | 117.34 | 733.51 | 195.03 | 321.28 | 193.4 | 1443.22 | 0.49 | 0.21 | 26.69 | 0.48 |
| 0.42712192 | 2420.25 | 1.169 | FL95-107 Wk9B1 | 7/25/2018 14:51 | Batch 1 | 43.5 | 9 | 8.12 | 6.08 | 2.71 | 281.53 | 3.93 | 275.19 | 88.42 | 14.35 | 45.39 | 9.41 | 97.02 | 42.81 | 6.96 | 4.44 | 31.81 | 75.55 | 569.44 | 102.77 | 201.59 | 111.8 | 985.61 | 0.62 | 0.27 | 28.56 | 0.40 |
| 0.507571043 | 1747.88 | 1.042 | FL95-109 Wk9B1 | 7/27/2018 6:07 | Batch 3 | 63.5 | 9 | 5.86 | 17.29 | 1.41 | 248.04 | 2.5 | 123.27 | 79.1 | 15.2 | 45.56 | 9.71 | 67.75 | 28.07 | 3.62 | 1.66 | 12.99 | 39.07 | 392.51 | 94.3 | 154.71 | 53.72 | 695.24 | 2.49 | 0.20 | 35.68 | 0.36 |
| 0.532924721 | 2090.1 | 1.121 | FL95-112 Wk9B1 | 7/26/2018 19:00 | Batch 3 | 42.9 | 9 | 6.31 | 23.58 | 2.82 | 288.17 | 5.5 | 169.42 | 83.27 | 16.09 | 59.58 | 12.18 | 75.86 | 34.11 | 3.88 | 2.97 | 14.28 | 50.19 | 489.49 | 99.36 | 185.61 | 67.44 | 841.9 | 2.80 | 0.33 | 34.23 | 0.65 |
| 0.482350954 | 2715.84 | 1.207 | FL95-113 Wk9B1 | 7/26/2018 17:09 | Batch 3 | 55.5 | 9 | 7.50 | 20.71 | 3.11 | 339.61 | 6.51 | 257.41 | 123.48 | 26.48 | 84.49 | 13.28 | 124.14 | 55.02 | 7.58 | 5.2 | 26.41 | 83.06 | 627.35 | 149.96 | 284.51 | 114.67 | 1176.49 | 1.76 | 0.26 | 28.87 | 0.55 |
| 0.763646702 | 2136.71 | 1.045 | FL95-135 Wk9B1 | 7/27/2018 4:16 | Batch 3 | 62.7 | 9 | 5.26 | 28.01 | 2.48 | 268.85 | 5.22 | 187.86 | 112.43 | 17 | 53.47 | 10.89 | 102.06 | 44.85 | 5.55 | 3.71 | 30.23 | 44.49 | 492.42 | 129.43 | 216.82 | 78.43 | 917.1 | 3.05 | 0.27 | 29.32 | 0.57 |
| 0.63947953 | 2386.48 | 1.1 | FL95-137 Wk9B1 | 7/25/2018 1:14 | Batch 1 | 30.8 | 9 | 4.68 | 6.92 | 4.49 | 468.22 | 7.44 | 479.36 | 205.87 | 34.92 | 123.31 | 28.43 | 171.31 | 90.07 | 20.86 | 5.09 | 70.57 | 78.99 | 966.43 | 240.79 | 433.98 | 154.65 | 1795.85 | 0.39 | 0.25 | 26.07 | 0.41 |
| 0.638649751 | 1764.86 | 1.033 | FL95-139 Wk9B1 | 7/27/2018 9:04 | Batch 3 | 50.7 | 9 | 6.16 | 14.79 | 3.93 | 369.72 | 5.11 | 235.34 | 115.34 | 20.1 | 81.5 | 17.53 | 110.03 | 38.28 | 5.83 | 5.39 | 30.97 | 63.47 | 628.89 | 135.44 | 253.17 | 99.83 | 1117.33 | 1.32 | 0.35 | 33.09 | 0.46 |
| 0.491791908 | 3156.53 | 1.384 | FL95-144 Wk9B1 | 7/26/2018 16:47 | Batch 3 | 45.0 | 9 | 6.81 | 37.41 | 4.54 | 533.24 | 6.71 | 423.11 | 202.59 | 37.92 | 122.39 | 25.56 | 137.08 | 53.61 | 14.8 | 5.66 | 33.43 | 113.68 | 1005.01 | 240.51 | 353.44 | 152.77 | 1751.73 | 2.14 | 0.26 | 30.44 | 0.38 |
| 0.527201632 | 2335 | 1.144 | FL95-151 Wk9B1 | 7/25/2018 16:41 | Batch 1 | 52.8 | 9 | 5.29 | 5.41 | 2.31 | 336.08 | 4.2 | 278.69 | 101.78 | 21.21 | 53.68 | 13.68 | 97.96 | 42.09 | 10.88 | 2.87 | 27.15 | 52.75 | 626.69 | 122.99 | 218.29 | 82.77 | 1050.74 | 0.51 | 0.22 | 31.99 | 0.40 |
| 0.401176635 | 2029.16 | 1.009 | FL95-154 Wk9B1 | 7/26/2018 18:38 | Batch 3 | 70.4 | 9 | 6.24 | 16.19 | 4.46 | 455.92 | 5.73 | 307.54 | 138.06 | 26 | 74.92 | 12.87 | 105.76 | 34.2 | 6.57 | 6.83 | 34.79 | 74.58 | 789.84 | 164.06 | 234.32 | 116.2 | 1304.42 | 1.24 | 0.34 | 34.95 | 0.44 |
| 0.270276954 | 2128.37 | 1.056 | FL95-168 Wk9B1 | 7/27/2018 8:20 | Batch 3 | 68.3 | 9 | 6.72 | 24.52 | 2.52 | 261.93 | 3.41 | 182.14 | 68.45 | 13.05 | 43.03 | 7.77 | 50.23 | 16.12 | 0.87 | 2.92 | 16.01 | 46.77 | 474.52 | 81.5 | 118.02 | 65.7 | 739.74 | 3.31 | 0.34 | 35.41 | 0.46 |
| 0.532020902 | 2258.86 | 1.14 | FL95-170 Wk9B1 | 7/25/2018 15:35 | Batch 1 | 55.1 | 9 | 5.58 | 9.08 | 4.18 | 445.82 | 5.24 | 415.59 | 175.08 | 30.85 | 110.8 | 17.54 | 193.68 | 83.03 | 15.38 | 5.57 | 60.07 | 86.98 | 879.91 | 205.93 | 420.43 | 152.62 | 1658.89 | 0.55 | 0.25 | 26.87 | 0.32 |
| 0.333544144 | 2256.18 | 1.092 | FL95-172 Wk9B1 | 7/25/2018 4:57 | Batch 1 | 47.0 | 9 | 9.39 | 13.99 | 5.33 | 462.05 | 6.63 | 372.54 | 168.89 | 36.99 | 110.3 | 17.69 | 201.58 | 71.08 | 14.03 | 9.97 | 57.49 | 149.56 | 860.54 | 205.88 | 414.68 | 217.02 | 1698.12 | 0.82 | 0.31 | 27.21 | 0.39 |
| 0.491361694 | 2114.96 | 1.129 | FL95-175 Wk9B1 | 7/26/2018 16:25 | Batch 3 | 72.0 | 9 | 5.81 | 13.99 | 2.4 | 226.29 | 2.5 | 154.69 | 68.32 | 11.22 | 54.49 | 8.04 | 59.81 | 20.99 | 4.72 | 2.73 | 14.56 | 36.88 | 399.87 | 79.54 | 148.05 | 54.17 | 681.63 | 2.05 | 0.35 | 33.20 | 0.37 |
| 0.439213049 | 1850.43 | 0.978 | FL95-180 Wk9B1 | 7/25/2018 6:04 | Batch 1 | 36.4 | 9 | 5.87 | 5.5 | 1.68 | 157.54 | 2.76 | 166.78 | 44.11 | 8.35 | 28.15 | 7.52 | 49.13 | 25.95 | 7.08 | 1.29 | 17.82 | 31.3 | 334.26 | 52.46 | 117.83 | 50.41 | 554.95 | 0.99 | 0.30 | 28.39 | 0.50 |
| 0.534013605 | 2702.83 | 1.237 | FL95-182 Wk9B1 | 7/26/2018 20:52 | Batch 3 | 84.9 | 9 | 5.78 | 15.79 | 4.28 | 516.76 | 7.16 | 282.63 | 143.3 | 22.15 | 85.59 | 16.11 | 134.45 | 42.85 | 11.16 | 7.05 | 30.18 | 73.48 | 826.62 | 165.45 | 290.16 | 110.71 | 1392.94 | 1.13 | 0.31 | 37.10 | 0.51 |
| 0.56436125 | 2214.57 | 1.126 | FL95-183 Wk9B1 | 7/26/2018 5:19 | Batch 2 | 57.2 | 9 | 6.24 | 3.67 | 2.76 | 247.02 | 3.2 | 235.45 | 82.34 | 11.32 | 46.25 | 7.95 | 66.51 | 31.17 | 5.8 | 3.1 | 24.64 | 48.02 | 492.1 | 93.66 | 157.68 | 75.76 | 819.2 | 0.45 | 0.34 | 30.15 | 0.39 |
| 0.535024313 | 2240.01 | 1.131 | FL95-186 Wk9B1 | 7/25/2018 15:13 | Batch 1 | 49.7 | 9 | 5.61 | 7.15 | 1.8 | 199.6 | 1.99 | 188.29 | 79.77 | 15.34 | 40.16 | 8.13 | 80.75 | 42.64 | 6.76 | 2.77 | 23.34 | 38.59 | 398.83 | 95.11 | 178.44 | 64.7 | 737.08 | 0.97 | 0.24 | 27.08 | 0.27 |
| 0.68299653 | 1924.53 | 1.041 | FL95-189 Wk9B1 | 7/25/2018 22:17 | Batch 2 | 27.2 | 9 | 5.62 | 14.46 | 4.32 | 446.03 | 6.5 | 454.18 | 208.42 | 33.97 | 103.97 | 12.89 | 129.68 | 56.36 | 12.63 | 3.67 | 36.1 | 86.88 | 925.49 | 242.39 | 315.53 | 126.65 | 1610.06 | 0.90 | 0.27 | 27.70 | 0.40 |
| 0.707326284 | 1478.33 | 0.9 | FL95-194 Wk9B1 | 7/27/2018 6:51 | Batch 3 | 70.6 | 9 | 5.30 | 25.82 | 2.12 | 493.02 | 3.7 | 243.43 | 206.13 | 40.07 | 91.27 | 12.82 | 176.33 | 56.34 | 10.43 | 9.59 | 46.13 | 69.22 | 768.09 | 246.2 | 347.19 | 124.94 | 1486.42 | 1.74 | 0.14 | 33.17 | 0.25 |
| 0.569498296 | 2397.73 | 1.217 | FL95-198 Wk9B1 | 7/26/2018 17:53 | Batch 3 | 54.6 | 9 | 8.47 | 21.85 | 5.43 | 344.97 | 4 | 222.03 | 171.88 | 32.67 | 76.8 | 16.86 | 123.73 | 53.39 | 9.76 | 5.61 | 37.2 | 98.05 | 598.28 | 204.55 | 280.54 | 140.86 | 1224.23 | 1.78 | 0.44 | 28.18 | 0.33 |
| 0.525311203 | 1655.97 | 0.936 | FL95-232 Wk9B1 | 7/25/2018 12:38 | Batch 1 | 44.7 | 9 | 7.40 | 11.23 | 4.13 | 454.42 | 5.32 | 337.81 | 173.17 | 28.92 | 111.93 | 18.52 | 165.18 | 63.27 | 16.85 | 5.39 | 52.18 | 109.85 | 812.91 | 202.09 | 375.75 | 167.42 | 1558.17 | 0.72 | 0.27 | 29.16 | 0.34 |
| 1.030744624 | 1833.15 | 0.964 | FL95-234 Wk9B1 | 7/25/2018 20:47 | Batch 2 | 55.5 | 9 | 4.49 | 4.79 | 2.06 | 133.48 | 2.1 | 172.74 | 46.51 | 5.28 | 24.27 | 11.08 | 29.89 | 16.59 | 0.57 | 2.18 | 8.8 | 19.38 | 315.17 | 51.79 | 82.4 | 30.36 | 479.72 | 1.00 | 0.43 | 27.82 | 0.44 |
| 0.416860154 | 2727.86 | 1.336 | FL95-239 Wk9B1 | 7/25/2018 19:17 | Batch 2 | 46.9 | 9 | 6.81 | 8.13 | 1.2 | 130.66 | 1.94 | 150.31 | 56.9 | 12.81 | 30.63 | 8.97 | 59.41 | 26.5 | 4.85 | 3.21 | 14.98 | 33.85 | 292.24 | 69.71 | 130.36 | 52.04 | 544.35 | 1.49 | 0.22 | 24.00 | 0.36 |
| 0.436698386 | 2105.98 | 1.019 | FL95-243 Wk9B1 | 7/26/2018 6:03 | Batch 2 | 48.4 | 9 | 7.73 | 6.9 | 3.11 | 244.56 | 3.2 | 241.84 | 105.98 | 16.27 | 53.71 | 11.07 | 92.37 | 36.92 | 7.61 | 7.22 | 33.51 | 64.57 | 499.61 | 122.25 | 201.68 | 105.3 | 928.84 | 0.74 | 0.33 | 26.33 | 0.34 |
| 0.631948782 | 2531.23 | 1.237 | FL95-244 Wk9B1 | 7/25/2018 13:22 | Batch 1 | 50.8 | 9 | 7.32 | 6.33 | 2.27 | 247.76 | 2.77 | 215.78 | 86.73 | 17.54 | 49.87 | 14.78 | 76.79 | 40.24 | 7.79 | 5.03 | 30.02 | 58.09 | 474.91 | 104.27 | 189.47 | 93.14 | 861.79 | 0.73 | 0.26 | 28.75 | 0.32 |
| 0.538385549 | 2647.81 | 1.249 | FL95-245 Wk9B1 | 7/26/2018 6:47 | Batch 2 | 38.0 | 9 | 8.41 | 2.59 | 0.96 | 117.1 | 1.03 | 105.18 | 44.08 | 7.7 | 25.86 | 8.71 | 43.74 | 30.5 | 2.74 | 2.77 | 11.66 | 34.15 | 226.86 | 51.78 | 111.55 | 48.58 | 438.77 | 0.59 | 0.22 | 26.69 | 0.23 |
| 0.513258587 | 2101.21 | 1.098 | FL95-258 Wk9B1 | 7/26/2018 22:00 | Batch 3 | 66.1 | 9 | 6.67 | 15.5 | 3.67 | 306.29 | 4.38 | 244.98 | 111.74 | 20.01 | 53.74 | 14.32 | 77.23 | 30.7 | 2.32 | 4.77 | 26.76 | 60.34 | 574.82 | 131.75 | 178.31 | 91.87 | 976.75 | 1.59 | 0.38 | 31.36 | 0.45 |

| % C18:0 | % C18:1n9 | % C18:1n7 | % C18:2n6 | % C20:3n6 | % C20:4n6 | % C22:4n6 | % C22:5n6 | % C20:5n3 | % C22:5n3 | % C22:6n3 | Σ SFA % | Σ MUFA % | Σ (n-6) PUFA % | Σ (n-3) PUFA % |
|---|---|---|---|---|---|---|---|---|---|---|---|---|---|---|
| 28.67 | 10.24 | 2.10 | 6.93 | 1.63 | 9.11 | 4.06 | 0.70 | 0.48 | 3.23 | 4.47 | 57.06 | 12.33 | 22.43 | 8.18 |
| 21.59 | 10.63 | 2.12 | 7.04 | 1.53 | 9.80 | 4.55 | 0.90 | 0.28 | 2.40 | 4.95 | 55.79 | 12.76 | 23.83 | 7.63 |
| 17.49 | 12.75 | 2.62 | 7.07 | 1.18 | 9.41 | 3.55 | 0.73 | 0.33 | 3.20 | 3.74 | 55.43 | 15.36 | 21.93 | 7.27 |
| 25.49 | 11.47 | 1.98 | 5.14 | 1.63 | 8.44 | 4.24 | 0.77 | 0.32 | 2.76 | 5.38 | 57.87 | 13.45 | 20.23 | 8.46 |
| 31.14 | 9.93 | 1.48 | 4.87 | 1.24 | 7.18 | 4.19 | 0.85 | 0.26 | 2.33 | 4.15 | 63.53 | 11.41 | 18.33 | 6.73 |
| 19.15 | 11.83 | 2.48 | 7.34 | 1.49 | 9.41 | 3.16 | 0.64 | 0.43 | 2.72 | 5.82 | 54.68 | 14.31 | 22.04 | 8.97 |
| 17.44 | 12.61 | 1.77 | 7.81 | 0.99 | 12.17 | 3.76 | 0.95 | 0.55 | 3.36 | 4.00 | 52.04 | 14.37 | 25.68 | 7.91 |
| 29.01 | 8.96 | 1.86 | 5.12 | 1.07 | 9.72 | 4.54 | 1.40 | 0.30 | 2.88 | 4.52 | 59.63 | 10.82 | 21.86 | 7.69 |
| 19.07 | 13.60 | 2.86 | 5.40 | 1.14 | 7.08 | 3.59 | 1.04 | 0.25 | 1.86 | 6.19 | 56.99 | 16.47 | 18.25 | 8.30 |
| 31.43 | 8.76 | 1.90 | 7.41 | 1.06 | 8.00 | 3.74 | 0.85 | 0.28 | 2.61 | 7.25 | 58.14 | 10.66 | 21.06 | 10.14 |
| 22.47 | 9.96 | 2.19 | 6.87 | 1.72 | 8.64 | 4.15 | 1.22 | 0.36 | 2.65 | 5.27 | 56.96 | 12.15 | 22.60 | 8.28 |
| 26.21 | 10.09 | 1.81 | 6.47 | 1.45 | 11.23 | 4.98 | 1.17 | 0.52 | 5.33 | 4.43 | 52.53 | 11.90 | 25.30 | 10.28 |
| 16.49 | 11.78 | 2.53 | 6.70 | 1.04 | 11.52 | 3.83 | 0.62 | 0.40 | 2.29 | 5.44 | 53.86 | 14.31 | 23.70 | 8.13 |
| 26.68 | 9.33 | 1.43 | 5.41 | 1.05 | 9.00 | 4.72 | 0.94 | 0.62 | 3.12 | 7.36 | 57.03 | 10.77 | 21.12 | 11.09 |
| 26.77 | 9.97 | 2.21 | 6.69 | 1.67 | 7.33 | 3.91 | 0.53 | 0.34 | 2.56 | 4.97 | 59.81 | 12.18 | 20.13 | 7.88 |
| 26.93 | 10.95 | 1.59 | 6.46 | 1.22 | 9.48 | 3.99 | 1.04 | 0.61 | 4.46 | 6.79 | 53.42 | 12.53 | 22.18 | 11.86 |
| 35.59 | 8.42 | 1.92 | 6.79 | 1.10 | 11.47 | 5.57 | 0.49 | 0.37 | 3.69 | 6.37 | 53.80 | 10.35 | 25.42 | 10.43 |
| 28.95 | 8.55 | 2.02 | 5.90 | 1.18 | 9.51 | 4.67 | 1.12 | 0.30 | 2.83 | 6.44 | 57.50 | 10.57 | 22.37 | 9.56 |
| 22.67 | 10.24 | 1.53 | 7.56 | 1.42 | 9.20 | 4.90 | 0.71 | 0.31 | 2.86 | 5.48 | 55.80 | 11.76 | 23.79 | 8.65 |
| 28.80 | 9.52 | 1.61 | 7.71 | 1.13 | 9.66 | 4.92 | 1.15 | 0.34 | 2.33 | 5.28 | 56.35 | 11.13 | 24.57 | 7.95 |
| 25.19 | 10.73 | 2.34 | 5.76 | 1.10 | 9.81 | 4.34 | 1.02 | 0.32 | 3.79 | 5.17 | 55.62 | 13.07 | 22.03 | 9.28 |
| 26.68 | 12.37 | 1.98 | 6.48 | 1.36 | 8.98 | 4.19 | 1.05 | 0.25 | 2.69 | 3.67 | 56.98 | 14.34 | 22.07 | 6.60 |
| 23.98 | 8.98 | 1.33 | 7.21 | 1.31 | 8.35 | 3.62 | 0.64 | 0.35 | 2.45 | 3.24 | 62.51 | 10.32 | 21.14 | 6.04 |
| 17.88 | 13.51 | 3.02 | 6.38 | 1.37 | 7.67 | 2.87 | 0.32 | 0.51 | 1.64 | 7.15 | 55.58 | 16.53 | 18.60 | 9.30 |
| 24.51 | 9.33 | 2.60 | 6.34 | 1.19 | 9.40 | 3.39 | 0.97 | 0.38 | 2.60 | 4.89 | 58.91 | 11.93 | 21.29 | 7.87 |
| 25.15 | 9.41 | 2.35 | 7.53 | 1.37 | 6.90 | 1.73 | 0.07 | 1.04 | 4.30 | 8.02 | 57.27 | 11.76 | 17.60 | 13.36 |
| 14.67 | 12.48 | 2.62 | 7.22 | 1.37 | 11.33 | 5.99 | 1.03 | 0.35 | 4.33 | 5.69 | 47.57 | 15.10 | 26.95 | 10.38 |
| 37.03 | 8.47 | 1.13 | 5.27 | 1.03 | 8.64 | 4.31 | 0.71 | 0.31 | 3.11 | 6.73 | 60.29 | 9.60 | 19.96 | 10.16 |
| 23.38 | 10.58 | 1.99 | 8.16 | 1.43 | 10.22 | 3.87 | 0.83 | 0.64 | 3.78 | 6.01 | 52.49 | 12.56 | 24.51 | 10.43 |
| 27.12 | 13.09 | 2.02 | 6.25 | 1.14 | 8.80 | 4.29 | 0.89 | 0.38 | 2.29 | 6.79 | 54.05 | 15.11 | 21.37 | 9.47 |
| 31.66 | 9.41 | 1.43 | 6.56 | 1.44 | 9.47 | 3.78 | 0.88 | 0.40 | 2.92 | 5.50 | 58.22 | 10.84 | 22.12 | 8.82 |
| 33.63 | 7.89 | 1.30 | 5.09 | 0.88 | 7.37 | 3.55 | 0.55 | 0.43 | 2.80 | 4.61 | 65.52 | 9.20 | 17.44 | 7.85 |
| 25.93 | 11.25 | 1.91 | 5.77 | 1.33 | 10.96 | 4.83 | 1.02 | 0.32 | 3.01 | 5.34 | 54.25 | 13.16 | 23.91 | 8.68 |
| 25.33 | 13.61 | 1.84 | 6.97 | 1.09 | 9.83 | 4.32 | 1.40 | 0.16 | 2.77 | 4.08 | 53.94 | 15.44 | 23.62 | 7.00 |
| 25.26 | 13.06 | 1.85 | 5.28 | 1.10 | 11.28 | 4.70 | 1.14 | 0.51 | 3.69 | 4.00 | 53.41 | 14.90 | 23.50 | 8.19 |
| 22.90 | 12.29 | 1.95 | 5.61 | 1.32 | 7.26 | 3.34 | 0.50 | 0.29 | 1.89 | 3.73 | 61.82 | 14.24 | 18.04 | 5.90 |
| 23.50 | 11.55 | 2.08 | 7.04 | 1.06 | 10.09 | 3.50 | 0.91 | 0.38 | 3.02 | 7.66 | 52.72 | 13.63 | 22.60 | 11.06 |
| 16.99 | 13.06 | 2.74 | 7.49 | 2.34 | 9.30 | 4.71 | 0.63 | 0.38 | 2.08 | 3.51 | 53.76 | 15.80 | 24.47 | 5.97 |
| 28.06 | 8.26 | 1.70 | 5.46 | 1.25 | 10.07 | 4.55 | 0.95 | 0.56 | 3.17 | 6.31 | 57.73 | 9.96 | 22.28 | 10.04 |
| 23.96 | 11.38 | 1.54 | 5.58 | 1.29 | 9.64 | 3.54 | 0.63 | 0.66 | 3.53 | 6.37 | 55.85 | 12.91 | 20.68 | 10.56 |
| 30.66 | 10.34 | 1.95 | 5.65 | 2.02 | 8.02 | 4.67 | 1.04 | 0.35 | 3.37 | 3.94 | 58.65 | 12.29 | 21.40 | 7.66 |
| 21.87 | 11.72 | 2.63 | 6.26 | 1.20 | 12.18 | 4.83 | 1.24 | 0.38 | 2.94 | 4.93 | 51.68 | 14.35 | 25.71 | 8.25 |
| 22.41 | 13.32 | 2.00 | 5.74 | 1.90 | 10.28 | 4.17 | 1.06 | 0.49 | 3.92 | 4.66 | 52.45 | 15.33 | 23.15 | 9.08 |
| 25.12 | 10.36 | 2.32 | 7.44 | 1.88 | 10.98 | 4.58 | 0.57 | 0.49 | 3.79 | 5.58 | 52.00 | 12.68 | 25.45 | 9.87 |
| 35.72 | 9.07 | 1.41 | 5.26 | 1.02 | 7.87 | 3.09 | 0.72 | 0.18 | 1.83 | 3.75 | 65.80 | 10.48 | 17.96 | 5.76 |
| 21.65 | 10.55 | 1.94 | 6.74 | 1.42 | 11.94 | 5.01 | 1.29 | 0.31 | 4.65 | 5.08 | 51.07 | 12.49 | 26.41 | 10.03 |
| 23.53 | 12.14 | 1.87 | 5.37 | 1.66 | 9.08 | 4.52 | 0.81 | 0.35 | 2.27 | 5.46 | 56.46 | 14.01 | 21.45 | 8.08 |
| 21.67 | 11.12 | 1.79 | 5.65 | 1.35 | 9.01 | 4.36 | 0.90 | 0.38 | 2.79 | 4.61 | 58.04 | 12.91 | 21.27 | 7.78 |
| 20.50 | 10.81 | 1.82 | 6.46 | 1.41 | 8.73 | 3.48 | 0.95 | 0.32 | 2.95 | 5.27 | 57.81 | 12.63 | 21.03 | 8.53 |
| 13.32 | 15.13 | 3.15 | 7.96 | 1.23 | 11.60 | 4.12 | 0.56 | 0.40 | 3.07 | 4.56 | 48.24 | 18.27 | 25.46 | 8.03 |
| 26.96 | 9.95 | 1.91 | 5.70 | 0.87 | 12.32 | 5.42 | 1.30 | 0.24 | 3.81 | 4.99 | 53.48 | 11.86 | 25.61 | 9.05 |
| 14.41 | 14.55 | 4.13 | 4.54 | 1.23 | 7.63 | 3.41 | 0.98 | 0.30 | 1.59 | 6.58 | 55.04 | 18.69 | 17.80 | 8.47 |
| 30.32 | 8.62 | 1.79 | 7.57 | 1.11 | 7.90 | 3.47 | 1.07 | 0.35 | 2.94 | 6.80 | 58.37 | 10.41 | 21.12 | 10.09 |
| 23.52 | 10.46 | 1.78 | 6.28 | 2.08 | 8.30 | 4.49 | 1.00 | 0.34 | 2.19 | 5.46 | 57.61 | 12.24 | 22.15 | 8.00 |
| 22.20 | 10.71 | 2.24 | 6.69 | 1.32 | 11.80 | 4.71 | 1.34 | 0.41 | 4.71 | 4.62 | 51.44 | 12.95 | 25.87 | 9.74 |
| 25.45 | 11.60 | 1.85 | 6.94 | 0.80 | 11.59 | 4.06 | 0.77 | 0.28 | 3.12 | 5.52 | 53.46 | 13.46 | 24.15 | 8.92 |
| 22.90 | 11.13 | 2.11 | 6.35 | 1.02 | 10.87 | 4.43 | 1.17 | 0.51 | 3.33 | 7.32 | 51.78 | 13.24 | 23.83 | 11.16 |
| 23.19 | 12.50 | 2.47 | 7.12 | 1.32 | 8.71 | 3.59 | 0.95 | 0.25 | 2.96 | 4.87 | 55.25 | 14.97 | 21.69 | 8.08 |
| 23.23 | 11.92 | 2.33 | 5.17 | 1.37 | 7.86 | 2.99 | 0.34 | 0.66 | 3.10 | 5.48 | 58.79 | 14.25 | 17.73 | 9.23 |
| 15.47 | 10.54 | 1.39 | 4.69 | 1.14 | 8.29 | 3.58 | 0.55 | 0.55 | 1.61 | 4.94 | 62.73 | 11.93 | 18.24 | 7.10 |
| 28.48 | 8.63 | 1.93 | 6.50 | 1.26 | 10.12 | 4.25 | 0.93 | 0.27 | 3.22 | 6.10 | 56.78 | 10.56 | 23.06 | 9.60 |
| 21.85 | 10.62 | 1.46 | 6.75 | 1.29 | 8.72 | 2.99 | 0.32 | 0.28 | 2.11 | 4.84 | 60.62 | 12.08 | 20.06 | 7.23 |
| 27.04 | 9.06 | 1.74 | 6.90 | 1.06 | 10.23 | 5.21 | 1.42 | 0.33 | 2.97 | 5.72 | 55.35 | 10.80 | 24.82 | 9.02 |
| 22.88 | 10.96 | 1.78 | 5.54 | 1.07 | 9.45 | 4.08 | 0.96 | 0.24 | 3.60 | 4.71 | 57.61 | 12.74 | 21.10 | 8.54 |
| 25.22 | 10.48 | 2.13 | 6.05 | 1.48 | 8.33 | 4.26 | 1.25 | 0.21 | 2.99 | 4.02 | 58.80 | 12.61 | 21.37 | 7.22 |
| 18.72 | 10.43 | 1.41 | 8.07 | 2.20 | 9.92 | 4.81 | 0.60 | 0.60 | 3.96 | 5.14 | 52.87 | 11.84 | 25.59 | 9.70 |
| 26.13 | 10.17 | 2.35 | 6.08 | 1.58 | 7.94 | 3.65 | 0.89 | 0.43 | 2.83 | 7.60 | 56.49 | 12.51 | 20.14 | 10.86 |
| 19.76 | 11.73 | 1.93 | 5.28 | 1.38 | 9.97 | 4.28 | 0.79 | 0.30 | 1.96 | 5.66 | 56.70 | 13.66 | 21.71 | 7.92 |
| 32.91 | 8.96 | 1.75 | 5.52 | 0.99 | 9.11 | 3.49 | 0.72 | 0.45 | 2.80 | 5.28 | 60.93 | 10.71 | 19.83 | 8.54 |
| 23.61 | 9.38 | 1.53 | 6.88 | 1.32 | 7.65 | 2.39 | 0.64 | 0.79 | 4.26 | 7.62 | 57.53 | 10.91 | 18.89 | 12.67 |
| 16.53 | 11.27 | 2.26 | 5.93 | 1.30 | 8.39 | 5.05 | 0.28 | 0.42 | 1.99 | 4.73 | 58.38 | 13.53 | 20.94 | 7.14 |
| 29.40 | 8.55 | 1.53 | 5.44 | 0.87 | 8.86 | 3.89 | 0.95 | 0.36 | 2.71 | 6.43 | 60.39 | 10.09 | 20.03 | 9.50 |
| 23.26 | 10.84 | 2.16 | 9.00 | 1.49 | 9.97 | 3.06 | 0.63 | 0.48 | 2.81 | 6.02 | 53.54 | 13.00 | 24.15 | 9.31 |
| 22.95 | 11.20 | 2.46 | 6.39 | 1.24 | 10.72 | 5.00 | 1.43 | 0.41 | 3.26 | 7.39 | 50.50 | 13.65 | 24.79 | 11.06 |
| 27.45 | 7.16 | 1.04 | 5.56 | 1.74 | 9.13 | 4.04 | 1.17 | 0.32 | 3.05 | 5.16 | 61.64 | 8.19 | 21.64 | 8.53 |
| 26.93 | 10.09 | 2.19 | 6.06 | 0.96 | 8.31 | 3.69 | 0.56 | 0.48 | 3.24 | 5.69 | 58.75 | 12.28 | 19.57 | 9.40 |
| 31.77 | 8.66 | 1.75 | 4.91 | 1.19 | 10.03 | 4.25 | 1.09 | 0.34 | 3.00 | 4.62 | 60.16 | 10.42 | 21.47 | 7.95 |
| 33.24 | 10.12 | 2.97 | 5.02 | 1.13 | 7.64 | 3.46 | 1.46 | 0.19 | 2.18 | 4.25 | 61.58 | 13.09 | 18.71 | 6.62 |
| 22.90 | 12.50 | 2.29 | 5.16 | 1.02 | 11.62 | 4.74 | 0.83 | 0.58 | 3.76 | 4.93 | 52.57 | 14.79 | 23.37 | 9.27 |
| 27.74 | 11.07 | 2.29 | 5.46 | 1.66 | 9.05 | 4.83 | 1.18 | 0.49 | 3.29 | 6.06 | 54.62 | 13.36 | 22.18 | 9.84 |
| 21.91 | 11.74 | 2.32 | 7.03 | 1.21 | 10.50 | 3.69 | 0.85 | 0.46 | 3.00 | 7.60 | 51.59 | 14.06 | 23.29 | 11.06 |
| 24.31 | 9.49 | 1.90 | 6.24 | 2.34 | 9.37 | 5.89 | 1.15 | 0.42 | 3.51 | 3.96 | 55.71 | 11.39 | 24.99 | 7.90 |
| 30.81 | 9.29 | 1.49 | 5.36 | 1.12 | 10.36 | 4.13 | 0.99 | 0.45 | 3.28 | 5.75 | 57.78 | 10.78 | 21.96 | 9.48 |
| 26.27 | 11.51 | 1.81 | 5.77 | 1.16 | 10.58 | 3.59 | 0.76 | 0.77 | 3.59 | 6.40 | 54.07 | 13.32 | 21.85 | 10.76 |
| 21.39 | 11.67 | 1.69 | 6.37 | 2.37 | 9.00 | 6.36 | 1.04 | 0.38 | 4.46 | 5.72 | 50.93 | 13.36 | 25.14 | 10.57 |
| 20.85 | 10.73 | 2.15 | 6.61 | 1.18 | 11.97 | 4.70 | 0.89 | 0.44 | 3.06 | 5.64 | 52.64 | 12.88 | 25.34 | 9.14 |
| 23.84 | 12.36 | 2.47 | 6.16 | 1.49 | 11.43 | 3.86 | 1.01 | 0.46 | 3.39 | 5.03 | 52.36 | 14.82 | 23.94 | 8.88 |
| 13.42 | 12.64 | 2.19 | 7.75 | 2.59 | 11.09 | 4.82 | 0.75 | 0.44 | 2.17 | 5.37 | 50.19 | 14.82 | 27.01 | 7.98 |
| 29.16 | 7.71 | 1.89 | 4.87 | 1.46 | 8.93 | 4.29 | 0.64 | 0.34 | 2.67 | 6.56 | 60.64 | 9.60 | 20.19 | 9.56 |
| 21.65 | 11.59 | 2.02 | 7.07 | 1.38 | 10.33 | 3.71 | 0.88 | 0.25 | 3.69 | 4.39 | 54.69 | 13.61 | 23.37 | 8.33 |
| 21.21 | 12.57 | 1.90 | 5.35 | 1.50 | 10.43 | 4.01 | 1.04 | 0.31 | 2.53 | 6.15 | 54.21 | 14.47 | 22.33 | 8.99 |
| 18.79 | 10.97 | 1.76 | 6.87 | 1.59 | 9.82 | 3.96 | 0.66 | 0.35 | 3.06 | 5.50 | 55.47 | 12.73 | 22.90 | 8.90 |
| 21.10 | 11.52 | 2.32 | 7.21 | 1.32 | 12.65 | 5.13 | 1.15 | 0.40 | 4.49 | 5.13 | 48.67 | 13.84 | 27.46 | 10.02 |
| 25.82 | 10.74 | 2.04 | 5.32 | 0.91 | 11.52 | 4.34 | 0.99 | 0.26 | 2.87 | 4.82 | 56.18 | 12.78 | 23.08 | 7.95 |
| 25.10 | 10.96 | 2.77 | 4.54 | 1.08 | 7.77 | 4.28 | 1.33 | 0.25 | 2.92 | 7.77 | 56.33 | 13.72 | 19.00 | 10.95 |
| 33.31 | 8.22 | 1.25 | 6.32 | 1.21 | 7.25 | 3.38 | 0.53 | 0.50 | 3.14 | 8.41 | 59.80 | 9.47 | 18.69 | 12.04 |
| 20.91 | 9.89 | 1.78 | 5.61 | 1.99 | 8.30 | 4.07 | 0.90 | 0.44 | 2.43 | 8.34 | 56.24 | 11.68 | 20.88 | 11.21 |
| 28.00 | 9.57 | 1.81 | 5.23 | 1.13 | 11.23 | 4.75 | 1.19 | 0.31 | 4.68 | 4.62 | 55.48 | 11.38 | 23.53 | 9.61 |
| 26.19 | 8.85 | 1.63 | 6.24 | 1.16 | 11.52 | 4.74 | 0.78 | 0.31 | 3.80 | 5.41 | 55.56 | 10.48 | 24.44 | 9.52 |
| 21.25 | 12.65 | 2.62 | 6.66 | 0.98 | 9.10 | 3.82 | 0.97 | 0.56 | 3.08 | 8.21 | 51.37 | 15.27 | 21.52 | 11.84 |
| 24.72 | 11.08 | 1.92 | 6.11 | 2.13 | 8.05 | 4.30 | 0.73 | 0.51 | 3.14 | 7.49 | 54.55 | 13.00 | 21.32 | 11.13 |
| 22.95 | 11.53 | 1.99 | 6.20 | 1.29 | 10.22 | 3.69 | 0.86 | 0.69 | 4.58 | 8.13 | 50.82 | 13.51 | 22.26 | 13.40 |
| 27.92 | 8.97 | 1.46 | 4.61 | 0.95 | 9.84 | 4.34 | 0.71 | 0.45 | 3.23 | 7.67 | 57.78 | 10.43 | 20.45 | 11.34 |
| 17.73 | 11.38 | 2.19 | 6.55 | 1.40 | 9.74 | 4.04 | 0.52 | 0.24 | 1.87 | 5.62 | 56.46 | 13.56 | 22.25 | 7.73 |
| 20.12 | 9.89 | 1.91 | 7.08 | 1.45 | 9.01 | 4.05 | 0.46 | 0.35 | 1.70 | 5.96 | 58.14 | 11.80 | 22.05 | 8.01 |
| 21.88 | 10.50 | 2.25 | 7.18 | 1.13 | 10.55 | 4.68 | 0.64 | 0.44 | 2.24 | 7.06 | 53.32 | 12.75 | 24.18 | 9.75 |
| 20.48 | 12.26 | 1.85 | 5.83 | 1.19 | 11.13 | 4.89 | 0.61 | 0.40 | 3.30 | 4.85 | 53.69 | 14.11 | 23.64 | 8.55 |
| 26.69 | 11.46 | 1.94 | 6.87 | 1.58 | 9.54 | 5.02 | 1.16 | 0.28 | 3.93 | 4.40 | 53.81 | 13.41 | 24.17 | 8.61 |
| 21.06 | 10.32 | 1.80 | 7.29 | 1.57 | 9.85 | 3.43 | 0.52 | 0.48 | 2.77 | 5.68 | 56.28 | 12.12 | 22.66 | 8.94 |
| 24.15 | 11.57 | 2.16 | 6.99 | 1.46 | 7.83 | 3.06 | 0.84 | 0.32 | 1.91 | 6.49 | 57.37 | 13.73 | 20.18 | 8.72 |
| 26.52 | 9.69 | 2.02 | 5.11 | 1.30 | 9.32 | 4.01 | 1.04 | 0.27 | 2.58 | 5.02 | 59.64 | 11.71 | 20.77 | 7.88 |
| 23.58 | 10.58 | 1.99 | 5.74 | 0.99 | 8.11 | 2.62 | 0.50 | 0.52 | 2.67 | 5.72 | 60.55 | 12.58 | 17.96 | 8.91 |
| 24.62 | 9.25 | 1.76 | 5.82 | 1.05 | 6.79 | 2.18 | 0.12 | 0.39 | 2.16 | 6.32 | 64.15 | 11.02 | 15.95 | 8.88 |
| 25.05 | 10.55 | 1.86 | 6.68 | 1.06 | 11.68 | 5.01 | 0.93 | 0.34 | 3.62 | 5.24 | 53.04 | 12.41 | 25.34 | 9.20 |
| 21.94 | 9.95 | 2.18 | 6.50 | 1.04 | 11.87 | 4.19 | 0.83 | 0.59 | 3.39 | 8.81 | 50.68 | 12.12 | 24.42 | 12.78 |
| 22.69 | 10.02 | 1.65 | 7.99 | 1.18 | 8.77 | 3.08 | 0.69 | 0.40 | 2.14 | 5.41 | 58.66 | 11.67 | 21.72 | 7.95 |
| 30.05 | 7.95 | 1.50 | 5.07 | 1.36 | 8.85 | 4.68 | 1.28 | 0.23 | 3.21 | 5.64 | 60.23 | 9.45 | 21.23 | 9.08 |
| 20.29 | 10.29 | 1.59 | 6.14 | 1.16 | 9.65 | 3.08 | 0.80 | 0.51 | 2.17 | 5.27 | 59.34 | 11.88 | 20.83 | 7.95 |
| 28.74 | 10.05 | 1.38 | 5.65 | 0.97 | 8.12 | 3.80 | 0.71 | 0.38 | 3.01 | 5.86 | 60.07 | 11.43 | 19.25 | 9.25 |
| 25.55 | 10.82 | 2.08 | 5.45 | 1.10 | 10.96 | 5.78 | 0.92 | 0.38 | 3.17 | 5.24 | 54.11 | 12.90 | 24.21 | 8.78 |
| 28.21 | 12.94 | 2.11 | 6.46 | 0.80 | 8.05 | 3.50 | 0.78 | 0.23 | 2.24 | 5.40 | 57.48 | 15.05 | 19.60 | 7.87 |
| 16.38 | 13.87 | 2.70 | 6.14 | 0.86 | 11.86 | 3.79 | 0.70 | 0.65 | 3.10 | 4.66 | 51.67 | 16.56 | 23.36 | 8.41 |
| 18.14 | 14.04 | 2.67 | 6.27 | 1.38 | 10.11 | 4.36 | 0.80 | 0.46 | 3.04 | 8.01 | 48.87 | 16.71 | 22.92 | 11.51 |
| 21.68 | 11.11 | 1.86 | 7.18 | 1.19 | 10.60 | 4.06 | 1.08 | 0.35 | 3.35 | 7.05 | 52.17 | 12.97 | 24.11 | 10.74 |
| 36.01 | 9.70 | 1.10 | 5.06 | 2.31 | 6.23 | 3.46 | 0.12 | 0.45 | 1.83 | 4.04 | 65.70 | 10.80 | 17.18 | 6.33 |
| 27.61 | 10.45 | 2.35 | 5.63 | 1.65 | 10.91 | 4.87 | 0.89 | 0.59 | 2.75 | 6.22 | 53.69 | 12.81 | 23.95 | 9.56 |
| 26.04 | 11.41 | 1.75 | 5.78 | 1.19 | 9.94 | 3.97 | 0.82 | 0.78 | 3.61 | 6.95 | 53.79 | 13.16 | 21.71 | 11.34 |
| 25.04 | 10.06 | 2.04 | 5.79 | 1.72 | 8.91 | 4.67 | 0.90 | 0.58 | 3.48 | 6.74 | 55.11 | 12.10 | 21.99 | 10.81 |
| 23.97 | 10.05 | 1.75 | 5.89 | 1.99 | 9.97 | 6.95 | 0.62 | 0.63 | 2.66 | 7.78 | 51.70 | 11.80 | 25.42 | 11.07 |
| 25.08 | 11.44 | 2.05 | 5.50 | 1.47 | 7.91 | 3.14 | 0.24 | 0.49 | 2.74 | 6.18 | 58.85 | 13.49 | 18.26 | 9.41 |
