## Supplemental Table 2 for "An 8 week randomized Dietary Guidelines for Americans -based diet intervention improves the omega-3 index of healthy women"

**Supplemental Table S2: Participant estimated dietary fatty acid intake (g/d) based on calorie adjust diet percriptions**

| Diet Group | Subject_ID | Body Wt (kg) | Diet Calories | Diet Series | Day | C10:0 | C12:0 | C14:0 | C15:0 | C16:0 | C17:0 | C18:0 | C19:0 | C20:0 | C21:0 | C22:0 | C24:0 | C14:1n5 | C16:1n7 | C18:1n9 | C18:1n7 | C19:1n9 | C20:1n9 | C18:2n6 | C18:3n6 | 9c,11t-CLA | C20:3n6 | C20:4n6 | C22:4n6 | C22:5n6 | C18:3n3 | C18:4n3 | C20:3n3 | C20:4n3 | C20:5n3 | C22:5n3 | C22:6n3 | Σ SFA | Σ MUFA | Σ (n-6) PUFA | Σ (n-3) PUFA | Total Fatty Acids (m[illegible] | EPA + DHA (g/d) |
|---|---|---|---|---|---|---|---|---|---|---|---|---|---|---|---|---|---|---|---|---|---|---|---|---|---|---|---|---|---|---|---|---|---|---|---|---|---|---|---|---|---|---|---|
| DGA | 9 | 77.85 | 1900 | R | D1 | 0.43 | 0.27 | 2.03 | 0.16 | 17.46 | 0.18 | 8.29 | 0.02 | 0.16 | 0.00 | 0.14 | 0.07 | 0.12 | 1.02 | 41.47 | 2.11 | 0.01 | 0.25 | 22.05 | 0.06 | 0.03 | 0.04 | 0.10 | 0.02 | 0.01 | 3.35 | 0.13 | 0.02 | 0.03 | 0.27 | 0.29 | 2.26 | 29.22 | 44.99 | 22.31 | 4.86 | 101.37 | 2.53 |
| DGA | 32 | 114.4 | 3000 | R | D1 | 0.64 | 0.39 | 2.98 | 0.24 | 25.70 | 0.26 | 12.20 | 0.04 | 0.24 | 0.01 | 0.21 | 0.11 | 0.17 | 1.50 | 61.04 | 3.11 | 0.01 | 0.37 | 32.46 | 0.08 | 0.04 | 0.05 | 0.15 | 0.04 | 0.02 | 4.93 | 0.19 | 0.03 | 0.04 | 0.40 | 0.43 | 3.32 | 43.01 | 66.22 | 32.85 | 7.15 | 149.23 | 3.72 |
| DGA | 72 | 84.75 | 2300 | R | D1 | 0.53 | 0.33 | 2.47 | 0.20 | 21.33 | 0.21 | 10.13 | 0.03 | 0.20 | 0.01 | 0.18 | 0.09 | 0.14 | 1.25 | 50.67 | 2.58 | 0.01 | 0.31 | 26.95 | 0.07 | 0.03 | 0.04 | 0.13 | 0.03 | 0.01 | 4.09 | 0.16 | 0.02 | 0.03 | 0.33 | 0.36 | 2.76 | 35.70 | 54.97 | 27.27 | 5.94 | 123.88 | 3.09 |
| DGA | 79 | 94.1 | 2300 | R | D1 | 0.53 | 0.33 | 2.47 | 0.20 | 21.33 | 0.21 | 10.13 | 0.03 | 0.20 | 0.01 | 0.18 | 0.09 | 0.14 | 1.25 | 50.67 | 2.58 | 0.01 | 0.31 | 26.95 | 0.07 | 0.03 | 0.04 | 0.13 | 0.03 | 0.01 | 4.09 | 0.16 | 0.02 | 0.03 | 0.33 | 0.36 | 2.76 | 35.70 | 54.97 | 27.27 | 5.94 | 123.88 | 3.09 |
| DGA | 84 | 98.8 | 2700 | R | D1 | 0.66 | 0.41 | 3.07 | 0.25 | 26.50 | 0.27 | 12.58 | 0.04 | 0.24 | 0.01 | 0.22 | 0.11 | 0.18 | 1.55 | 62.96 | 3.21 | 0.02 | 0.38 | 33.48 | 0.09 | 0.04 | 0.05 | 0.16 | 0.04 | 0.02 | 5.08 | 0.20 | 0.03 | 0.04 | 0.41 | 0.44 | 3.43 | 44.36 | 68.30 | 33.88 | 7.38 | 153.91 | 3.84 |
| DGA | 96 | 70.2 | 1800 | R | D1 | 0.42 | 0.25 | 1.93 | 0.16 | 16.67 | 0.17 | 7.91 | 0.02 | 0.15 | 0.00 | 0.14 | 0.07 | 0.11 | 0.97 | 39.59 | 2.02 | 0.01 | 0.24 | 21.06 | 0.05 | 0.03 | 0.03 | 0.10 | 0.02 | 0.01 | 3.20 | 0.12 | 0.02 | 0.03 | 0.26 | 0.28 | 2.15 | 27.90 | 42.96 | 21.30 | 4.64 | 96.80 | 2.41 |
| DGA | 97 | 59.65 | 1700 | R | D1 | 0.42 | 0.26 | 1.94 | 0.16 | 16.70 | 0.17 | 7.93 | 0.02 | 0.15 | 0.00 | 0.14 | 0.07 | 0.11 | 0.98 | 39.66 | 2.02 | 0.01 | 0.24 | 21.09 | 0.05 | 0.03 | 0.03 | 0.10 | 0.02 | 0.01 | 3.20 | 0.12 | 0.02 | 0.03 | 0.26 | 0.28 | 2.16 | 27.94 | 43.03 | 21.34 | 4.65 | 96.96 | 2.42 |
| DGA | 102 | 69.95 | 1800 | R | D1 | 0.42 | 0.25 | 1.93 | 0.16 | 16.67 | 0.17 | 7.91 | 0.02 | 0.15 | 0.00 | 0.14 | 0.07 | 0.11 | 0.97 | 39.59 | 2.02 | 0.01 | 0.24 | 21.06 | 0.05 | 0.03 | 0.03 | 0.10 | 0.02 | 0.01 | 3.20 | 0.12 | 0.02 | 0.03 | 0.26 | 0.28 | 2.15 | 27.90 | 42.96 | 21.30 | 4.64 | 96.80 | 2.41 |
| DGA | 107 | 92.3 | 2200 | R | D1 | 0.54 | 0.33 | 2.51 | 0.20 | 21.60 | 0.22 | 10.26 | 0.03 | 0.20 | 0.01 | 0.18 | 0.09 | 0.15 | 1.26 | 51.31 | 2.61 | 0.01 | 0.31 | 27.29 | 0.07 | 0.04 | 0.04 | 0.13 | 0.03 | 0.01 | 4.14 | 0.16 | 0.02 | 0.03 | 0.34 | 0.36 | 2.79 | 36.15 | 55.66 | 27.61 | 6.01 | 125.44 | 3.13 |
| DGA | 113 | 105.9 | 2600 | R | D1 | 0.60 | 0.37 | 2.78 | 0.23 | 24.00 | 0.24 | 11.40 | 0.03 | 0.22 | 0.01 | 0.20 | 0.10 | 0.16 | 1.40 | 57.02 | 2.91 | 0.01 | 0.35 | 30.33 | 0.08 | 0.04 | 0.05 | 0.14 | 0.03 | 0.01 | 4.60 | 0.18 | 0.03 | 0.04 | 0.37 | 0.40 | 3.10 | 40.17 | 61.86 | 30.68 | 6.68 | 139.40 | 3.48 |
| DGA | 139 | 72.55 | 1800 | R | D1 | 0.42 | 0.25 | 1.93 | 0.16 | 16.67 | 0.17 | 7.91 | 0.02 | 0.15 | 0.00 | 0.14 | 0.07 | 0.11 | 0.97 | 39.59 | 2.02 | 0.01 | 0.24 | 21.06 | 0.05 | 0.03 | 0.03 | 0.10 | 0.02 | 0.01 | 3.20 | 0.12 | 0.02 | 0.03 | 0.26 | 0.28 | 2.15 | 27.90 | 42.96 | 21.30 | 4.64 | 96.80 | 2.41 |
| DGA | 154 | 93.8 | 2200 | R | D1 | 0.54 | 0.33 | 2.51 | 0.20 | 21.60 | 0.22 | 10.26 | 0.03 | 0.20 | 0.01 | 0.18 | 0.09 | 0.15 | 1.26 | 51.31 | 2.61 | 0.01 | 0.31 | 27.29 | 0.07 | 0.04 | 0.04 | 0.13 | 0.03 | 0.01 | 4.14 | 0.16 | 0.02 | 0.03 | 0.34 | 0.36 | 2.79 | 36.15 | 55.66 | 27.61 | 6.01 | 125.44 | 3.13 |
| DGA | 172 | 92.65 | 2400 | R | D1 | 0.55 | 0.34 | 2.57 | 0.21 | 22.17 | 0.22 | 10.53 | 0.03 | 0.20 | 0.01 | 0.18 | 0.09 | 0.15 | 1.30 | 52.67 | 2.68 | 0.01 | 0.32 | 28.01 | 0.07 | 0.04 | 0.05 | 0.13 | 0.03 | 0.01 | 4.25 | 0.16 | 0.02 | 0.04 | 0.34 | 0.37 | 2.87 | 37.11 | 57.14 | 28.34 | 6.17 | 128.75 | 3.21 |
| DGA | 182 | 108.5 | 2400 | R | D1 | 0.55 | 0.34 | 2.57 | 0.21 | 22.17 | 0.22 | 10.53 | 0.03 | 0.20 | 0.01 | 0.18 | 0.09 | 0.15 | 1.30 | 52.67 | 2.68 | 0.01 | 0.32 | 28.01 | 0.07 | 0.04 | 0.05 | 0.13 | 0.03 | 0.01 | 4.25 | 0.16 | 0.02 | 0.04 | 0.34 | 0.37 | 2.87 | 37.11 | 57.14 | 28.34 | 6.17 | 128.75 | 3.21 |
| DGA | 189 | 77.45 | 2000 | R | D1 | 0.45 | 0.28 | 2.12 | 0.17 | 18.24 | 0.18 | 8.66 | 0.03 | 0.17 | 0.00 | 0.15 | 0.07 | 0.12 | 1.07 | 43.34 | 2.21 | 0.01 | 0.26 | 23.05 | 0.06 | 0.03 | 0.04 | 0.11 | 0.03 | 0.01 | 3.50 | 0.14 | 0.02 | 0.03 | 0.28 | 0.30 | 2.36 | 30.53 | 47.02 | 23.32 | 5.08 | 105.95 | 2.64 |
| DGA | 194 | 69.75 | 1800 | R | D1 | 0.42 | 0.25 | 1.93 | 0.16 | 16.67 | 0.17 | 7.91 | 0.02 | 0.15 | 0.00 | 0.14 | 0.07 | 0.11 | 0.97 | 39.59 | 2.02 | 0.01 | 0.24 | 21.06 | 0.05 | 0.03 | 0.03 | 0.10 | 0.02 | 0.01 | 3.20 | 0.12 | 0.02 | 0.03 | 0.26 | 0.28 | 2.15 | 27.90 | 42.96 | 21.30 | 4.64 | 96.80 | 2.41 |
| DGA | 198 | 104.1 | 2350 | R | D1 | 0.54 | 0.33 | 2.52 | 0.20 | 21.75 | 0.22 | 10.33 | 0.03 | 0.20 | 0.01 | 0.18 | 0.09 | 0.15 | 1.27 | 51.67 | 2.63 | 0.01 | 0.31 | 27.48 | 0.07 | 0.04 | 0.04 | 0.13 | 0.03 | 0.01 | 4.17 | 0.16 | 0.02 | 0.03 | 0.34 | 0.36 | 2.81 | 36.40 | 56.05 | 27.80 | 6.05 | 126.31 | 3.15 |
| DGA | 234 | 98.7 | 2300 | R | D1 | 0.53 | 0.33 | 2.47 | 0.20 | 21.33 | 0.21 | 10.13 | 0.03 | 0.20 | 0.01 | 0.18 | 0.09 | 0.14 | 1.25 | 50.67 | 2.58 | 0.01 | 0.31 | 26.95 | 0.07 | 0.03 | 0.04 | 0.13 | 0.03 | 0.01 | 4.09 | 0.16 | 0.02 | 0.03 | 0.33 | 0.36 | 2.76 | 35.70 | 54.97 | 27.27 | 5.94 | 123.88 | 3.09 |
| DGA | 243 | 75.75 | 2300 | R | D1 | 0.53 | 0.33 | 2.47 | 0.20 | 21.33 | 0.21 | 10.13 | 0.03 | 0.20 | 0.01 | 0.18 | 0.09 | 0.14 | 1.25 | 50.67 | 2.58 | 0.01 | 0.31 | 26.95 | 0.07 | 0.03 | 0.04 | 0.13 | 0.03 | 0.01 | 4.09 | 0.16 | 0.02 | 0.03 | 0.33 | 0.36 | 2.76 | 35.70 | 54.97 | 27.27 | 5.94 | 123.88 | 3.09 |
| DGA | 244 | 101.25 | 2500 | R | D1 | 0.57 | 0.35 | 2.68 | 0.22 | 23.06 | 0.23 | 10.95 | 0.03 | 0.21 | 0.01 | 0.19 | 0.09 | 0.15 | 1.35 | 54.78 | 2.79 | 0.01 | 0.33 | 29.13 | 0.08 | 0.04 | 0.05 | 0.14 | 0.03 | 0.01 | 4.42 | 0.17 | 0.02 | 0.04 | 0.36 | 0.38 | 2.98 | 38.60 | 59.43 | 29.48 | 6.42 | 133.93 | 3.34 |
| DGA | 245 | 108.8 | 2300 | R | D1 | 0.53 | 0.33 | 2.47 | 0.20 | 21.33 | 0.21 | 10.13 | 0.03 | 0.20 | 0.01 | 0.18 | 0.09 | 0.14 | 1.25 | 50.67 | 2.58 | 0.01 | 0.31 | 26.95 | 0.07 | 0.03 | 0.04 | 0.13 | 0.03 | 0.01 | 4.09 | 0.16 | 0.02 | 0.03 | 0.33 | 0.36 | 2.76 | 35.70 | 54.97 | 27.27 | 5.94 | 123.88 | 3.09 |
| DGA | 258 | 78.1 | 2200 | R | D1 | 0.54 | 0.33 | 2.51 | 0.20 | 21.60 | 0.22 | 10.26 | 0.03 | 0.20 | 0.01 | 0.18 | 0.09 | 0.15 | 1.26 | 51.31 | 2.61 | 0.01 | 0.31 | 27.29 | 0.07 | 0.04 | 0.04 | 0.13 | 0.03 | 0.01 | 4.14 | 0.16 | 0.02 | 0.03 | 0.34 | 0.36 | 2.79 | 36.15 | 55.66 | 27.61 | 6.01 | 125.44 | 3.13 |
| TAD | 3 | 109.35 | 2200 | S | D1 | 2.75 | 2.71 | 10.41 | 0.90 | 51.05 | 0.77 | 20.19 | 0.07 | 0.16 | 0.01 | 0.10 | 0.07 | 0.95 | 2.87 | 62.56 | 3.42 | 0.03 | 0.12 | 30.02 | 0.03 | 0.22 | 0.05 | 0.18 | 0.03 | 0.01 | 3.59 | 0.02 | 0.00 | 0.01 | 0.02 | 0.12 | 0.15 | 89.19 | 69.95 | 30.55 | 3.81 | 193.50 | 0.17 |
| TAD | 20 | 81.4 | 2000 | S | D1 | 2.46 | 2.42 | 9.30 | 0.81 | 45.62 | 0.69 | 18.04 | 0.06 | 0.15 | 0.01 | 0.09 | 0.06 | 0.85 | 2.57 | 55.90 | 3.06 | 0.02 | 0.11 | 26.83 | 0.03 | 0.19 | 0.05 | 0.16 | 0.03 | 0.01 | 3.21 | 0.01 | 0.00 | 0.00 | 0.02 | 0.11 | 0.14 | 79.70 | 62.51 | 27.30 | 3.40 | 172.91 | 0.15 |
| TAD | 51 | 95.8 | 2300 | S | D1 | 2.86 | 2.82 | 10.83 | 0.94 | 53.09 | 0.80 | 20.99 | 0.07 | 0.17 | 0.01 | 0.11 | 0.07 | 0.99 | 2.99 | 65.06 | 3.56 | 0.03 | 0.13 | 31.22 | 0.04 | 0.22 | 0.06 | 0.19 | 0.03 | 0.01 | 3.73 | 0.02 | 0.00 | 0.01 | 0.02 | 0.13 | 0.16 | 92.75 | 72.74 | 31.77 | 3.96 | 201.22 | 0.18 |
| TAD | 52 | 82.9 | 1900 | S | D1 | 2.35 | 2.31 | 8.88 | 0.77 | 43.56 | 0.66 | 17.23 | 0.06 | 0.14 | 0.01 | 0.09 | 0.06 | 0.81 | 2.45 | 53.38 | 2.92 | 0.02 | 0.10 | 25.62 | 0.03 | 0.18 | 0.05 | 0.15 | 0.03 | 0.01 | 3.06 | 0.01 | 0.00 | 0.00 | 0.02 | 0.10 | 0.13 | 76.11 | 59.69 | 26.07 | 3.25 | 165.11 | 0.15 |
| TAD | 53 | 88.15 | 2300 | S | D1 | 2.86 | 2.82 | 10.83 | 0.94 | 53.09 | 0.80 | 20.99 | 0.07 | 0.17 | 0.01 | 0.11 | 0.07 | 0.99 | 2.99 | 65.06 | 3.56 | 0.03 | 0.13 | 31.22 | 0.04 | 0.22 | 0.06 | 0.19 | 0.03 | 0.01 | 3.73 | 0.02 | 0.00 | 0.01 | 0.02 | 0.13 | 0.16 | 92.75 | 72.74 | 31.77 | 3.96 | 201.22 | 0.18 |
| TAD | 69 | 77.3 | 2200 | S | D1 | 2.75 | 2.71 | 10.41 | 0.90 | 51.05 | 0.77 | 20.19 | 0.07 | 0.16 | 0.01 | 0.10 | 0.07 | 0.95 | 2.87 | 62.56 | 3.42 | 0.03 | 0.12 | 30.02 | 0.03 | 0.22 | 0.05 | 0.18 | 0.03 | 0.01 | 3.59 | 0.02 | 0.00 | 0.01 | 0.02 | 0.12 | 0.15 | 89.19 | 69.95 | 30.55 | 3.81 | 193.50 | 0.17 |
| [illegible] | [illegible] | [illegible] | [illegible] | [illegible] | [illegible] | [illegible] | [illegible] | [illegible] | [illegible] | [illegible] | [illegible] | [illegible] | [illegible] | [illegible] | [illegible] | [illegible] | [illegible] | [illegible] | [illegible] | [illegible] | [illegible] | [illegible] | [illegible] | [illegible] | [illegible] | [illegible] | [illegible] | [illegible] | [illegible] | [illegible] | [illegible] | [illegible] | [illegible] | [illegible] | [illegible] | [illegible] | [illegible] | [illegible] | [illegible] | [illegible] | [illegible] | [illegible] | [illegible] |
