## Supplemental Table 3 for "An 8 week randomized Dietary Guidelines for Americans -based diet intervention improves the omega-3 index of healthy women"

**Supplemental Table S3: Daily gram weights of foods providing >0.1g of omega-3 fatty acids and NDSR-estimated fatty acid amounts in the TAD and DGAD menu plans[a]**

| TAD | | | | DAGD | | | |
|---|---|---|---|---|---|---|---|
| FoodDescription | Food (g/d) | EPA+DHA (g/d) | ALA (g/d) | FoodDescription | Food (g/d) | EPA+DHA (g/d) | ALA (g/d) |
| Estimated 8-day rotation Content | | 0.5 | 13.7 | | | 4.2 | 14.9 |
| | | | | Day 1 | | | |
| shrimp, cooked from frozen | 30 | 0.1 | | BAKED SALMON | 80 | 1.7 | 0.1 |
| dressing for salads, Hidden Valley Original Ranch - bottled | 30 | | 1.2 | RANCH Dressing | 30 | | 1.4 |
| spaghetti noodles, white, cooked in unsalted water | 100 | | 0.3 | ROASTED POTATOES | 225 | | 0.3 |
| corn chips, ingredient fat known | 10 | | 0.1 | cereal, ready-to-eat, Cinnamon Chex (General Mills) | 30 | | 0.2 |
| Cheddar cheese, natural | 30 | | 0.1 | taco, tortilla or nacho chips, regular, salted, ingredient fat known | 10 | | 0.1 |
| ROASTED POTATOES | 75 | | 0.1 | EGG PATTY IQF | 43 | | 0.1 |
| EGG PATTY IQ | 43 | | 0.1 | SOUTHWEST SOUP | 180 | | 0.1 |
| SOUTHWEST SOUP | 180 | | 0.1 | beans, black, canned - drained, regular | 30 | | 0.1 |
| cream, half and half, regular (10-12% fat) | 30 | | 0.1 | | | | |
| | | | | Day 2 | | | |
| PIZZA | 226 | | 0.4 | STUFFING | 150 | | 1.4 |
| pizza, crust, white, thin | 86 | | 0.3 | MASHED POTATOES | 150 | | 0.4 |
| MASHED POTATOES | 75 | | 0.2 | pizza, crust, white, thin | 86 | | 0.3 |
| stuffing, white bread, made from mix | 75 | | 0.1 | cereal, ready-to-eat, Cinnamon Chex (General Mills) | 30 | | 0.2 |
| CRISPY RICE TREAT | 45 | | 0.1 | turkey, bacon, regular | 20 | | 0.1 |
| turkey, bacon, regular | 20 | | 0.1 | lettuce, romaine or cos | 50 | | 0.1 |
| PUMPKIN CAKE ICING | 15 | | 0.1 | ROASTED MUSHROOMS | 25 | | 0.1 |
| sauce, marinara, commercial | 50 | | 0.1 | | | | |
| ROASTED MUSHROOMS | 25 | | 0.1 | | | | |
| cream, half and half, regular (10-12% fat) | 30 | | 0.1 | | | | |
| | | | | Day 3 | | | |
| BAGEL | 88 | | 0.1 | GODDESS DRESSING | 30 | | 0.8 |
| Cheddar cheese, natural | 30 | | 0.1 | ROASTED POTATOES | 150 | | 0.2 |
| ROASTED POTATOES | 75 | | 0.1 | lettuce, romaine or cos | 50 | | 0.1 |
| turkey, bacon, regular | 20 | | 0.1 | | | | |
| BBQ BRISKET | 100 | | 0.1 | | | | |
| cream, half and half, regular (10-12% fat) | 30 | | 0.1 | | | | |
| | | | | Day 4 | | | |
| SALSA RANCH DRESSING | 30 | | 1.2 | SALSA RANCH | 30 | | 1.3 |
| PASTA WITH MEAT SAUCE | 340 | | 0.5 | taco, tortilla or nacho chips, regular, salted, ingredient fat known | 30 | | 0.2 |
| spaghetti noodles, white, cooked in unsalted water | 100 | | 0.3 | TOMATO-BASIL SAUCE | 150 | | 0.1 |
| sauce, marinara, commercial | 100 | | 0.1 | ROASTED MUSHROOMS | 50 | | 0.1 |
| Cheddar cheese, natural | 15 | | 0.1 | MASHED PINTOS | 80 | | 0.1 |
| ground beef or hamburger, 10% fat (90% lean meat) | 120 | | 0.1 | green or string beans, cooked from frozen | 120 | | 0.1 |
| cream, half and half, regular (10-12% fat) | 30 | | 0.1 | oil, olive | 7 | | 0.1 |
| | | | | Day 5 | | | |
| shrimp, cooked from frozen | 30 | 0.1 | | BAKED SALMON | 80 | 1.7 | 0.1 |
| BREAKFAST MUFFINS | 65 | | 0.2 | CORN CHOWDER | 180 | 0.1 | 0 |
| ROASTED POTATOES | 75 | | 0.1 | SALSA RANCH | 30 | | 1.3 |
| Colby Jack cheese, regular | 21.3 | | 0.1 | walnuts | 5 | | 0.5 |
| CORN CHOWDER | 180 | | 0.1 | CINNAMON APPLES | 150 | | 0.1 |
| cream, half and half, regular (10-12% fat) | 30 | | 0.1 | cereal, ready-to-eat, Cinnamon Chex (General Mills) | 15 | | 0.1 |
| | | | | BREAKFAST MUFFIN | 65 | | 0.1 |
| | | | | lettuce, romaine or cos | 50 | | 0.1 |
| | | | | Day 6 | | | |
| dressing for salads, Girard's Caesar | 30 | | 1 | RANCH | 30 | | 1.4 |
| CORN-CHEDDAR BISCUIT | 130 | | 0.3 | walnuts | 5 | | 0.5 |
| granola bars, Kellogg's Nutri-Grain Cereal Bar - all flavors | 37 | | 0.2 | ROASTED POTATOES | 150 | | 0.2 |
| ROASTED POTATOES | 75 | | 0.1 | CORN-CHEDDAR BISCUIT | 65 | | 0.1 |
| CHILI BASE | 80 | | 0.1 | lettuce, romaine or cos | 50 | | 0.1 |
| Cheddar cheese, natural | 15 | | 0.1 | Cheddar cheese, natural | 15 | | 0.1 |
| butter, regular, unsalted | 16 | | 0.1 | MASHED PINTOS | 40 | | 0.1 |
| cream, half and half, regular (10-12% fat) | 30 | | 0.1 | | | | |
| | | | | Day 7 | | | |
| SEAFOOD SALAD | 100 | 0.3 | 0.5 | canned, white albacore tuna, water pack, regular, drained - not rinsed | 80 | 0.7 | 0.1 |
| crackers, Keebler Club - Original | 28 | | 0.4 | dressing for salads, thousand island, regular | 30 | | 0.7 |
| POPCORN CHICKEN | 35 | | 0.4 | walnuts | 5 | | 0.5 |
| corn chips, ingredient fat known | 30 | | 0.4 | CINNAMON APPLES | 150 | | 0.1 |
| cookies and bars, Nabisco Oreo Golden Sandwich - Original | 20 | | 0.2 | lettuce, romaine or cos | 50 | | 0.1 |
| MASHED PINTOS | 60 | | 0.2 | | | | |
| CHEESE SAUCE | 60 | | 0.1 | | | | |
| pecans, raw (dried) | 10 | | 0.1 | | | | |
| sauce, sweet and sour, commercial | 90 | | 0.1 | | | | |
| cream, half and half, regular (10-12% fat) | 30 | | 0.1 | | | | |
| | | | | Day 8 | | | |
| commercial pre-coated or breaded chicken, patty | 85 | | 1.3 | MASHED PINTOS | 80 | | 0.1 |
| PANCAKES | 150 | | 0.1 | BLUEBERRY SAUCE | 100 | | 0.1 |
| roast beef, chuck, no visible fat eaten | 100 | | 0.1 | turkey, bacon, regular | 20 | | 0.1 |
| Cheddar cheese, natural | 30 | | 0.1 | roast beef, chuck, no visible fat eaten | 50 | | 0.1 |
| turkey, bacon, regular | 20 | | 0.1 | SEASONED RICE | 150 | | 0.1 |
| CHEESE SAUCE | 30 | | 0.1 | | | | |
| cream, half and half, regular (10-12% fat) | 30 | | 0.1 | | | | |

*a* - NDSR - Nutrition Data System for Research (v 2014), University of Minnesota - Nutrition Coordinating Center (http://www.ncc.umn.edu/products/). Differences in estimated fatty acid levels for similar items are related to the manner of preparation.
