## Supplemental Table 4 for "An 8 week randomized Dietary Guidelines for Americans -based diet intervention improves the omega-3 index of healthy women"

**Supplemental Table 4: Average Block food frequency questionaire summary outputs**

| Subject_ID | Exclusison Comments | Diet Group | Study Week | Diet Week | Baseline Adj OM3 Change Cluster | DT_KCAL | DT_PROT | DT_CARB | DT_TFAT | DT_ALCO |
|---|---|---|---|---|---|---|---|---|---|---|
| 3 | | TAD | 1 | 0 | TAD | 5231 | 222.47 | 725.98 | 175.37 | 0.942 |
| 9 | | DGAD | 1 | 0 | DGAD Low Resp | 3266.58 | 112.38 | 387.02 | 144.24 | 1.09 |
| 20 | | TAD | 1 | 0 | TAD | 2929.32 | 103.54 | 392.14 | 110.22 | 0.936 |
| 32 | | DGAD | 1 | 0 | DGAD Low Resp | 2511.62 | 98.34 | 303.62 | 107.41 | 1.25 |
| 51 | Missing terminal blood draw | TAD | 1 | 0 | TAD | 1999.45 | 84.77 | 234.01 | 80.4 | 3.41 |
| 52 | | TAD | 1 | 0 | TAD | 1787.9 | 74.01 | 161.04 | 93.9 | 4.32 |
| 53 | | TAD | 1 | 0 | TAD | 4044.64 | 133.65 | 492.64 | 178.7 | 0.538 |
| 69 | | TAD | 1 | 0 | TAD | 1465.08 | 34.82 | 102.27 | 36.34 | 87.45 |
| 72 | | DGAD | 1 | 0 | DGAD Hi Resp | 1354.59 | 52.53 | 122.93 | 73.01 | 3.88 |
| 79 | | DGAD | 1 | 0 | DGAD Hi Resp | 1882.21 | 74.55 | 203.93 | 87.5 | 0.00115 |
| 84 | | DGAD | 1 | 0 | DGAD Hi Resp | 2634.05 | 97.05 | 345.04 | 101.13 | 2.62 |
| 85 | | TAD | 1 | 0 | TAD | 2269.79 | 87.91 | 272.95 | 80.76 | 20.07 |
| 87 | | TAD | 1 | 0 | TAD | 1565.91 | 78.55 | 184.07 | 59.78 | 0.745 |
| 96 | | DGAD | 1 | 0 | DGAD Hi Resp | 1284.41 | 37.31 | 175.74 | 52.05 | 1.17 |
| 97 | | DGAD | 1 | 0 | DGAD Hi Resp | 2168.15 | 83.67 | 268.72 | 85.77 | 2.88 |
| 102 | | DGAD | 1 | 0 | DGAD Hi Resp | 1297.24 | 48.47 | 127.8 | 54.64 | 18.47 |
| 107 | | DGAD | 1 | 0 | DGAD Hi Resp | 3315.06 | 123.5 | 403.02 | 141.83 | 0 |
| 109 | | TAD | 1 | 0 | TAD | 1705.79 | 73.98 | 177.23 | 66.56 | 16.97 |
| 112 | | TAD | 1 | 0 | TAD | 1838.88 | 69.79 | 201.4 | 84.7 | 1.76 |
| 113 | | DGAD | 1 | 0 | DGAD Hi Resp | 7378.84 | 329.19 | 822.34 | 307.83 | 8.08 |
| 135 | | TAD | 1 | 0 | TAD | 3635.83 | 139.03 | 467.64 | 140.26 | 0.306 |
| 137 | | TAD | 1 | 0 | TAD | 1998.46 | 93.67 | 183.63 | 101.94 | 0 |
| 139 | | DGAD | 1 | 0 | DGAD Hi Resp | 1308.07 | 52.11 | 159.71 | 51.9 | 4 |
| 144 | | TAD | 1 | 0 | TAD | 1568.83 | 51.23 | 133.62 | 96 | 1.99 |
| 151 | Missing baseline blood draw | TAD | 1 | 0 | TAD | 2573.4 | 96.31 | 313.57 | 112.57 | 0.01 |
| 154 | | DGAD | 1 | 0 | DGAD Low Resp | 2225.16 | 77.8 | 240.49 | 111.74 | 2.26 |
| 168 | | TAD | 1 | 0 | TAD | 2297.4 | 96.26 | 216.08 | 111.64 | 12.61 |
| 170 | | TAD | 1 | 0 | TAD | 2935.12 | 108.15 | 332.68 | 136.12 | 0.139 |
| 172 | | DGAD | 1 | 0 | DGAD Hi Resp | 1176.16 | 52.17 | 114.81 | 51.56 | 8.34 |
| 175 | | TAD | 1 | 0 | TAD | 3358.5 | 145.02 | 395.32 | 139.72 | 0.889 |
| 180 | | TAD | 1 | 0 | TAD | 3308.69 | 127.26 | 369.47 | 158.6 | 0.00231 |
| 182 | | DGAD | 1 | 0 | DGAD Low Resp | 2371.8 | 109.07 | 300.84 | 89.62 | 0.289 |
| 183 | | TAD | 1 | 0 | TAD | 2942.6 | 105.27 | 367.56 | 120.97 | 0.647 |
| 186 | | TAD | 1 | 0 | TAD | 2109.47 | 96.63 | 151.89 | 103.6 | 28.08 |
| 189 | | DGAD | 1 | 0 | DGAD Low Resp | 2621.52 | 117.6 | 289.46 | 116.71 | 1.01 |
| 194 | | DGAD | 1 | 0 | DGAD Low Resp | 1635.95 | 77.2 | 152.29 | 78.29 | 6.69 |
| 198 | screening and Visit 1 TG mismatch | DGAD | 1 | 0 | DGAD Hi Resp | 2599.99 | 87.84 | 283.17 | 127.67 | 0 |
| 232 | | TAD | 1 | 0 | TAD | 547.46 | 22.37 | 62.93 | 23.59 | 1.31 |
| 234 | | DGAD | 1 | 0 | DGAD Low Resp | 1581.16 | 72.1 | 162.12 | 72.37 | 0.937 |
| 239 | | TAD | 1 | 0 | TAD | 1801.79 | 59.06 | 170.31 | 95.85 | 7.39 |
| 243 | | DGAD | 1 | 0 | DGAD Low Resp | 810.08 | 36.84 | 100.66 | 27.84 | 3.23 |
| 244 | | DGAD | 1 | 0 | DGAD Hi Resp | 1117.58 | 53.91 | 130.24 | 43.7 | 0 |
| 245 | | DGAD | 1 | 0 | DGAD Hi Resp | 2943.95 | 132.28 | 311.8 | 132.53 | 1.17 |
| 258 | | DGAD | 1 | 0 | DGAD Low Resp | 1504.57 | 61.54 | 161.29 | 68.9 | 3.38 |

| DT_SUG_T | DT_FIBE | DT_MOIS | DT_SFAT | DT_MFAT | DT_PFAT | DT_CHOL | CARB/FAT_DT | MFAT/SFAT | PFAT/SFAT | SFAT/TFAT | MFAT/TFAT | PFAT/TFAT |
|---|---|---|---|---|---|---|---|---|---|---|---|---|
| 302.54 | 91.79 | 4984.76 | 50.93 | 64.39 | 44.35 | 463.32 | 4.139705 | 1.264284 | 0.870803 | 0.290415 | 0.367167 | 0.252894 |
| 155.32 | 27.49 | 2193.2 | 52.5 | 51.23 | 29.52 | 322.11 | 2.683167 | 0.97581 | 0.562286 | 0.363977 | 0.355172 | 0.204659 |
| 151.92 | 45.92 | 3010.69 | 35.75 | 39.08 | 26.8 | 201.51 | 3.557794 | 1.093147 | 0.74965 | 0.324351 | 0.354564 | 0.24315 |
| 112.79 | 42.83 | 3148.39 | 34.16 | 40.95 | 23.44 | 334.34 | 2.826739 | 1.19877 | 0.686183 | 0.318034 | 0.381249 | 0.218229 |
| 71.58 | 21.58 | 1308.29 | 25.91 | 30.3 | 17.04 | 266.8 | 2.910572 | 1.169433 | 0.657661 | 0.322264 | 0.376866 | 0.21194 |
| 48.75 | 17.16 | 1930.98 | 34.3 | 35.94 | 16.53 | 311.09 | 1.715016 | 1.047813 | 0.481924 | 0.365282 | 0.382748 | 0.176038 |
| 195.94 | 44.18 | 3023.67 | 54.2 | 71.88 | 39.64 | 386.39 | 2.756799 | 1.326199 | 0.731365 | 0.303302 | 0.402238 | 0.221824 |
| 54.94 | 5.54 | 2741.7 | 12.82 | 12.92 | 7.26 | 142.48 | 2.814254 | 1.0078 | 0.566303 | 0.352779 | 0.355531 | 0.19978 |
| 43.2 | 16.45 | 2051.72 | 23.41 | 27.62 | 16.79 | 190.26 | 1.683742 | 1.179838 | 0.717215 | 0.320641 | 0.378304 | 0.229968 |
| 55.51 | 18.75 | 817.63 | 30.6 | 31.95 | 17.51 | 265.76 | 2.330629 | 1.044118 | 0.572222 | 0.349714 | 0.365143 | 0.200114 |
| 125.79 | 35.84 | 3440.96 | 31.32 | 37.39 | 23.78 | 240.56 | 3.411846 | 1.193806 | 0.759259 | 0.3097 | 0.369722 | 0.235143 |
| 105.18 | 26.15 | 2959.12 | 25.66 | 29.48 | 18.64 | 262.08 | 3.379767 | 1.14887 | 0.726422 | 0.317732 | 0.365032 | 0.230807 |
| 69.51 | 17.93 | 1611.9 | 17.03 | 23.42 | 14.21 | 178.89 | 3.079123 | 1.37522 | 0.83441 | 0.284878 | 0.39177 | 0.237705 |
| 87.41 | 15.86 | 3728.03 | 15.64 | 18.16 | 14.55 | 104.48 | 3.376369 | 1.161125 | 0.930307 | 0.30048 | 0.348895 | 0.279539 |
| 118.3 | 22.87 | 2494.62 | 26.9 | 31.64 | 20.19 | 258.09 | 3.13303 | 1.176208 | 0.750558 | 0.313629 | 0.368894 | 0.235397 |
| 46.36 | 15.87 | 2010.42 | 13.98 | 21.46 | 14.97 | 168.77 | 2.338946 | 1.53505 | 1.070815 | 0.255857 | 0.392753 | 0.273975 |
| 182.93 | 37.37 | 3334.34 | 46.31 | 51.67 | 32.32 | 597.29 | 2.841571 | 1.115742 | 0.697905 | 0.326518 | 0.364309 | 0.227878 |
| 65.86 | 14.69 | 2492.13 | 21.02 | 25.13 | 14.22 | 429.77 | 2.66271 | 1.195528 | 0.676499 | 0.315805 | 0.377554 | 0.213642 |
| 66.42 | 16.16 | 1811.11 | 25.94 | 32.42 | 19.22 | 253.56 | 2.377804 | 1.249807 | 0.740941 | 0.306257 | 0.382763 | 0.226919 |
| 357.61 | 59 | 5904.95 | 94.29 | 115.56 | 69.84 | 1202.3 | 2.67141 | 1.225581 | 0.740694 | 0.306305 | 0.375402 | 0.226878 |
| 281.31 | 28.61 | 4501.78 | 53.12 | 48.25 | 26.9 | 462.08 | 3.334094 | 0.908321 | 0.506401 | 0.378725 | 0.344004 | 0.191787 |
| 68.42 | 17.27 | 1721.72 | 40.1 | 35.42 | 18.28 | 350.73 | 1.801354 | 0.883292 | 0.45586 | 0.393369 | 0.347459 | 0.179321 |
| 96.85 | 12.87 | 2196.76 | 20.45 | 18.21 | 9.17 | 152.74 | 3.077264 | 0.890465 | 0.448411 | 0.394027 | 0.350867 | 0.176686 |
| 65.73 | 19.39 | 2180.44 | 30.89 | 43.39 | 15.11 | 304.28 | 1.391875 | 1.404662 | 0.489155 | 0.321771 | 0.451979 | 0.157396 |
| 128.58 | 44.97 | 3274.86 | 30.07 | 43.67 | 31.11 | 325.25 | 2.785556 | 1.452278 | 1.034586 | 0.267123 | 0.387936 | 0.276361 |
| 96.69 | 38.54 | 2637.92 | 32.45 | 46.22 | 24.29 | 304.21 | 2.152228 | 1.424345 | 0.748536 | 0.290406 | 0.413639 | 0.21738 |
| 82.26 | 19.53 | 2453.87 | 37.84 | 41.78 | 23.72 | 296.08 | 1.935507 | 1.104123 | 0.62685 | 0.338947 | 0.374239 | 0.212469 |
| 121.08 | 34.12 | 3222.35 | 51.24 | 48.22 | 25.29 | 351.73 | 2.44402 | 0.941062 | 0.49356 | 0.376433 | 0.354246 | 0.185792 |
| 52.32 | 8.49 | 2065.7 | 18.35 | 18.72 | 10.16 | 284.95 | 2.226726 | 1.020163 | 0.553678 | 0.355896 | 0.363072 | 0.197052 |
| 156.1 | 40.04 | 2860.79 | 37.47 | 55.51 | 35.42 | 335.09 | 2.829373 | 1.481452 | 0.94529 | 0.268179 | 0.397295 | 0.253507 |
| 147 | 51.33 | 2534.46 | 40.18 | 58.29 | 47.52 | 337.54 | 2.329571 | 1.450722 | 1.182678 | 0.253342 | 0.367528 | 0.299622 |
| 132.22 | 40.48 | 3302.37 | 24.32 | 37.38 | 20.07 | 312.71 | 3.35684 | 1.537007 | 0.825247 | 0.271368 | 0.417094 | 0.223946 |
| 135.57 | 27.55 | 2864.19 | 41.75 | 44.58 | 24.37 | 365.99 | 3.038439 | 1.067784 | 0.583713 | 0.345127 | 0.368521 | 0.201455 |
| 32.37 | 13.6 | 2293.59 | 36.18 | 40.28 | 17.77 | 332.65 | 1.46612 | 1.113322 | 0.491155 | 0.349228 | 0.388803 | 0.171525 |
| 124.81 | 37.09 | 3660.16 | 37.23 | 41.81 | 27.9 | 370.71 | 2.480165 | 1.123019 | 0.749396 | 0.318996 | 0.358238 | 0.239054 |
| 49.32 | 19.59 | 2799.58 | 27.35 | 28.87 | 15.35 | 355.49 | 1.945204 | 1.055576 | 0.561243 | 0.349342 | 0.368757 | 0.196066 |
| 94.42 | 24.84 | 2193.19 | 42.8 | 42.25 | 32.41 | 282.84 | 2.217984 | 0.98715 | 0.757243 | 0.335239 | 0.330931 | 0.253858 |
| 24.12 | 7.6 | 2058.17 | 6.75 | 8.72 | 5.98 | 73.1 | 2.667656 | 1.291852 | 0.885926 | 0.286138 | 0.369648 | 0.253497 |
| 53.43 | 12.62 | 1946.18 | 25.55 | 27.17 | 13.17 | 334.62 | 2.240155 | 1.063405 | 0.51546 | 0.353047 | 0.375432 | 0.181981 |
| 77.18 | 14.2 | 1940.53 | 26.41 | 40.53 | 22.15 | 197.39 | 1.776839 | 1.534646 | 0.838697 | 0.275535 | 0.422848 | 0.23109 |
| 42.57 | 8.32 | 1550.41 | 10.25 | 9.9 | 4.87 | 101.09 | 3.615661 | 0.965854 | 0.475122 | 0.368175 | 0.355603 | 0.174928 |
| 63.31 | 9.53 | 1463.95 | 17.43 | 14.93 | 7.25 | 173 | 2.98032 | 0.856569 | 0.41595 | 0.398856 | 0.341648 | 0.165904 |
| 84.91 | 35.29 | 3131.85 | 46.25 | 47.44 | 27.51 | 451.1 | 2.352675 | 1.02573 | 0.594811 | 0.348978 | 0.357957 | 0.207576 |
| 52.31 | 21.68 | 3089.68 | 26.04 | 25.51 | 12.03 | 218.11 | 2.340929 | 0.979647 | 0.461982 | 0.377939 | 0.370247 | 0.174601 |

| CARB/kcal | FAT/kcal | PROT/kcal | dt_vitd | dt_vitk | dt_vitc | dt_thia | dt_ribo | dt_niac | dt_vitb6 | dt_tfol | fol_dfe | dt_folac |
|---|---|---|---|---|---|---|---|---|---|---|---|---|
| 0.138784 | 0.033525 | 0.042529 | 13.76 | 584.65 | 435.63 | 5.91 | 4.28 | 65.42 | 6.75 | 1254.63 | 1378.59 | 177.32 |
| 0.118479 | 0.044156 | 0.034403 | 6.34 | 101.64 | 44.11 | 2.48 | 2.39 | 27.59 | 2.14 | 594.59 | 766.78 | 250.84 |
| 0.133867 | 0.037626 | 0.035346 | 3.3 | 170.04 | 109.63 | 2.23 | 2.12 | 25.39 | 2.59 | 651.51 | 775.73 | 183.43 |
| 0.120886 | 0.042765 | 0.039154 | 5.2 | 773.31 | 187.49 | 2.27 | 2.03 | 26.35 | 2.91 | 667.2 | 749.66 | 116.94 |
| 0.117037 | 0.040211 | 0.042397 | 4.73 | 131.33 | 65.01 | 1.92 | 1.71 | 23.62 | 1.94 | 416.25 | 555.42 | 198.08 |
| 0.090072 | 0.05252 | 0.041395 | 5.82 | 198.4 | 81.21 | 1.54 | 1.37 | 15.89 | 1.62 | 404.95 | 521.92 | 166.31 |
| 0.121801 | 0.044182 | 0.033044 | 9.63 | 160.07 | 129.95 | 3.3 | 2.48 | 33.32 | 3.27 | 706.49 | 904.58 | 283.13 |
| 0.069805 | 0.024804 | 0.023767 | 3.54 | 34.2 | 33.46 | 1.07 | 0.558 | 10.36 | 0.676 | 113.61 | 141.13 | 39.39 |
| 0.090751 | 0.053898 | 0.038779 | 2.35 | 135.57 | 64.12 | 1.01 | 0.821 | 12.4 | 1.14 | 292.47 | 328.87 | 52.64 |
| 0.108346 | 0.046488 | 0.039608 | 4.11 | 89.76 | 57.47 | 1.69 | 1.64 | 22.26 | 1.89 | 489.22 | 690.58 | 287.78 |
| 0.130992 | 0.038393 | 0.036844 | 5.61 | 158.69 | 116.65 | 3.45 | 3.02 | 37.75 | 3.45 | 624.16 | 844.24 | 315.24 |
| 0.120253 | 0.03558 | 0.03873 | 5.51 | 259.68 | 122.9 | 2.82 | 2.06 | 29.99 | 2.71 | 561.45 | 713.28 | 216.83 |
| 0.117548 | 0.038176 | 0.050163 | 2.93 | 185.51 | 75.67 | 2.22 | 1.67 | 29.43 | 2.41 | 367.49 | 484.42 | 167.17 |
| 0.136825 | 0.040524 | 0.029048 | 1.72 | 239.81 | 99.35 | 1.45 | 0.975 | 14.33 | 1.07 | 334.76 | 401.83 | 94.67 |
| 0.12394 | 0.039559 | 0.038591 | 4.99 | 101.5 | 98.68 | 1.94 | 1.59 | 22.49 | 1.99 | 407.36 | 510.26 | 146.1 |
| 0.098517 | 0.04212 | 0.037364 | 2.14 | 224.55 | 74.96 | 1.19 | 0.898 | 12.82 | 1.34 | 263.92 | 294.22 | 43.66 |
| 0.121572 | 0.042784 | 0.037254 | 10.47 | 257.76 | 86.58 | 3.6 | 2.32 | 32.93 | 3.05 | 577.65 | 717.48 | 199.53 |
| 0.103899 | 0.03902 | 0.04337 | 4.63 | 96.03 | 53.5 | 2.05 | 1.22 | 16.71 | 1.75 | 424.77 | 549.57 | 177.9 |
| 0.109523 | 0.046061 | 0.037952 | 2.55 | 120.59 | 83.17 | 1.94 | 1.23 | 23.85 | 2.17 | 370.45 | 478.98 | 153.27 |
| 0.111446 | 0.041718 | 0.044613 | 12.93 | 319.89 | 306.42 | 6.06 | 5.11 | 86.48 | 6.56 | 1145.14 | 1449.22 | 434.7 |
| 0.12862 | 0.038577 | 0.038239 | 17.99 | 180.25 | 237.07 | 4.49 | 2.69 | 34.6 | 3.33 | 621.91 | 772.29 | 215.14 |
| 0.091886 | 0.051009 | 0.046871 | 4.2 | 84.95 | 97.59 | 2.2 | 1.41 | 21.2 | 2.03 | 363.54 | 467.18 | 148.66 |
| 0.122096 | 0.039677 | 0.039837 | 2.3 | 35.8 | 72.91 | 1.58 | 0.782 | 12.76 | 1.34 | 190.68 | 223.82 | 48.48 |
| 0.085172 | 0.061192 | 0.032655 | 6.41 | 400.3 | 128.41 | 1.32 | 0.854 | 16.02 | 1.72 | 352.69 | 385.13 | 45.97 |
| 0.12185 | 0.043744 | 0.037425 | 5.25 | 548.72 | 225.54 | 2.43 | 2.35 | 26.85 | 3 | 737.72 | 864.55 | 183.57 |
| 0.108078 | 0.050217 | 0.034964 | 3.28 | 414.12 | 193.07 | 1.96 | 1.55 | 21.42 | 2.34 | 531.91 | 579.75 | 70.01 |
| 0.094054 | 0.048594 | 0.0419 | 4.76 | 107.53 | 38.65 | 2.18 | 1.44 | 26.65 | 2.12 | 421.56 | 536.68 | 164.67 |
| 0.113345 | 0.046376 | 0.036847 | 7.06 | 278.85 | 156.85 | 2.57 | 2.2 | 28.18 | 2.87 | 588.14 | 729 | 198.81 |
| 0.097614 | 0.043838 | 0.044356 | 4.8 | 71.96 | 52.52 | 1.52 | 0.774 | 13.81 | 1.14 | 201.79 | 239.08 | 53.53 |
| 0.117707 | 0.041602 | 0.04318 | 9.79 | 194.47 | 134.02 | 3.43 | 2.95 | 45.41 | 3.57 | 710.57 | 905.4 | 281.05 |
| 0.111667 | 0.047934 | 0.038462 | 7.59 | 409.59 | 184.47 | 3.3 | 2.84 | 43.75 | 4.28 | 1052.1 | 1375.36 | 463.42 |
| 0.12684 | 0.037786 | 0.045986 | 10.17 | 770.26 | 208.34 | 2.79 | 1.88 | 28.29 | 3.06 | 655.76 | 722.16 | 94.2 |
| 0.12491 | 0.04111 | 0.035774 | 7.02 | 105.98 | 139.08 | 2.83 | 2.51 | 32.73 | 2.89 | 718.13 | 1042.94 | 464.72 |
| 0.072004 | 0.049112 | 0.045808 | 2.78 | 71.4 | 34.52 | 2.13 | 1.38 | 28.69 | 2 | 315.8 | 381.25 | 93 |
| 0.110417 | 0.04452 | 0.044859 | 5.68 | 499.29 | 244.58 | 2.25 | 1.88 | 32.51 | 2.96 | 625 | 715.26 | 131.68 |
| 0.09309 | 0.047856 | 0.04719 | 5.49 | 158.22 | 83.84 | 1.89 | 1.36 | 19.48 | 1.84 | 364.31 | 441.62 | 110.33 |
| 0.108912 | 0.049104 | 0.033785 | 5.14 | 266.72 | 97.98 | 2.14 | 1.81 | 23.35 | 1.88 | 468.09 | 579.24 | 160.3 |
| 0.114949 | 0.04309 | 0.040861 | 1.77 | 92.49 | 48.97 | 0.531 | 0.495 | 7.49 | 0.676 | 173.03 | 225.95 | 75.67 |
| 0.102532 | 0.04577 | 0.045599 | 2.22 | 81.37 | 40.28 | 1.31 | 1.31 | 20.11 | 1.52 | 263.87 | 342.59 | 112.19 |
| 0.094523 | 0.053197 | 0.032779 | 4.11 | 155.16 | 60.37 | 1.55 | 1.14 | 17.85 | 1.46 | 275.67 | 352.39 | 109.96 |
| 0.124259 | 0.034367 | 0.045477 | 3.27 | 55.57 | 24 | 1.22 | 0.804 | 10.04 | 0.884 | 177.38 | 228.75 | 73.08 |
| 0.116538 | 0.039102 | 0.048238 | 3.64 | 79.88 | 30.54 | 1.29 | 0.837 | 15.54 | 1.22 | 193.18 | 237.06 | 62.56 |
| 0.105912 | 0.045018 | 0.044933 | 4.97 | 208.74 | 92.98 | 2.49 | 2.24 | 34.51 | 2.95 | 571.83 | 698.21 | 181.98 |
| 0.1072 | 0.045794 | 0.040902 | 2.33 | 210.15 | 64.56 | 1.63 | 1.24 | 12.51 | 1.35 | 383.66 | 448.57 | 93.17 |

| dt_folfd | dt_vb12 | dt_zinc | group_fried_fish_fish_sticks_sandwich_breaded_fillets_total_grams FFQ | group_high_omega3_fish_total_grams FFQ | group_other_fish_dishes_low_omega3_total_grams FFQ | group_shellfish_except_oysters_total_grams FFQ | sup_dha FFQ | dha_epa FFQ | (FFQ_EPA+DHA):TFAT | (FFQ_EPA+DHA:SFAT) |
|---|---|---|---|---|---|---|---|---|---|---|
| 1076.4 | 15.24 | 35.1 | 3.76 | 12.23 | | 10.54 | | 0.236 | 0.001346 | 0.004634 |
| 342.85 | 5.9 | 15.21 | | | | 2.26 | | 0.0359 | 0.000249 | 0.000684 |
| 470.37 | 3.9 | 16.22 | | | | | | 0.0428 | 0.000388 | 0.001197 |
| 549.95 | 5.07 | 15.89 | | 14.57 | 14.77 | 10.54 | | 0.219 | 0.002039 | 0.006411 |
| 217.19 | 5.46 | 12.98 | 8.69 | 1.68 | 1.68 | 6.54 | | 0.098 | 0.001219 | 0.003782 |
| 238.17 | 5.51 | 11.68 | 3.76 | 12.23 | 14.77 | 4.57 | | 0.234 | 0.002492 | 0.006822 |
| 422.66 | 8 | 20.8 | 17.54 | | 6.4 | 19.57 | | 0.144 | 0.000806 | 0.002657 |
| 74.27 | 2.28 | 5.11 | 3.76 | 2.62 | 0.981 | 4.29 | | 0.061 | 0.001679 | 0.004758 |
| 239.86 | 2.94 | 8.07 | 1.86 | 5.3 | 3.4 | 10.54 | | 0.105 | 0.001438 | 0.004485 |
| 201 | 5.38 | 12.25 | 1.86 | 7.85 | 1.68 | 6.54 | | 0.13 | 0.001486 | 0.004248 |
| 308.46 | 7.4 | 17.6 | 3.76 | 5.3 | | 4.57 | | 0.117 | 0.001157 | 0.003736 |
| 344.88 | 5.32 | 12.84 | | | 3.16 | 2.26 | | 0.0555 | 0.000687 | 0.002163 |
| 200.51 | 4.84 | 11.65 | | 3.4 | 1.68 | 1.4 | | 0.0762 | 0.001275 | 0.004474 |
| 240.09 | 1.88 | 5.95 | 1.12 | 2.1 | 0.981 | 6.54 | | 0.0539 | 0.001036 | 0.003446 |
| 261.17 | 5.95 | 13.29 | 3.76 | 5.3 | 6.4 | 10.54 | | 0.128 | 0.001492 | 0.004758 |
| 220.23 | 2.76 | 8.22 | | 7.85 | 1.68 | 10.54 | | 0.119 | 0.002178 | 0.008512 |
| 376.91 | 6.87 | 19.16 | 4.48 | 4.55 | 4.55 | 20.04 | | 0.183 | 0.00129 | 0.003952 |
| 247.06 | 5.13 | 9.74 | 9.71 | 2.62 | 3.16 | 2.26 | | 0.113 | 0.001698 | 0.005376 |
| 217.26 | 4.22 | 9.78 | | 1.68 | 1.68 | 3.08 | | 0.0607 | 0.000717 | 0.00234 |
| 713.06 | 24.32 | 61.88 | 98.79 | 1.68 | 54.86 | 62.32 | 17.14 | 0.847 | 0.002752 | 0.008983 |
| 406.62 | 11.34 | 23.42 | 7.6 | | | | | 0.0648 | 0.000462 | 0.00122 |
| 214.38 | 5.12 | 12.85 | | | | | | 0.0374 | 0.000367 | 0.000933 |
| 142.02 | 3.32 | 7.73 | | | | 4.29 | | 0.0265 | 0.000511 | 0.001296 |
| 307.24 | 3.77 | 7.27 | | 29.14 | 14.77 | 2.26 | | 0.341 | 0.003552 | 0.011039 |
| 553.72 | 4.06 | 15.22 | | | 6.4 | 2.26 | | 0.0623 | 0.000553 | 0.002072 |
| 461.78 | 3.28 | 12.63 | | 12.23 | | 2.26 | | 0.158 | 0.001414 | 0.004869 |
| 257.19 | 5.07 | 13.27 | | | | | 120 | 0.0354 | 0.000317 | 0.000936 |
| 389.16 | 7.08 | 16.2 | | 5.3 | 6.4 | 2.26 | | 0.126 | 0.000926 | 0.002459 |
| 148.09 | 3.71 | 6.76 | | 7.85 | 3.4 | 10.54 | | 0.144 | 0.002793 | 0.007847 |
| 429.39 | 8.24 | 20.7 | 38.86 | 22.71 | 6.4 | 19.57 | | 0.415 | 0.00297 | 0.011076 |
| 588.76 | 7.47 | 25.11 | | 21.23 | 3.16 | 20.04 | 17.14 | 0.296 | 0.001866 | 0.007367 |
| 561.72 | 6.81 | 15.94 | 5.23 | 18 | 25.78 | 17.33 | 120 | 0.327 | 0.003649 | 0.013446 |
| 253.01 | 6.3 | 17.35 | 3.77 | 3.4 | 0.981 | 2.83 | 87.99 | 0.096 | 0.000794 | 0.002299 |
| 222.84 | 5.35 | 15.37 | | | 3.16 | 4.29 | | 0.0788 | 0.000761 | 0.002178 |
| 494.73 | 4.83 | 15.83 | 3.76 | 2.62 | 3.16 | 8.68 | | 0.153 | 0.001311 | 0.00411 |
| 253.99 | 5.48 | 10.63 | 1.86 | 7.85 | 29.14 | 1.4 | 22 | 0.218 | 0.002785 | 0.007971 |
| 307.54 | 4.38 | 11.93 | 7.6 | 12.23 | 3.16 | 2.83 | | 0.172 | 0.001347 | 0.004019 |
| 97.41 | 1.67 | 4.79 | 1.86 | 5.3 | 6.4 | 2.83 | 75 | 0.086 | 0.003646 | 0.012741 |
| 152 | 3.2 | 9.65 | | | | 2.26 | | 0.059 | 0.000815 | 0.002309 |
| 166.09 | 4.52 | 9.14 | 1.86 | 7.85 | 1.68 | 1.4 | | 0.124 | 0.001294 | 0.004695 |
| 104.51 | 3.51 | 6.22 | 1.12 | 2.1 | 4.58 | 1.4 | | 0.0466 | 0.001674 | 0.004546 |
| 130.53 | 2.94 | 7.51 | | 2.62 | | 2.26 | | 0.0612 | 0.0014 | 0.003511 |
| 388.78 | 7.14 | 19.58 | 3.76 | 2.62 | 21.23 | 20.04 | | 0.179 | 0.001351 | 0.00387 |
| 290.47 | 3.08 | 10.38 | 1.86 | | 1.68 | 0.659 | | 0.0312 | 0.000453 | 0.001198 |
